# Sex differences in circulating metabolites across glycemic status and risk of coronary heart disease

**DOI:** 10.1101/2024.07.23.24310540

**Authors:** Yilin Yoshida, Danting Li, Xiang Li, Vivian A. Fonseca, Lu Qi, Franck Mauvais-Jarvis

## Abstract

**Background:** Women with type 2 diabetes (T2D) have a 50% excess risk of coronary heart disease (CHD) than men with T2D. We compared circulating metabolites and their associations with CHD in men and women across glycemic status.

**Methods:** We used metabolomic data (lipoproteins, fatty acids, amino acids, glycolysis, ketones, inflammation, and fluid balance) for 87,326 CHD-free UK Biobank participants. We used linear regressions to examine the association of sex and metabolites (log) in newly diagnosed T2D (diagnosis<2 yrs from baseline), prediabetes (A1c 5.7-6.5%), and euglycemia, accounting for age, race, Deprivation Index, income, smoking, alcohol drinking, obesity, physical activity, medications for hypertension, hyperlipidemia, and diabetes. We used Cox models to evaluate the association of metabolites and CHD risk by sex, adjusting the same covariates and menopausal status (women). All analyses were FDR-adjusted.

**Findings:** We included 1250 individuals with new T2D, 12,706 with prediabetes, and 83,315 with euglycemia. In adjusted linear regressions, women showed a progressive increase in atherogenic lipid and lipoprotein markers and inflammatory marker, glycoprotein acetyls, compared to men as their glycemic status advanced. However, women had lower levels of albumin during this transition. Menopausal status did not alter these sex differences. In a 10-year follow-up, an SD higher total TG, TG in VLDL, LDL, and HDL, saturated fatty acids (SFA) were positively associated with a higher risk of CHD in women with T2D but not in men (p-interactions 0.03-0.15).

**Interpretation:** With advancing glycemic status, women exhibited higher levels of atherogenic lipids and lipoproteins, as well as inflammatory markers, but lower circulating albumin. Women with T2D appear to be at a higher risk of CHD associated with TG, VLDL-TG, LDL-TG, and HDL-TG, and SFA than men with T2D.

## Background

Women with type 2 diabetes (T2D) face a 50% greater excess risk of coronary heart disease (CHD) compared to men.^1^ In the transition from normoglycemia to T2D, women have a greater and faster degradation of cardiometabolic risk factors compared to men, including greater worsening of atherogenic lipids, which may account for the heightened risk of CHD in women with T2D. ^2, 3^ However, little has been done to characterize sex differences in subtle and progressive changes in metabolic biomarkers along the diabetic continuum.

Nuclear magnetic resonance metabolome profiling (metabolomics) provides detailed information on metabolic markers, including lipoprotein particle concentrations, compositions, size, lipid species, amino acids, ketones, and metabolite intermediates of glycolysis or fluid balance. ^4^

Metabolomics can provide comprehensive and accurate biomarker profiling to detect sex differences associated with the pathogenesis of T2D and associated CHD across glycemic status (i.e., euglycemic, pre-diabetes [pre-DM], and T2D). Leveraging blood metabolome data from a large prospective cohort, we sought to compare levels of lipids, lipoproteins, and other metabolites in men and women from groups of normoglycemia, pre-DM to newly diagnosed T2D. We examined the associations between metabolites and incident CHD in women and men across glycemic status. Because estrogen improves the metabolic risk profiles and may underpin the sex differences in metabolic risk profiles, we compared the sex differences of serum metabolites by menopausal status.

## Methods Data Source

This study is based on the UK Biobank. UK Biobank is a large prospective observational cohort study that has recruited over 500,000 adults across 22 centers located throughout the United Kingdom. The full protocol of the UK Biobank study is publicly available, and the study design and measurement methods have been described elsewhere. ^5^ Briefly, participants aged 40–69 were enrolled between 2006 and 2010, and most participants agreed to participate and attended the baseline assessment, which included questionnaires on lifestyle, medical history, and physical and functional measurements. ^5^ Additionally, blood, urine, and saliva samples were taken. ^5^ All participants provided written informed consent. We included a total of 87,326 CVD-free participants with known glycemic status and baseline plasma biomarker measurements by NMR spectroscopy. (Supplemental Figure 1 for sample selection)

### Measurements

Metabolite quantification: Detailed protocols on sample collection and metabolomic quantification are presented elsewhere. ^4^ Briefly, EDTA plasma samples were collected at baseline recruitment and repeat assessments. Samples were prepared directly in 96-well plates by UK Biobank, with each plate containing a serum mimic as a quantification consistency monitor and a mixture of 2 low-molecular-weight metabolites as a technical reference. 118,461 plasma samples were measured at Nightingale Health’s laboratories in Finland between June 2019 and April 2020. Accredited quality control was done during the whole process to eliminate systemic and technical variance, and only samples and biomarkers that underwent the quality control process were stored in the UK Biobank dataset and used in this study. Each sample included 168 metabolites in absolute level (mmol/L) spanning fatty acids, glycolysis metabolites, ketones, amino acids, lipids, and lipoproteins, and 81 in ratio measurement. ^4^

Glycemic status: Participants self-reported their medical history of diabetes, including age at first diagnosis of diabetes and the use of diabetic medications. We defined newly diagnosed T2D as those with a duration of T2D ≤2 years. Pre-DM was defined by HbA1c 5.7-6.4 without T2D diagnosis or anti-diabetic medication.

Outcome: Incident CHD was defined by information from the International Statistical Classification of Disease and Related Health Problems (ICD)-10 for fatal or non-fatal myocardial infarction (I21-I21.4, I21.9, I22-I22.1, I22.8, I22.9, I23-I23.6, I23.8, I24.1, and I25.2. Outcomes adjudication involved linkage with hospital admissions data from England, Scotland, and Wales and the national death register to identify the date of the first known MI after the date of baseline assessment. ^6^ Follow-up started at inclusion in the UK Biobank and ended on January 31, 2021, date of death, or upon the occurrence of outcome for all included participants.

Covariates: Age, sex, and ethnicity were obtained at baseline. Townsend deprivation index (based on the participant’s postcode; a higher score indicates a higher degree of deprivation) was obtained from local National Health Service Primary Care Trust registries and the name of the recruitment center. Average household income and use of medications (anti-diabetic, lipid-lowering, and antihypertensive) were assessed by the touch screen questionnaire during the initial assessment center visit. The International Physical Activity Questionnaire was used to derive minutes per day of moderate-to-vigorous physical activity (MVPA) and converted to the score in metabolic equivalent of task (MET) minutes/day for comparison. We used MET 150 mins/week at a cut-off for recommended physical activity. ^7^ Alcohol intake was self-reported as never, special occasions only, 1 to 3× per month, once or twice per week, 3 to 4× per week, and daily or almost daily. Smoking status was also self-reported, classified as a current smoker or non-smoker (former or never). Height and body weight were measured during the initial assessment center visit; body mass index was calculated as weight in kilograms divided by squared height in meters (kg/m^2^). Apolipoprotein B (Apo B), apolipoprotein A (Apo A), LDL cholesterol (LDL-c), HDL cholesterol (HDL-c), triglycerides (TG), total cholesterol, and albumin were measured in the blood sample collected at baseline. Details on serum sample handling and assays in the UK Biobank have been described previously. ^8^ Specifically, a series of biological samples comprised 45 ml of blood and 9 ml of urine. Serum lipid traits were measured by immunoturbidimetric analysis on a Beckman automated hematology analyzer (Beckman Coulter AU5800). Serum albumin levels were estimated using a colorimetric approach and a Beckman Coulter AU5800 assay (Beckman Coulter, United Kingdom) with an analytical range of 15–60 g/l.

### Analysis

We used multivariable linear regressions to examine the associations between sex and metabolites adjusted for age, race, income, area deprivation, smoking, alcohol drinking, physical activity, BMI, and medications for hypertension, hyperlipidemia, and diabetes. β coefficients for sex were reported for each biomarker in each model, with positive values representing higher relative levels in women. The magnitude of the association of the biomarker with sex can be inferred from the absolute value of β coefficients. We calculated the adjusted means of each metabolite by sex and glycemic status. We evaluated the influence of menopausal status on metabolite changes by comparing sex differences of pre- or postmenopausal women with those of their age-matched men. We conducted a sensitivity analysis in those without lipid-lowering medications at baseline. We also validated the results using clinical chemistry-measured biomarkers, including Total cholesterol, LDL-c, HDL-c, triglycerides, Apo B, Apo A, and albumin. The false discovery rate (FDR) method was used to adjust for multiple comparisons. We used Cox proportional hazard models to separately evaluate the association between metabolites and CHD risk in men and women, adjusting the same covariates.

## Results

A total of 83,315 individuals with euglycemia (56% women), 12,706 individuals with pre-DM (56% women), and 1250 individuals with newly diagnosed T2D (40% women) were included in the analysis. Participants in the T2D group were older and had a higher proportion of non-Whites participants from low-income backgrounds, with obesity, and with a history of lipid-lowering and anti-hypertensive medication use. (Table 1. Baseline Characteristics)

**Table 1.**
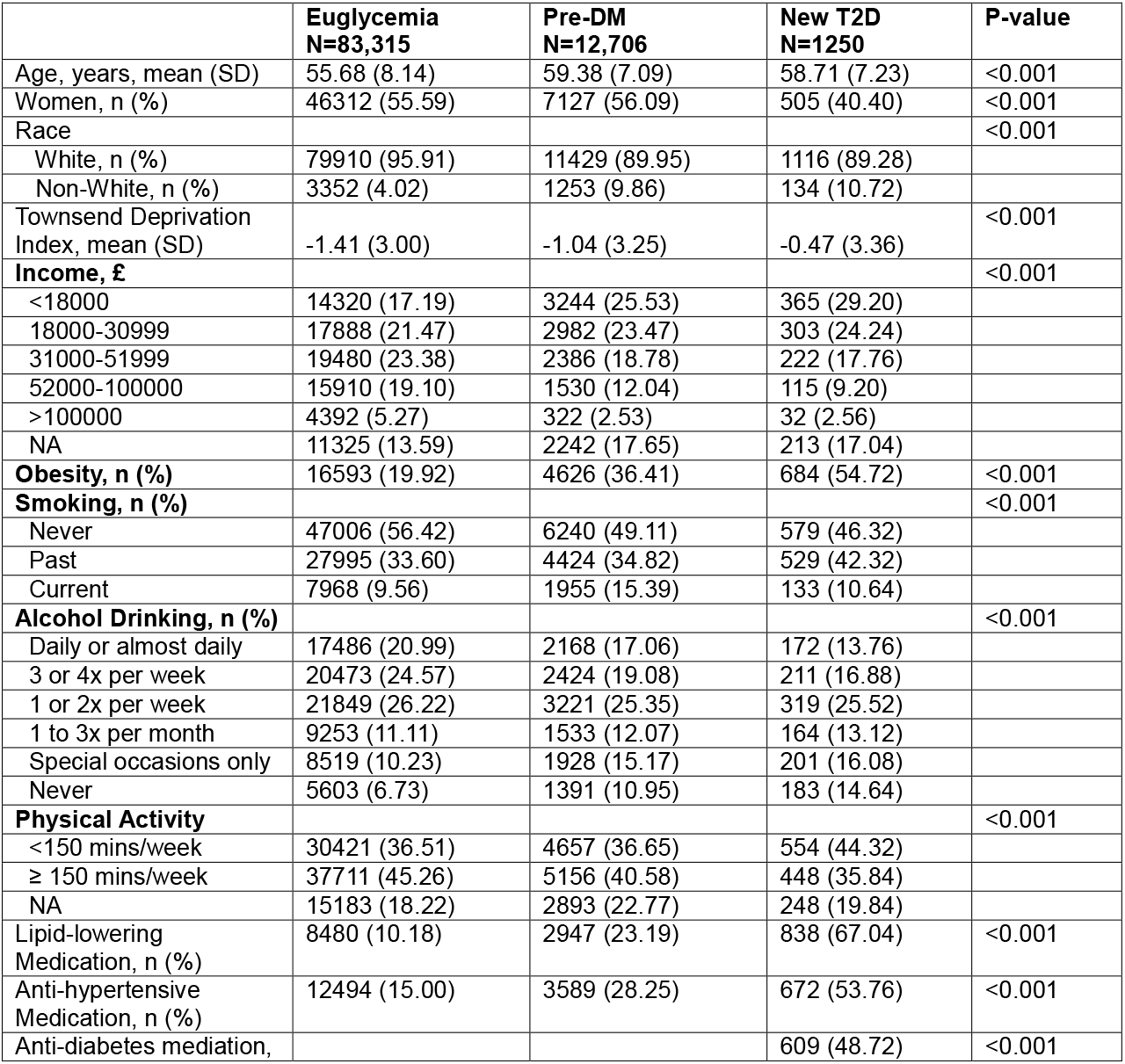
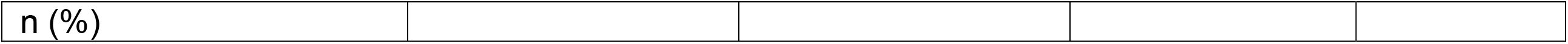
Baseline Characteristics By Glycemic Status.

### Lipoproteins, Lipids, and Fatty Acids

In adjusted linear regressions (Figure 1-Heatmap 1 and Supplemental Table 1), women had statistically significantly higher concentrations of total cholesterol, non-HDL-c, remnant-c, LDL-c, total free cholesterol, and total phospholipids and free cholesterol and phospholipids in LDL than men across all glycemic groups. These sex differences were most striking in the newly diagnosed T2D group. Men had higher levels of TG across lipoprotein particles and a higher level of Apo B in the euglycemia group (Figure 1-Heatmap and Supplemental Table 1). In contrast, in the T2D group, women had higher TG concentrations in LDL, HDL, and Apo B than men (Figure 2 volcano plot). Women also had higher levels of HDL-c, Apo A1, and higher lipids concentrations in HDL, such as cholesteryl esters, free cholesterol, total lipids, and phospholipids in HDL; however, the favorable lipid traits in women became less evident with advancing glycemic status (Figure 1-Heatmap 1 and Supplemental Table 2). The average diameter for VLDL particles was bigger in men, while the average diameter for HDL particles was bigger in women across the glycemic status. Total concentrations of lipoprotein particles were slightly higher in women than men with T2D (Figure 1-Heatmap 1 and Supplemental Table 2). Women had higher levels of nearly all fatty acids and fatty acid ratio measures across the glycemic groups. Among fatty acids measures, women’s higher total fatty acids, omega-6 fatty acids, polyunsaturated fatty acids (PUFA), saturated fatty acids (SFA), and linoleic acid relative to men became more striking with the advancement of glycemic groups. (Supplemental Table 1 and Figure 1 Heatmap; Figure 2 volcano plot).

**Figure 1.**
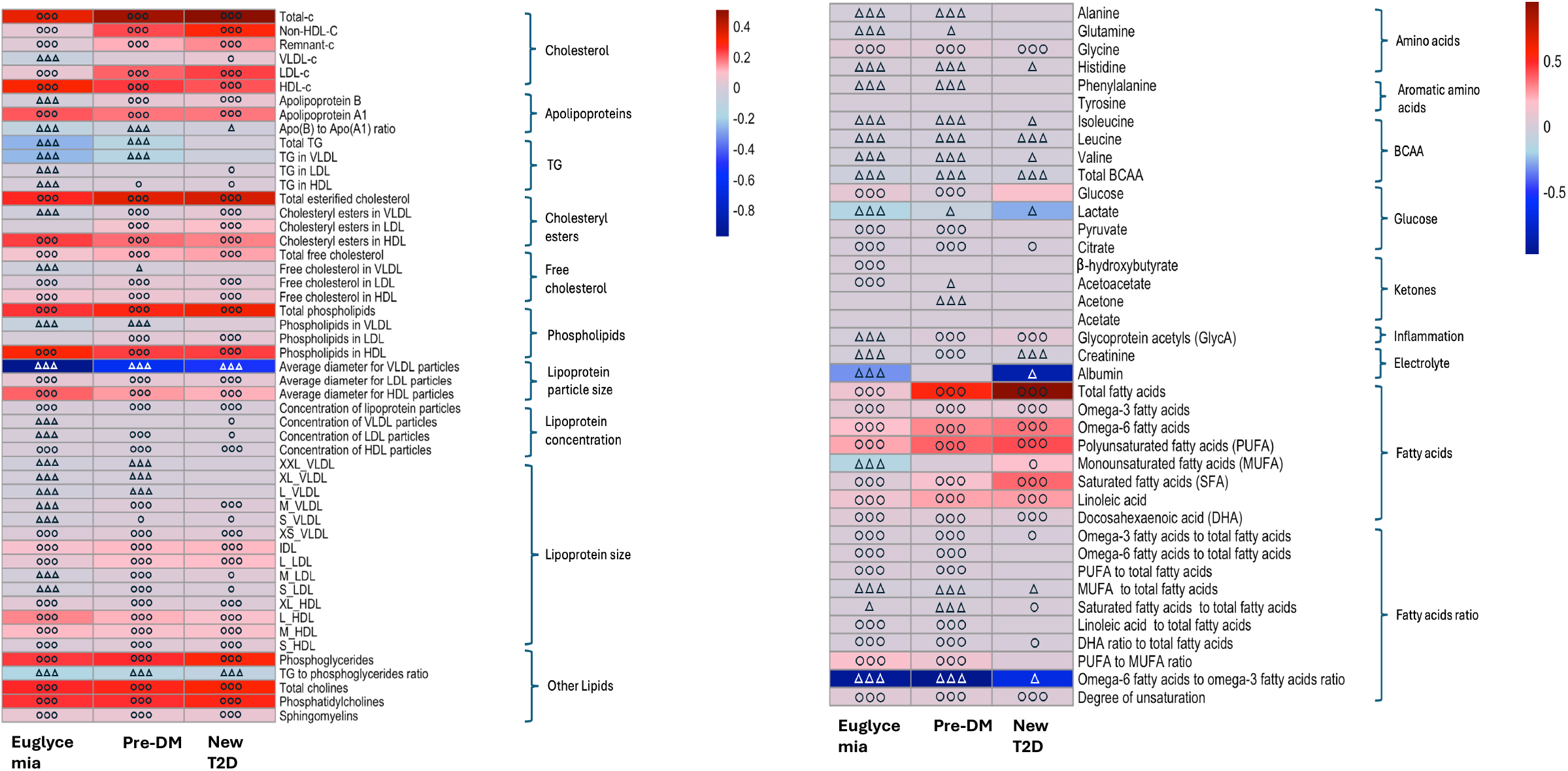
Heatmap of Sex Differences in Metabolites Across Glycemic Status. Positive adjusted β coefficients (red) represent biomarkers that are higher in women ○○○ P<.0001; ○○ <.001; ○<.05 and negative β (blue) represent biomarkers higher in men. ΔΔΔ FDR-adjusted P<.0001;ΔΔ <.001; Δ<.05. Darker shade indicates larger coefficients (i.e., greater sex difference)

**Figure 2.**
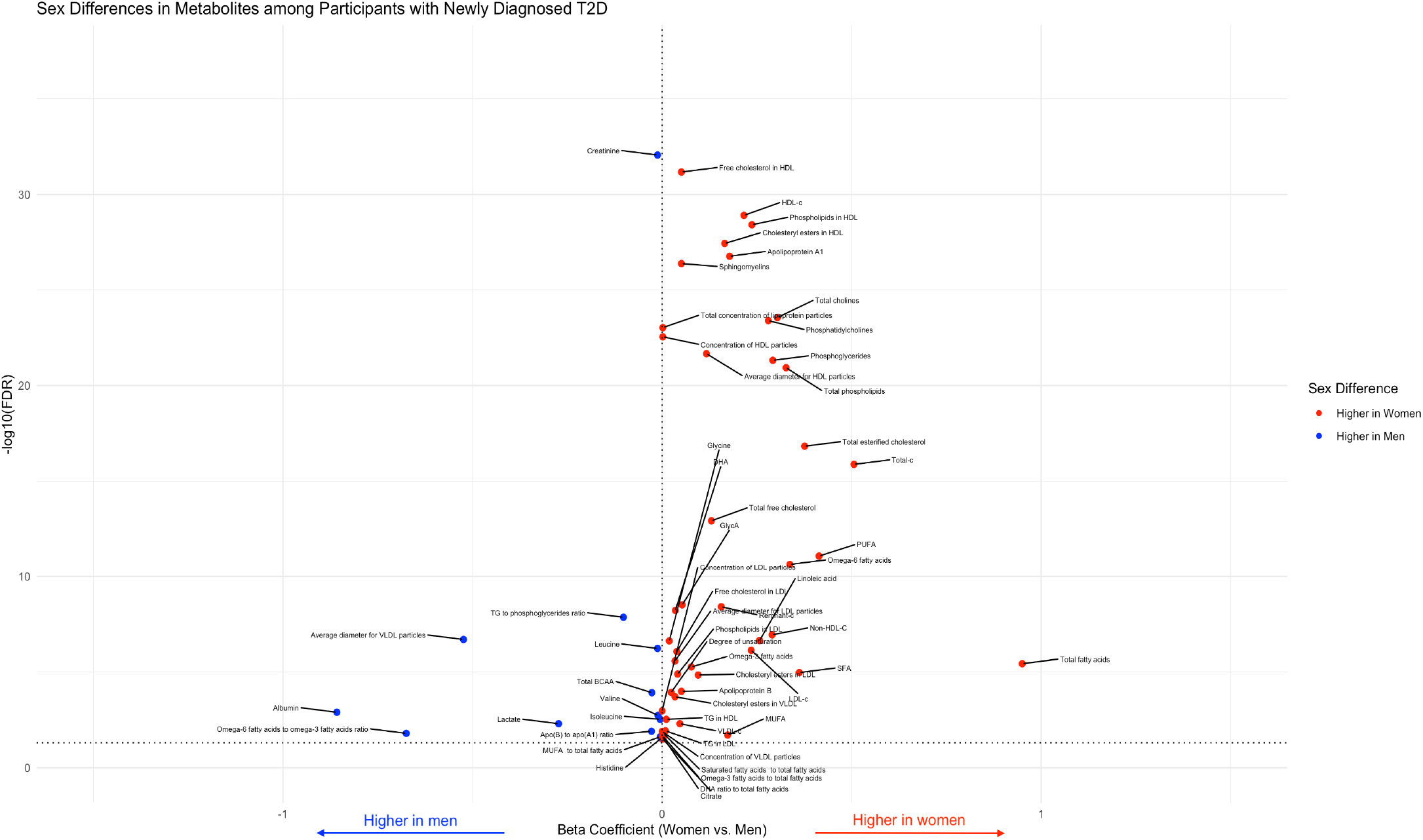
Volcano Plot for Metabolites Significantly Differed (FDR-adjusted p<.1) between Men and Women with Newly Diagnosed T2D. Positive β coefficients (red) represent biomarkers that are higher in women and negative β (blue) represent biomarkers higher in men. The results shown were FDR-adjusted P<.1

### Renal function, glycolysis, inflammation, amino acids, and ketones

Renal function markers, albumin and creatinine were consistently higher in men than women across the glycemic groups, and these sex differences were the most striking in the T2D group (Figure 1-Heatmap 1 and Supplemental Table 2; Figure 2 volcano plot). Glycolysis-related metabolites, including glucose, pyruvate, and citrate, were all higher in women across the glycemic groups, except for lactate, which was higher in men, particularly in the T2D group (Figure 1-Heatmap 1 and Supplemental Table 2; Figure 2 volcano plot). Glycoprotein acetyls (GlycA), an inflammatory marker, ^9^ was higher in men in the euglycemia group, while it was higher in women in the pre-DM and T2D groups (Figure 1-Heatmap 1 and Supplemental Table 2; Figure 2 volcano plot) (Supplemental Table 1 and Figure 1 Heatmap). In terms of amino acids, men with pre-DM or euglycemia had statistically significantly higher levels of alanine, glutamine, and histidine, but the sex differences were absent in the T2D group. Women had higher glycine levels across glycemic status (Supplemental Table 1 and Figure 1 Heatmap).

Men had higher levels of branched-chain amino acids (BCAA), such as isoleucine, leucine, valine, and total BCAA (Supplemental Table 1 and Figure 1 Heatmap). No sex differences were observed among ketones across the glycemic status.

### Stratified analyses by menopause

When stratifying menopausal status (Supplemental Table 2 and Supplemental Figure 2), we found similar sex differences in lipids, lipoproteins, and other metabolites across glycemic status in the postmenopausal group compared with all participants. In comparison between pre-menopausal women and matched young men, we found the estrogen-protective effect on metabolomics profiles was only significant in euglycemic women. Generally, premenopausal euglycemic women had lower levels of atherogenic lipid markers, such as total and non-HDL cholesterol, remnant cholesterol, VLDL cholesterol, TG, Apo B, and lipids concentrations in different lipoprotein particles, as well as lower levels of fatty acids and amino acids except glycine. However, men and women in the premenopausal group with T2D had similar atherogenic lipid traits.

In the sensitivity analysis, excluding those with lipid-lowering medications, results remain similar to the main analysis (Supplemental Table 4).

### Association between Metabolites and Risk of CHD in Men and Women across Glycemic Groups

In a mean follow-up of 10 years, we evaluated the sex-specific association of metabolites and the risk of CHD. Adjusted Cox proportional hazard regression showed that total TG (hazard ratio [HR] 2.5, 95% CI 1.1-5.8 in women and 1.1, 0.7-1.9 in men, respectively, p-interaction 0.11) and TG in VLDL (2.1, 1.0-4.1 in women and 1.1, 0.7-1.7 in men, p-interaction 0.14), TG in LDL (3.1, 0.9-10.7 in women and 1.2, 0.5-2.4 in men, p-interaction 0.15), and TG in HDL (3.3, 1.0-10.5 in women and 1.0, 0.5-1.9 in men, p-interaction 0.07) were positively associated with increased risk of CHD in women with T2D but not in men. In the euglycemia group, the association between total TG or TG cargo in VLDL, LDL, and HDL were all positively associated with the risk of CHD (HRs ranged from 1.1 to 2.0), and the magnitude of the associations appeared to be stronger in women than in men (p-interaction ranged from 0.01 to 0.16). (Figure 3.A. Forest plot, Supplemental Table 3).

**Figure 3. A.**
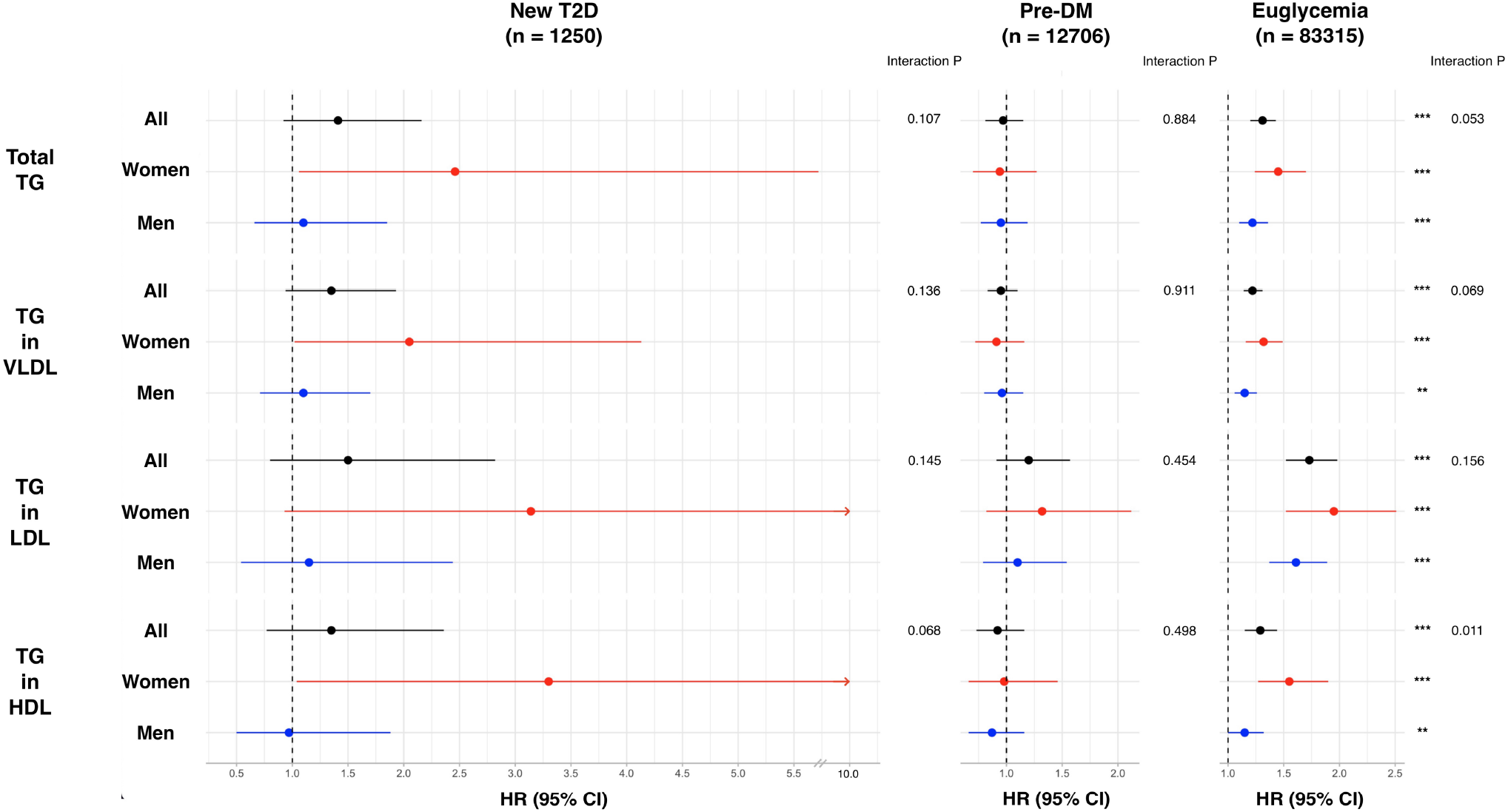
Hazard Ratios (95% CI) of Metabolites and Risk of CHD by Sex Across Glycemic Status.

SFA was associated with increased risk of CHD in both sexes in the euglycemic group (1.4 and 1.8 in men and women, respectively); however, in the T2D group, the association was only significant in female patients (0.9, 0.4-2.0 and 5.2, 1.2-21.9, respectively, p-interaction=0.033) (Figure 3.B. Forest plot). Ketones also showed sex differences in relation to the risk of CHD (Figure 3.B. Forest plot). The increase in acetone was significantly associated with an increased risk of CHD in men with newly diagnosed T2D (2.3, 1.1-4.9), but the association was inverse in diabetic women (0.11, 0.02-0.56, p-interaction 0.001) (supplemental Table 3). A negative association between β-hydroxybutyrate and risk of CHD was also found in women with T2D only (0.68, 0.48-0.95, p-interaction 0.001).

**Figure 3. B.**
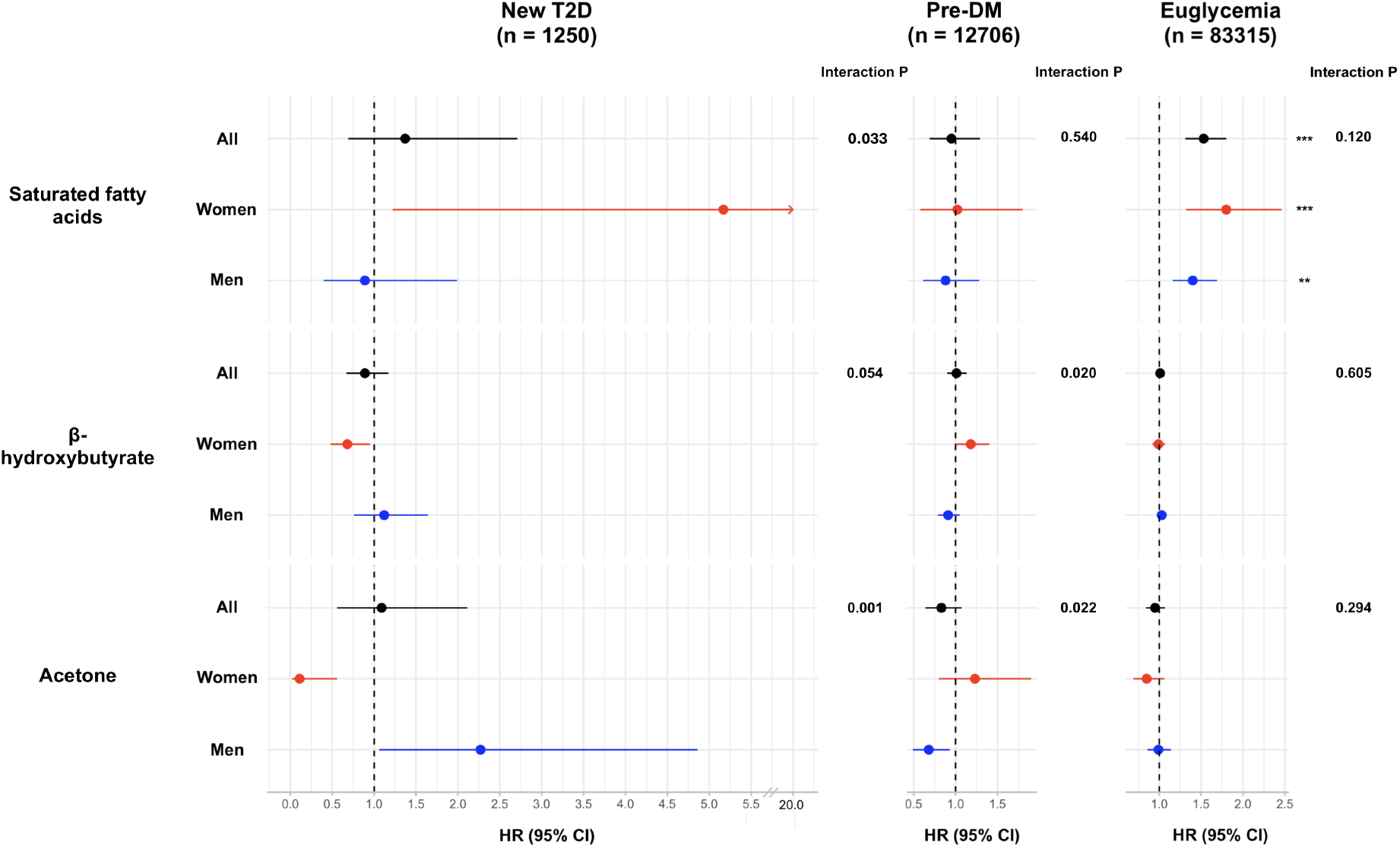
Hazard Ratios (95% CI) of Metabolites and Risk of CHD by Sex Across Glycemic Status.

Our replicated analysis using available biochemistry-measured lipoprotein markers generally showed consistent findings with the metabolomics analysis (Supplemental Table 5). In our sensitivity analysis, excluding those with lipid-lowering medications, results remain similar to the main analysis (Supplemental Table 6).

## Discussion

Leveraging a large population-based cohort and a wide range of molecular biomarkers not commonly measured in clinical practice, we characterized sex differences in metabolic derangement across the glycemic status. These findings provide additional mechanistic insights into women’s excess risk of CHD in diabetes. Our principal findings are threefold. First, women have a progressive increase in atherogenic lipid markers compared to men as glycemic status is advanced. Meanwhile, women also have lower levels of serum albumin during this transition.

Second, we found that in the premenopausal group, the sex difference in CHD risk metabolites, such as atherogenic lipids, did not differ in the T2D group, supporting the concept that diabetes attenuates the female biological advantage from estrogens. Third, we observed a positive association between TG, SFA, and CHD risk, respectively, in women but not in men in this cohort.

Previous studies have reported women’s worse lipid profiles than men at or before the T2D diagnosis. ^2, 3 10^ However, these studies relied on traditional lipid panels and could not distinguish between lipoproteins’ size, density, and compositions. These features have varying relevance to the risk of CHD. ^11^ Our study extended the evidence of women’s greater worsening of lipid profiles transitioning from normoglycemia to T2D than men by including detailed lipid and lipoprotein examinations by NMR. Specifically, the atherogenic lipoproteins, LDL and VLDL, and lipid concentrations (e.g., total lipid, cholesterol, and TG) in these lipoprotein particles were significantly higher in women than men in the newly diagnosed T2D group. The sex differences in lipids and lipoproteins changes influenced by hyperglycemia and insulin resistance are incompletely understood. It was speculated that the greater adverse effect of hyperglycemia on lipoprotein concentrations in women may be related to a greater rate of entry of TG-rich VLDL into the circulation in women. A greater generation of HDL and LDL would likely result from increased VLDL entry. ^12^ Because of these differences, an equivalent reduction in insulin action on lipoprotein lipase–mediated removal of VLDL TG from the circulation, among other possible effects, may be responsible for greater lipoprotein abnormalities in women than in men. ^12^ Additionally, the changes in lipoprotein size and subclass particle concentrations were positively associated with the degree of insulin resistance and adiposity. ^13^ This adiposity/insulin-resistant metabolic disarray that is at the root of this diabetes phenotype could, itself, lead to lipid abnormalities regardless of hyperglycemia. It has been reported that women are more centrally obese at T2D diagnosis. ^1^ The sex differential adiposity phenotype and associated adverse metabolic effects serve as another possible explanation for women’s progressive lipid and lipoprotein worsening with increased glycemia relative to men. Further, gender differences in lipid risk screening and treatments play a role in women’s unfavorable lipid and lipoprotein profiles at the onset of T2D. Previous studies suggested that women, despite having higher LDL-c, were less frequently receiving statin therapy, particularly high-intensity therapy, than men.^14^

Another key observation of our study is that men had higher levels of serum albumin, and the advancement of glycemic status augments the sex difference. Serum albumin is the most abundant protein in circulation and exhibits significant antioxidant, anti-inflammatory, and anticoagulant activity. Low levels of albumin independently predict the risk of CVD. ^15^ In diabetes, albumin synthesis and secretion are decreased due to insulin deficiency. Meanwhile, lower serum albumin is associated with glycation of albumin and hemoglobin and accelerates the onset of diabetes. In our study, men appeared to have the advantage of preserving serum albumin with increased glycemia compared to women. The higher serum albumin level in men than in women was also reported in populations with or without T2D or glucose intolerance. ^16^ Differences in nitric oxide metabolism and oxidative stress between women and men, as well as the influence of sex hormones, have been proposed as explanations for the sex differences in albumin. ^17^ However, the mechanism for the sex difference in albumin influenced by the progression of hyperglycemia is ill-defined and warrants further investigations.

It has been proposed that diabetes nullifies the protective effects of estrogens on women’s cardiovascular health. ^18, 19^ Our study extends the evidence by showing the attenuating effect of menopausal status on a comprehensive metabolic risk profile in pre-menopausal women with T2D. Diabetes impairs endothelial response more detrimentally in women than in men by interrupting the hemodynamic regulation of estrogen through complex interactions between insulin and estrogen signaling. ^20^ Estrogens are also known for their vital role in maintaining energy metabolism, body composition, and regulating lipid metabolism in women via estrogen signaling pathways. ^20^ Hyperglycemia significantly disrupts estrogen action, accelerating the atherogenic process in women with T2D. In this study, we found that estrogen’s beneficial effect on lipoprotein and lipid metabolism was attenuated in premenopausal women with T2D, indicating the detrimental interaction between T2D and estrogens in young women.

Our study highlights that TG concentration overall and in different lipoprotein particles are associated with CHD risk in women but not in men with T2D, even accounting for concomitant risk factors, including lipid-lowering treatment. TG is one of the leading predictors of atherosclerotic CVD among patients with diabetes, comparable to LDL-C, which exceeds the predictive power of HbA1c ^21^ and independent of prior diagnosis of CVD. ^22, 23^ Recent evidence also demonstrated the promising role of TG in LDL as a marker of atherogenic remnant lipoproteins in predicting the risk of atherosclerotic CVD in general European populations. ^24^ However, specific differences in TG in relation to CHD risk between men and women with or without T2D have been noted rarely. Our study demonstrated sex as a significant effect modifier to the link between TG and CHD in a group of newly diagnosed T2D. Hypertriglyceridemia is the most common serum lipid abnormality in people with diabetes. The enhanced detrimental effects of TG in women than men observed in our study may be of significant clinical implication. There remains a gap in effective approaches to modifying the CHD risk in patients with diabetes who have sustained moderate hypertriglyceridemia despite appropriate lifestyle modifications and statin-optimized LDL.^25^ The potentially sex-varying association between TG and diabetic CHD can stimulate further research to validate the need for sex-specific diagnostic and prognostic cut-off of TG in people with T2D. We also want to stress the importance of TG in the development of CHD in nondiabetic populations. As indicated in our results, TG was significantly associated with CHD risk in the normoglycemic group. TG elevation is considered one of the major causes of lipid-dependent residual CVD risks. ^26^ In the normoglycemic group, our results pointed to the risk of CHD preferentially in women than in men. Despite unclear mechanisms for these sex differences, a previous report from the Fringminham Heart Study hinted at a stronger correlation between TG and CVD in women than in men. ^22^

Women with T2D had higher levels of fatty acids, including heart-protective PUFA and MUFA, and heart-risk SFA. This is consistent with previous research reporting that women generally have higher fatty acid species in this study than men. ^27^ Interestingly, the positive association between SFA and CHD was only significant in women with T2D. Meanwhile, in the normoglycemic group, CHD risk associated with SFA generally showed a similar direction and significance in men and women. However, sex-disaggregated examinations of SFA have been scantly reported. Our results indicated a potentially augmented effect of SFA on CHD risk in women along the diabetes continuum.

Another interesting finding worth noting is that the ketone marker acetone was associated with an elevated risk of CHD in men with T2D but not in women with T2D, while β-hydroxybutyrate appeared to be a protective ketone metabolite for women with T2D. Ketones are primarily generated in the liver from fatty acids and delivered to extrahepatic tissues. Ketone metabolism increases in various physiological states such as pregnancy, exercise, and fasting ^28^ and potentially implicated in numerous human disease states relevant to cardiovascular disease, including diabetes and atherosclerosis. ^29^ Despite the ketones’ effect in reducing oxidative stress and mitochondrial uncoupling effect, their association with CHD has been limited and results remained inconsistent. ^30 31^ It is currently unknown if acetone and β-hydroxybutyrate exert a direct role in CVD pathogenesis. Very few analyses examined the sex differences in acetone, β-hydroxybutyrate, and its association with CHD with or without diabetes. A recent study examined overall ketone metabolites and found no heterogeneity of ketone bodies and heart failure by diabetes or sex groups. ^31^ The potential sex differences in acetone in relation to CHD in the context of T2D, such as the interplay of ketone bodies and insulin reaction as well as therapeutic modulation of ketones (for instance, via SGLT2i or very low carbohydrate diets), among other explanations, are worthy of further investigation. ^32^

The study’s strengths include unprecedented sample size with NMR metabolome profiling, prospective design, and extensive phenotypic details on the participants. However, several limitations of the study merit mentioning. First, no causation or mechanism can be made due to the study’s observational nature. UK biobank participants are predominantly white; therefore, generalization to other diverse populations is unknown. Further, when conducting the analysis, metabolome data was only available for serum samples at the UK biobank baseline. We, therefore, cannot take into account the sex differences in changes in metabolites over time.

Future studies, with the ability to include multiple metabolomics assessments, can further substantiate our findings.

## Conclusion

Based on the metabolomics data from a large cohort, we demonstrated sex differences in lipids and lipoproteins and metabolites representing electrolytes/fluid balance, fatty acids, glycolysis, inflammation, amino acids, and ketone bodies across glycemic status. Overall, women had progressively worsening metabolic risk profiles represented by increased atherogenic lipids, inflammation markers, and lower serum albumin. The protective effect of estrogen was greatly attenuated in young participants with T2D. Further, TG, fatty acids, acetone, and β-hydroxybutyrate are differentially associated with the risk of CHD in men and women with T2D. In sum, our findings highlight that women’s metabolic risk profiles were in a worse shape before and at a new diagnosis of T2D, predisposing them to a heightened risk of CHD than men in the later course of T2D.

## Data Availability

All data produced are available online at
Uk Biobank.

https://www.ukbiobank.ac.uk/

---

**Supplemental Figure 1.**
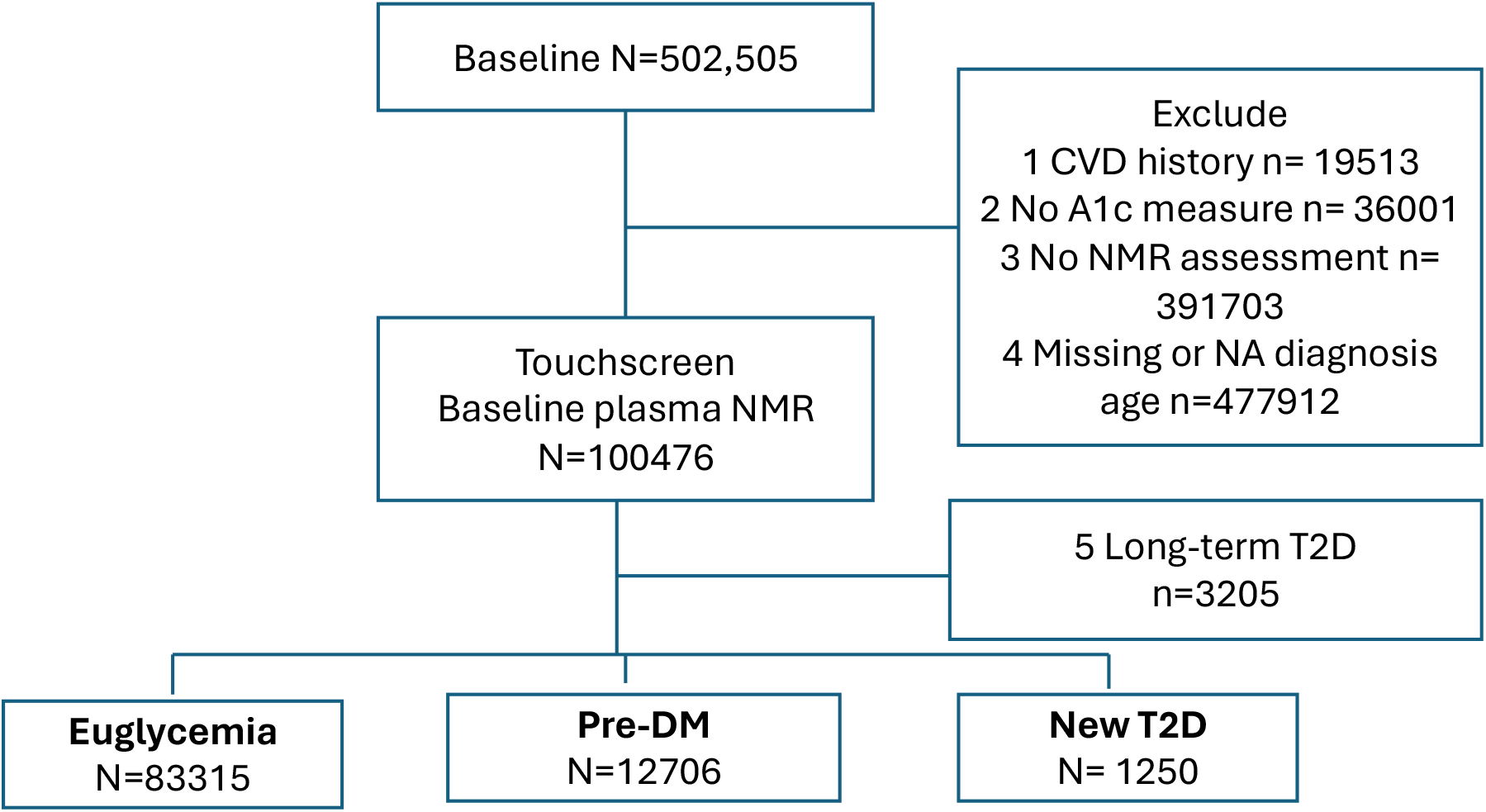
Sample Selection.

**Suppl Fig 2.**
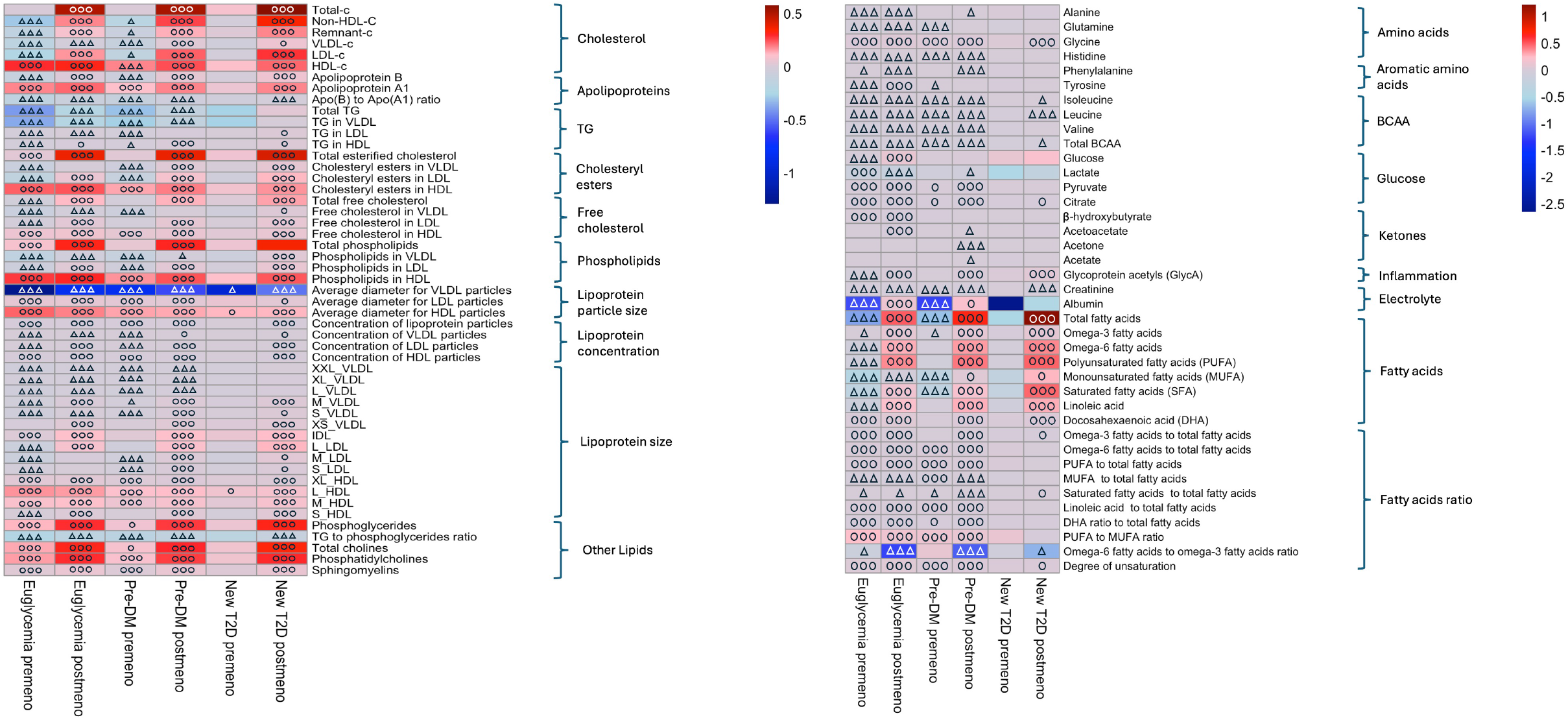
Heatmap of Sex Differences in Metabolites by Menopausal Status and Glycemic Status. Positive adjusted β coefficients (red) represent biomarkers that are higher in women ○○○ P<.0001; ○○ <.001; ○<.05 and negative β (blue) represent biomarkers higher in men. ΔΔΔ FDR-adjusted P<.0001;ΔΔ <.001; Δ<.05. Darker shade indicates larger coefficients (i.e., greater sex difference)

## References

1. Kautzky-Willer A, Leutner M, Harreiter J. Sex differences in type 2 diabetes. Diabetologia. 2023/06/01 2023;66(6):986–1002. doi:10.1007/s00125-023-05891-x

2. Yoshida Y, Chen Z, Baudier RL, et al. Sex Differences in the Progression of Metabolic Risk Factors in Diabetes Development. JAMA Network Open. 2022;5(7):e2222070–e2222070. doi:10.1001/jamanetworkopen.2022.22070

3. Yoshida Y, Chen Z, Fonseca VA, Mauvais-Jarvis F. Sex differences in cardiometabolic biomarkers during the pre-diabetes stage. Diabetes research and clinical practice. 2023/09/01/ 2023;203:110856. doi:10.1016/j.diabres.2023.110856

4. Julkunen H, Cichońska A, Tiainen M, et al. Atlas of plasma NMR biomarkers for health and disease in 118,461 individuals from the UK Biobank. Nature Communications. 2023/02/03 2023;14(1):604. doi:10.1038/s41467-023-36231-7

5. Bycroft C, Freeman C, Petkova D, et al. The UK Biobank resource with deep phenotyping and genomic data. Nature. Oct 2018;562(7726):203–209. doi:10.1038/s41586-018-0579-z

6. Christian Schnier KB, John Nolan, and Cathie Sudlow On behalf of UK Biobank Outcome Adjudication Group. UK Biobank Outcome Adjudication Group. Definitions of MI for UK Biobank Phase 1 Outcomes Adjudication. 2017. August 2017. https://biobank.ndph.ox.ac.uk/showcase/showcase/docs/alg_outcome_mi.pdf

7. Roscoe C, Sheridan C, Geneshka M, et al. Green Walkability and Physical Activity in UK Biobank: A Cross-Sectional Analysis of Adults in Greater London. Int J Environ Res Public Health. Apr 2 2022;19(7)doi:10.3390/ijerph19074247

8. Elliott P, Peakman TC. The UK Biobank sample handling and storage protocol for the collection, processing and archiving of human blood and urine. Int J Epidemiol. Apr 2008;37(2):234–44. doi:10.1093/ije/dym276

9. Chiesa ST, Charakida M, Georgiopoulos G, et al. Glycoprotein Acetyls: A Novel Inflammatory Biomarker of Early Cardiovascular Risk in the Young. Journal of the American Heart Association. 2022;11(4):e024380. doi:doi:10.1161/JAHA.121.024380

10. Du T, Fernandez C, Barshop R, et al. Sex Differences in Cardiovascular Risk Profile From Childhood to Midlife Between Individuals Who Did and Did Not Develop Diabetes at Follow-up: The Bogalusa Heart Study. Diabetes Care. 2019;42(4):635–643. doi:10.2337/dc18-2029

11. Holmes MV, Millwood IY, Kartsonaki C, et al. Lipids, Lipoproteins, and Metabolites and Risk of Myocardial Infarction and Stroke. J Am Coll Cardiol. Feb 13 2018;71(6):620–632. doi:10.1016/j.jacc.2017.12.006

12. Walden CE, Knopp RH, Wahl PW, Beach KW, Strandness E. Sex Differences in the Effect of Diabetes Mellitus on Lipoprotein Triglyceride and Cholesterol Concentrations. New England Journal of Medicine. 1984;311(15):953–959. doi:10.1056/nejm198410113111505

13. Garvey WT, Kwon S, Zheng D, et al. Effects of Insulin Resistance and Type 2 Diabetes on Lipoprotein Subclass Particle Size and Concentration Determined by Nuclear Magnetic Resonance. Diabetes. 2003;52(2):453–462. doi:10.2337/diabetes.52.2.453

14. Karalis DG, Wild RA, Maki KC, et al. Gender differences in side effects and attitudes regarding statin use in the Understanding Statin Use in America and Gaps in Patient Education (USAGE) study. Journal of Clinical Lipidology. 2016/07/01/ 2016;10(4):833–841. doi:10.1016/j.jacl.2016.02.016

15. Arques S. Human serum albumin in cardiovascular diseases. European Journal of Internal Medicine. 2018/06/01/ 2018;52:8–12. doi:10.1016/j.ejim.2018.04.014

16. Weaving G, Batstone GF, Jones RG. Age and sex variation in serum albumin concentration: an observational study. Ann Clin Biochem. Jan 2016;53(Pt 1):106-11. doi:10.1177/0004563215593561

17. Melsom T, Norvik JV, Enoksen IT, et al. Sex Differences in Age-Related Loss of Kidney Function. Journal of the American Society of Nephrology. 2022;33(10):1891–1902. doi:10.1681/asn.2022030323

18. Sowers JR. Diabetes Mellitus and Cardiovascular Disease in Women. Archives of Internal Medicine. 1998;158(6):617–621. doi:10.1001/archinte.158.6.617

19. Kaseta JR, Skafar DF, Ram JL, Jacober SJ, Sowers JR. Cardiovascular Disease in the Diabetic Woman. The Journal of Clinical Endocrinology & Metabolism. 1999;84(6):1835–1838. doi:10.1210/jcem.84.6.5735

20. Dantas AP, Fortes ZB, de Carvalho MH. Vascular disease in diabetic women: Why do they miss the female protection? Exp Diabetes Res. 2012;2012:570598. doi:10.1155/2012/570598

21. Hirano T. Pathophysiology of Diabetic Dyslipidemia. J Atheroscler Thromb. Sep 1 2018;25(9):771– 782. doi:10.5551/jat.RV17023

22. Castelli WP. The triglyceride issue: a view from Framingham. Am Heart J. Aug 1986;112(2):432–7. doi:10.1016/0002-8703(86)90296-6

23. Hokanson JE, Austin MA. Plasma triglyceride level is a risk factor for cardiovascular disease independent of high-density lipoprotein cholesterol level: a meta-analysis of population-based prospective studies. J Cardiovasc Risk. Apr 1996;3(2):213–9.

24. Balling M, Afzal S, Smith GD, et al. Elevated LDL Triglycerides and Atherosclerotic Risk. Journal of the American College of Cardiology. 2023;81(2):136–152. doi:doi:10.1016/j.jacc.2022.10.019

25. Alexopoulos AS, Qamar A, Hutchins K, Crowley MJ, Batch BC, Guyton JR. Triglycerides: Emerging Targets in Diabetes Care? Review of Moderate Hypertriglyceridemia in Diabetes. Curr Diab Rep. Feb 26 2019;19(4):13. doi:10.1007/s11892-019-1136-3

26. Castañer O, Pintó X, Subirana I, et al. Remnant Cholesterol, Not LDL Cholesterol, Is Associated With Incident Cardiovascular Disease. J Am Coll Cardiol. Dec 8 2020;76(23):2712–2724. doi:10.1016/j.jacc.2020.10.008

27. Lu Y, Ding Q, Xu X, et al. Sex Differences in Omega-3 and -6 Fatty Acids and Health Status Among Young Adults With Acute Myocardial Infarction: Results From the VIRGO Study. J Am Heart Assoc. May 30 2018;7(11)doi:10.1161/jaha.117.008189

28. Puchalska P, Crawford PA. Multi-dimensional Roles of Ketone Bodies in Fuel Metabolism, Signaling, and Therapeutics. Cell Metab. Feb 7 2017;25(2):262–284. doi:10.1016/j.cmet.2016.12.022

29. Bedi KC, Jr., Snyder NW, Brandimarto J, et al. Evidence for Intramyocardial Disruption of Lipid Metabolism and Increased Myocardial Ketone Utilization in Advanced Human Heart Failure. Circulation. Feb 23 2016;133(8):706–16. doi:10.1161/circulationaha.115.017545

30. Gormsen LC, Svart M, Thomsen HH, et al. Ketone Body Infusion With 3-Hydroxybutyrate Reduces Myocardial Glucose Uptake and Increases Blood Flow in Humans: A Positron Emission Tomography Study. J Am Heart Assoc. Feb 27 2017;6(3)doi:10.1161/jaha.116.005066

31. Shemesh E, Chevli PA, Islam T, et al. Circulating ketone bodies and cardiovascular outcomes: the MESA study. European Heart Journal. 2023;44(18):1636–1646. doi:10.1093/eurheartj/ehad087

32. Cho IY, Chang Y, Sung E, et al. Fasting ketonuria is inversely associated with coronary artery calcification in non-diabetic individuals. Atherosclerosis. 2022/05/01/ 2022;348:1–7. doi:10.1016/j.atherosclerosis.2022.03.018

