## Supplemental Table 1 for "Sex differences in circulating metabolites across glycemic status and risk of coronary heart disease"

**Supplemental Table 1. Adjusted** **β Coefficients (95% CI) for Sex Differences of and Adjusted Mean (95% CI) of Metabolites Across Glycemic Status**

|  | **Sex Difference** | | | **Euglycemia** | | **Pre-DM** | | **New T2D** | |
| --- | --- | --- | --- | --- | --- | --- | --- | --- | --- |
|  | **Euglycemia** | **Pre-DM** | **New T2D** | **Women** | **Men** | **Women** | **Men** | **Women** | **Men** |
| **Cholesterol (mmol/l)** | **β coefficient** | **β coefficient** | **β coefficient** | **Adjusted mean** | **Adjusted mean** | **Adjusted mean** | **Adjusted mean** | **Adjusted mean** | **Adjusted mean** |
| Total-c | 0.35 (0.34, 0.36) ^***^ | 0.47 (0.43, 0.51) ^***^ | 0.51 (0.39, 0.62) ^***^ | 4.45, (4.42, 4.47) | 4.10, (4.08, 4.12) | 4.53, (4.49, 4.58) | 4.06, (4.03, 4.10) | 4.23, (4.10, 4.36) | 3.72, (3.61, 3.84) |
| Non-HDL-c | 0.05 (0.04, 0.06) ^***^ | 0.23 (0.19, 0.26) ^***^ | 0.29 (0.19, 0.39) ^***^ | 3.01, (2.99, 3.04) | 2.98, (2.96, 3.00) | 3.19, (3.15, 3.23) | 2.96, (2.93, 3.00) | 2.97, (2.86, 3.09) | 2.68, (2.59, 2.78) |
| Remnant-c | 0.03 (0.02, 0.03) ^***^ | 0.12 (0.1, 0.13) ^***^ | 0.16 (0.11, 0.21) ^***^ | 1.42, (1.41, 1.43) | 1.40, (1.39, 1.41) | 1.51, (1.49, 1.53) | 1.39, (1.37, 1.41) | 1.42, (1.36, 1.48) | 1.26, (1.22, 1.31) |
| VLDL-c | -0.06 (-0.06, -0.05) ^***^ | 0.01 (0, 0.02) ^**^ | 0.05 (0.02, 0.08) ^**^ | 0.62, (0.61, 0.62) | 0.68, (0.67, 0.69) | 0.69, (0.68, 0.71) | 0.68, (0.67, 0.69) | 0.68, (0.64, 0.72) | 0.63, (0.60, 0.66) |
| LDL-c | 0.06 (0.04, 0.06) ^***^ | 0.11 (0.09, 0.13) ^***^ | 0.13 (0.08, 0.19) ^***^ | 1.60, (1.58, 1.61) | 1.58, (1.57, 1.59) | 1.68, (1.66, 1.70) | 1.57, (1.55, 1.59) | 1.56, (1.49, 1.62) | 1.42, (1.37, 1.48) |
| HDL-c | 0.30 (0.29, 0.31) ^***^ | 0.24 (0.23, 0.26) ^***^ | 0.22 (0.18, 0.25) ^***^ | 1.43, (1.42, 1.44) | 1.12, (1.12, 1.13) | 1.34, (1.33, 1.36) | 1.10, (1.09, 1.11) | 1.26, (1.22, 1.30) | 1.04, (1.01, 1.07) |
| **Apolipoproteins (g/l)** | **Eugly β** | **PreDM β** | **New T2D β** | **Eugly W a-mean** | **Eugly M a-mean** | **PreDM W a-mean** | **PreDM M a-mean** | **T2D W a-mean** | **T2D M a-mean** |
| Apolipoprotein B | -0.01 (-0.02, -0.01) *** | 0.01 (0.001, 0.02) ** | 0.05 (0.03, 0.08) *** | 0.77, (0.76, 0.77) | 0.78, (0.78, 0.79) | 0.82, (0.81, 0.83) | 0.78, (0.78, 0.79) | 0.78, (0.75, 0.80) | 0.73, (0.70, 0.75) |
| Apolipoprotein A1 | 0.21 (0.20, 0.21) *** | 0.21 (0.19, 0.22) *** | 0.19 (0.16, 0.22) *** | 1.54, (1.53, 1.54) | 1.33, (1.32, 1.33) | 1.49, (1.48, 1.50) | 1.31, (1.30, 1.32) | 1.44, (1.41, 1.48) | 1.27, (1.24, 1.29) |
| Apo(B) to apo(A1) ratio | -0.09 (-0.10, -0.09) *** | -0.07 (-0.08, -0.06) *** | -0.03 (-0.05, -0.01) ** | 0.51, (0.50, 0.51) | 0.60, (0.60, 0.61) | 0.56, (0.55, 0.57) | 0.61, (0.60, 0.62) | 0.55, (0.53, 0.58) | 0.58, (0.56, 0.60) |
| **Triglycerides (mmol/l)** | **Eugly β** | **PreDM** **β** | **New T2D** **β** | **Eugly W a-mean** | **Eugly M a-mean** | **PreDM W a-mean** | **PreDM M a-mean** | **T2D W a-mean** | **T2D M a-mean** |
| Total TG | -0.25 (-0.26, -0.24) ^***^ | -0.16 (-0.19, -0.12) ^***^ | -0.01 (-0.1, 0.08) | 1.14, (1.13, 1.16) | 1.40, (1.39, 1.41) | 1.30, (1.28, 1.33) | 1.44, (1.41, 1.47) | 1.45, (1.35, 1.55) | 1.47, (1.38, 1.55) |
| TG in VLDL | -0.24 (-0.25, -0.23) ^***^ | -0.17 (-0.20-0.14) ^***^ | -0.04 (-0.11, 0.03) | 0.77, (0.75, 0.78) | 1.01, (1.00, 1.03) | 0.90, (0.88, 0.93) | 1.04, (1.02, 1.07) | 1.02, (0.94, 1.11) | 1.07, (0.99, 1.14) |
| TG in LDL | -5.98e-03 (-6.63e-03, -5.33e-03) ^***^ | -3.26e-04 (-2.87e-03, 2.22e-03) ^*^ | 0.01 (0, 0.02) ^**^ | 1.39e-01, (1.38e-01, 1.40e-01) | 0.15, (0.14, 0.15) | 0.15, (0.15, 0.15) | 0.15, (0.15, 0.15) | 0.16, (0.15, 0.17) | 0.15, (0.14, 0.16) |
| TG in HDL | -1.57e-03 (-2.32e-03, -8.13e-04) ^***^ | 0.01 (0, 0.01) ^***^ | 0.01 (0.01, 0.02) ^**^ | 1.42e-01, (1.40e-01, 1.43e-01) | 0.14, (0.14, 0.14) | 0.15, (0.15, 0.15) | 0.15, (0.14, 0.15) | 0.16, (0.15, 0.17) | 0.15, (0.14, 0.15) |
| **Cholesteryl esters (mmol/l)** | **Eugly β** | **PreDM** **β** | **New T2D** **β** | **Eugly W a-mean** | **Eugly M a-mean** | **PreDM W a-mean** | **PreDM M a-mean** | **T2D W a-mean** | **T2D M a-mean** |
| Total esterified cholesterol | 0.28 (0.27, 0.29) ^***^ | 0.33 (0.29, 0.36) ^***^ | 0.38 (0.30, 0.46) ^***^ | 3.25, (3.23, 3.26) | 2.98, (2.96, 2.99) | 3.30, (3.27, 3.33) | 2.95, (2.92, 2.97) | 3.07, (2.98, 3.17) | 2.70, (2.62, 2.78) |
| Cholesteryl esters in VLDL | -0.02 (-0.02, -0.02) ^***^ | 0 (-0.01, 0.01) | 0.04 (0.02, 0.05) ^***^ | 3.71e-01, (3.68e-01, 3.75e-01) | 0.40, (0.39, 0.40) | 0.41, (0.41, 0.42) | 0.39, (0.39, 0.40) | 0.39, (0.37, 0.41) | 0.36, (0.34, 0.38) |
| Cholesteryl esters in LDL | 3.72e-03 (-1.24e-03, 8.69e-03) | 0.04 (0.02, 0.05) ^**^ | 0.10 (0.06, 0.14) ^***^ | 1.17, (1.16, 1.17) | 1.17, (1.16, 1.18) | 1.23, (1.22, 1.25) | 1.16, (1.15, 1.18) | 1.15, (1.10, 1.20) | 1.06, (1.02, 1.10) |
| Cholesteryl esters in HDL | 0.23 (0.23, 0.24) ^***^ | 0.22 (0.2, 0.23) ^***^ | 0.17 (0.14, 0.20) ^***^ | 1.12, (1.11, 1.12) | 0.88, (0.87, 0.88) | 1.04, (1.03, 1.06) | 0.86, (0.85, 0.87) | 0.98, (0.94, 1.01) | 0.81, (0.78, 0.84) |
| **Free cholesterol (mmol/l)** | **Eugly β** | **PreDM** **β** | **New T2D** **β** | **Eugly W a-mean** | **Eugly M a-mean** | **PreDM W a-mean** | **PreDM M a-mean** | **T2D W a-mean** | **T2D M a-mean** |
| Total free cholesterol | 0.08 (0.07, 0.08) ^***^ | 0.10 (0.09, 0.12) ^***^ | 0.13 (0.1, 0.16) ^***^ | 1.20, (1.19, 1.20) | 1.13, (1.12, 1.13) | 1.24, (1.22, 1.25) | 1.12, (1.11, 1.13) | 1.16, (1.12, 1.19) | 1.03, (0.99, 1.06) |
| Cholesterol in VLDL | -0.04 (-0.04, -0.03) ^***^ | -0.02 (-0.02, -0.01) ^***^ | 0.01 (0, 0.03) ^*^ | 0.25, (0.24, 0.25) | 0.28, (0.28, 0.29) | 0.28, (0.28, 0.29) | 0.29, (0.28, 0.29) | 0.29, (0.27, 0.30) | 0.27, (0.26, 0.29) |
| Cholesterol in LDL | 0.02 (0.02, 0.02) ^***^ | 0.03 (0.02, 0.03) ^***^ | 0.04 (0.02, 0.05) ^***^ | 4.31e-01, (4.28e-01, 4.34e-01) | 0.41, (0.41, 0.42) | 0.45, (0.44, 0.45) | 0.41, (0.40, 0.42) | 0.40, (0.39, 0.42) | 0.37, (0.35, 0.38) |
| Cholesterol in HDL | 0.07 (0.07, 0.07) ^***^ | 0.07 (0.06, 0.07) ^***^ | 0.05 (0.05, 0.06) ^***^ | 0.32, (0.31, 0.32) | 0.25, (0.24, 0.25) | 0.30, (0.30, 0.30) | 0.24, (0.24, 0.25) | 0.28, (0.27, 0.29) | 0.23, (0.22, 0.24) |
| **Phospholipids (mmol/l)** | **Eugly β** | **PreDM** **β** | **New T2D** **β** | **Eugly W a-mean** | **Eugly M a-mean** | **PreDM W a-mean** | **PreDM M a-mean** | **T2D W a-mean** | **T2D M a-mean** |
| Total phospholipids | 0.25 (0.24, 0.26) ^***^ | 0.30 (0.27, 0.33) ^***^ | 0.34 (0.28, 0.40) ^***^ | 2.91, (2.90, 2.92) | 2.66, (2.65, 2.67) | 2.94, (2.91, 2.96) | 2.65, (2.63, 2.67) | 2.85, (2.77, 2.92) | 2.52, (2.46, 2.58) |
| Phospholipids in VLDL | -0.07 (-0.07, -0.06) ^***^ | -0.04 (-0.05, -0.02) ^***^ | 0.02 (-0.01, 0.04) | 0.40, (0.40, 0.41) | 0.47, (0.47, 0.48) | 0.46, (0.45, 0.47) | 0.48, (0.47, 0.49) | 0.48, (0.45, 0.51) | 0.46, (0.44, 0.49) |
| Phospholipids in LDL | 1.06e-03 (-1.10e-03, 3.22e-03) | 0.01 (0.01, 0.02) ^**^ | 0.04 (0.02, 0.06) ^***^ | 5.59e-01, (5.56e-01, 5.63e-01) | 0.56, (0.56, 0.56) | 0.59, (0.58, 0.60) | 0.56, (0.55, 0.57) | 0.55, (0.53, 0.57) | 0.51, (0.50, 0.53) |
| Phospholipids in HDL | 2.90e-01 (2.85e-01, 2.94e-01) ^***^ | 0.29 (0.27, 0.31) ^***^ | 0.25 (0.21, 0.28) ^***^ | 1.67, (1.67, 1.68) | 1.38, (1.37, 1.38) | 1.60, (1.59, 1.61) | 1.36, (1.35, 1.37) | 1.55, (1.51, 1.60) | 1.31, (1.28, 1.35) |
| **Lipoprotein particle diameter (nm)** | **Eugly β** | **PreDM** **β** | **New T2D** **β** | **Eugly W a-mean** | **Eugly M a-mean** | **PreDM W a-mean** | **PreDM M a-mean** | **T2D W a-mean** | **T2D M a-mean** |
| Average diameter for VLDL particles | -0.94(-0.96, -0.92) ^***^ | -0.77 (-0.85, -0.7) ^***^ | -0.51 (-0.69, -0.33) ^***^ | 38.14, (38.11, 38.17) | 39.11, (39.09, 39.14) | 38.53, (38.47, 38.58) | 39.16, (39.10, 39.21) | 38.80, (38.59, 39.02) | 39.33, (39.14, 39.51) |
| Average diameter for LDL particles | 3.82e-02 (3.68e-02, 3.96e-02) ^***^ | 0.04 (0.03, 0.04) ^***^ | 0.03 (0.02, 0.04) ^***^ | 2.39e+01, (2.39e+01, 2.39e+01) | 23.89, (23.89, 23.89) | 23.92, (23.92, 23.93) | 23.89, (23.88, 23.89) | 23.90, (23.88, 23.91) | 23.86, (23.85, 23.88) |
| Average diameter for HDL particles | 1.93e-01 (1.90e-01, 1.96e-01) ^***^ | 0.18 (0.17, 0.19) ^***^ | 0.12 (0.10, 0.14) ^***^ | 9.71, (9.71, 9.72) | 9.51, (9.51, 9.52) | 9.65, (9.64, 9.66) | 9.51, (9.50, 9.52) | 9.60, (9.58, 9.63) | 9.48, (9.46, 9.51) |
| **Concentration of lipoprotein particles (mmol/l)** | 1.74e-03 (1.70e-03, 1.77e-03) ^***^ | 1.78e-03 (1.64e-03, 1.92e-03) ^***^ | 1.92e-03 (1.59e-03, 2.25e-03) ^***^ | 1.73e-02, (1.73e-02, 1.74e-02) | 0.02, (0.02, 0.02) | 0.02, (0.02, 0.02) | 0.02, (0.02, 0.02) | 0.02, (0.02, 0.02) | 0.01, (0.01, 0.02) |
| Concentration of VLDL particles | -1.06e-05 (-1.13e-05, -9.92e-06) ^***^ | -3.25e-06 (-5.90e-06, -6.02e-07) | 8.46e-06 (2.67e-06, 1.43e-05) ^**^ | 1.28e-04, (1.26e-04, 1.29e-04) | 1.39e-04, (1.38e-04, 1.40e-04) | 1.42e-04, (1.39e-04, 1.44e-04) | 1.41e-04, (1.39e-04, 1.43e-04) | 1.42e-04, (1.36e-04, 1.49e-04) | 1.34e-04, (1.29e-04, 1.40e-04) |
| Concentration of LDL particles | -2.40e-05 (-2.83e-05, -1.96e-05) ^***^ | 7.44e-06 (-8.74e-06, 2.36e-05) | 6.11e-05 (2.67e-05, 9.54e-05) ^**^ | 1.08e-03, (1.08e-03, 1.09e-03) | 1.11e-03, (1.11e-03, 1.12e-03) | 1.15e-03, (1.14e-03, 1.17e-03) | 1.11e-03, (1.10e-03, 1.13e-03) | 1.09e-03, (1.05e-03, 1.13e-03) | 1.03e-03, (9.97e-04, 1.06e-03) |
| Concentration of HDL particles | 1.76e-03 (1.72e-03, 1.80e-03) ^***^ | 1.75e-03 (1.62e-03, 1.89e-03) ^***^ | 1.82e-03 (1.50e-03, 2.14e-03) ^***^ | 1.58e-02, (1.58e-02, 1.59e-02) | 0.01, (0.01, 0.01) | 0.02, (0.02, 0.02) | 0.01, (0.01, 0.01) | 0.02, (0.01, 0.02) | 1.33e-02, (1.30e-02, 1.36e-02) |
| **Lipoprotein size** | **Eugly β** | **PreDM** **β** | **New T2D** **β** | **Eugly W a-mean** | **Eugly M a-mean** | **PreDM W a-mean** | **PreDM M a-mean** | **T2D W a-mean** | **T2D M a-mean** |
| XXL_VLDL | -0.02 (-0.02, -0.02) ^***^ | -0.02 (-0.02, -0.01) ^***^ | -3.45e-03 (-0.01, 0) | 4.37e-02, (4.26e-02, 4.48e-02) | 0.07, (0.07, 0.07) | 0.06, (0.05, 0.06) | 0.07, (0.07, 0.07) | 0.07, (0.06, 0.07) | 0.07, (0.07, 0.08) |
| XL_VLDL | -0.01 (-0.02, -0.01) ^***^ | -0.01 (-0.01, -0.01) ^***^ | -1.25e-03 (-0.01, 0) | 4.18e-02, (4.11e-02, 4.26e-02) | 0.06, (0.06, 0.06) | 0.05, (0.05, 0.05) | 0.06, (0.06, 0.06) | 0.06, (0.05, 0.06) | 0.06, (0.05, 0.06) |
| L_VLDL | -0.02 (-0.02, -0.02) ^***^ | -0.02 (-0.02, -0.01) ^***^ | -1.57e-03 (-0.01, 0) | 7.77e-02, (7.64e-02, 7.89e-02) | 0.10, (0.10, 0.10) | 0.09, (0.09, 0.10) | 0.10, (0.10, 0.11) | 0.10, (0.09, 0.11) | 0.10, (0.10, 0.11) |
| M_VLDL | -7.35e-04 (-1.74e-03, 2.67e-04) | 0.01 (0, 0.01) ^**^ | 0.02 (0.01, 0.03) ^***^ | 0.14, (0.14, 0.15) | 0.15, (0.15, 0.15) | 0.16, (0.16, 0.16) | 0.14, (0.14, 0.15) | 0.14, (0.13, 0.15) | 0.12, (0.12, 0.13) |
| S_VLDL | -0.01 (-0.01, -0.01) ^***^ | -3.42e-03 (-0.01, 0) ^**^ | 0.01 (0, 0.02) ^**^ | 0.14, (0.13, 0.14) | 0.15, (0.15, 0.15) | 0.15, (0.15, 0.15) | 0.15, (0.15, 0.15) | 0.15, (0.14, 0.15) | 0.14, (0.13, 0.14) |
| XS_VLDL | 0.02 (0.02, 0.02) ^***^ | 0.02 (0.02, 0.02) ^***^ | 0.03 (0.02, 0.03) ^***^ | 0.17, (0.17, 0.18) | 0.16, (0.16, 0.16) | 0.18, (0.18, 0.18) | 0.16, (0.16, 0.16) | 0.17, (0.16, 0.18) | 0.14, (0.14, 0.15) |
| IDL | 0.08 (0.08, 0.09) ^***^ | 0.1 (0.09, 0.11) ^***^ | 0.11 (0.09, 0.13) ^***^ | 0.80, (0.80, 0.81) | 0.72, (0.71, 0.72) | 0.82, (0.81, 0.83) | 0.71, (0.70, 0.72) | 0.74, (0.71, 0.77) | 0.63, (0.61, 0.65) |
| L_LDL | 0.04 (0.04, 0.05) ^***^ | 0.07 (0.05, 0.08) ^***^ | 0.1 (0.07, 0.14) ^***^ | 1.05, (1.05, 1.06) | 1.01, (1.01, 1.02) | 1.10, (1.08, 1.11) | 1.00, (0.99, 1.02) | 1.01, (0.97, 1.05) | 0.90, (0.87, 0.94) |
| M_LDL | -0.02 (-0.02, -0.01) ^***^ | -4.82e-03 (-1.17e-02, 2.06e-03) | 0.02 (0.01, 0.04) ^**^ | 3.80e-01, (3.78e-01, 3.83e-01) | 0.40, (0.40, 0.40) | 0.41, (0.40, 0.42) | 0.40, (0.39, 0.40) | 0.39, (0.37, 0.40) | 0.36, (0.35, 0.38) |
| S_LDL | -0.01 (-0.01, -0.01) ^***^ | -2.13e-03 (-4.70e-03, 4.45e-04) | 0.01 (0, 0.01) | 1.63e-01, (1.62e-01, 1.64e-01) | 0.17, (0.17, 0.17) | 0.17, (0.17, 0.18) | 0.17, (0.17, 0.17) | 0.16, (0.16, 0.17) | 0.16, (0.15, 0.16) |
| XL_HDL | 0.03 (0.03, 0.03) ^***^ | 0.03 (0.02, 0.03) ^***^ | 0.01 (0.01, 0.02) ^***^ | 8.83e-02, (8.74e-02, 8.91e-02) | 0.06, (0.06, 0.06) | 0.08, (0.08, 0.08) | 0.06, (0.06, 0.06) | 0.07, (0.06, 0.07) | 0.05, (0.05, 0.06) |
| L_HDL | 0.16 (0.15, 0.16) ^***^ | 0.14 (0.14, 0.15) ^***^ | 0.09 (0.07, 0.1) ^***^ | 3.50e-01, (3.46e-01, 3.54e-01) | 0.19, (0.19, 0.19) | 0.30, (0.29, 0.30) | 0.18, (0.18, 0.19) | 0.25, (0.23, 0.27) | 0.16, (0.15, 0.18) |
| M_HDL | 0.11 (0.10, 0.11) ^***^ | 0.10 (0.09, 0.11) ^***^ | 0.09 (0.08, 0.1) ^***^ | 0.54, (0.54, 0.55) | 0.44, (0.43, 0.44) | 0.52, (0.51, 0.52) | 0.43, (0.42, 0.43) | 0.49, (0.48, 0.51) | 0.41, (0.39, 0.42) |
| S_HDL | 0.01 (0.01, 0.01) ^***^ | 0.01 (0.01, 0.02) ^***^ | 0.03 (0.02, 0.04) ^***^ | 4.49e-01, (4.47e-01, 4.51e-01) | 0.44, (0.44, 0.44) | 0.45, (0.45, 0.46) | 0.43, (0.43, 0.43) | 0.45, (0.44, 0.46) | 0.42, (0.41, 0.43) |
| **Other lipids (mmol/L)** | **Eugly β** | **PreDM** **β** | **New T2D** **β** | **Eugly W a-mean** | **Eugly M a-mean** | **PreDM W a-mean** | **PreDM M a-mean** | **T2D W a-mean** | **T2D M a-mean** |
| Phosphoglycerides | 0.24 (0.24, 0.25) ^***^ | 0.28 (0.26, 0.30) ^***^ | 0.31 (0.25, 0.36) ^***^ | 2.32, (2.31, 2.33) | 2.08, (2.07, 2.08) | 2.32, (2.30, 2.34) | 2.07, (2.05, 2.08) | 2.28, (2.22, 2.34) | 1.99, (1.93, 2.04) |
| TG to phosphoglycerides ratio | -0.17 (-0.17, -0.17) ^***^ | -0.14 (-0.15, -0.13) ^***^ | -0.10 (-0.14, -0.07) ^***^ | 0.49, (0.49, 0.50) | 0.66, (0.66, 0.67) | 0.56, (0.55, 0.57) | 0.69, (0.68, 0.70) | 0.62, (0.58, 0.66) | 0.72, (0.69, 0.76) |
| Total cholines | 0.26 (0.26, 0.27) ^***^ | 0.30 (0.28, 0.32) ^***^ | 0.32 (0.26, 0.37) ^***^ | 2.59, (2.58, 2.60) | 2.33, (2.32, 2.34) | 2.59, (2.57, 2.61) | 2.31, (2.29, 2.33) | 2.52, (2.46, 2.58) | 2.22, (2.16, 2.27) |
| Phosphatidylcholines | 0.25 (0.24, 0.26) ^***^ | 0.29 (0.26, 0.31) ^***^ | 0.29 (0.24, 0.34) ^***^ | 2.14, (2.13, 2.15) | 1.89, (1.88, 1.90) | 2.13, (2.11, 2.14) | 1.87, (1.86, 1.89) | 2.08, (2.02, 2.14) | 1.80, (1.75, 1.85) |
| Sphingomyelins | 0.05 (0.04, 0.05) ^***^ | 0.05 (0.045, 0.05) ^***^ | 0.05 (0.04, 0.06) ^***^ | 0.45, (0.44, 0.45) | 0.40, (0.40, 0.40) | 0.45, (0.44, 0.45) | 0.40, (0.40, 0.40) | 0.43, (0.42, 0.44) | 0.37, (0.37, 0.38) |
| **Amino acids (mmol/L)** | **Eugly β** | **PreDM** **β** | **New T2D** **β** | **Eugly W a-mean** | **Eugly M a-mean** | **PreDM W a-mean** | **PreDM M a-mean** | **T2D W a-mean** | **T2D M a-mean** |
| Alanine | -0.01 (-0.02, -0.01) ^***^ | -0.01 (-0.01, 0) ^**^ | 5.46e-05 (-0.01, 0.01) | 2.90e-01, (2.88e-01, 2.92e-01) | 0.31, (0.30, 0.31) | 0.30, (0.30, 0.30) | 0.31, (0.30, 0.31) | 0.34, (0.33, 0.36) | 0.34, (0.33, 0.36) |
| Glutamine | -0.02 (-0.02, -0.01) ^***^ | -0.01 (-0.02, -0.01) ^***^ | -0.01 (-0.02, 0) | 5.12e-01, (5.10e-01, 5.14e-01) | 0.53, (0.53, 0.53) | 0.52, (0.51, 0.52) | 0.52, (0.52, 0.52) | 0.50, (0.49, 0.52) | 0.51, (0.50, 0.52) |
| Glycine | 4.23e-02 (4.13e-02, 4.33e-02) ^***^ | 0.04 (0.03, 0.04) ^***^ | 0.02 (0.01, 0.03) ^***^ | 0.17, (0.17, 0.18) | 0.13, (0.13, 0.13) | 0.16, (0.16, 0.17) | 0.13, (0.12, 0.13) | 0.15, (0.14, 0.15) | 0.13, (0.12, 0.13) |
| Histidine | -2.23e-03 (-2.40e-03, -2.06e-03) ^***^ | -1.63e-03 (-2.26e-03, -1.01e-03) ^***^ | -1.62e-03 (-3.07e-03, -1.56e-04) ^**^ | 6.28e-02, (6.25e-02, 6.31e-02) | 0.06, (0.06, 0.07) | 0.06, (0.06, 0.06) | 0.06, (0.06, 0.06) | 0.06, (0.06, 0.06) | 0.06, (0.06, 0.07) |
| **Aromatic amino acides (mmol/L)** | **Eugly β** | **PreDM** **β** | **New T2D** **β** | **Eugly W a-mean** | **Eugly M a-mean** | **PreDM W a-mean** | **PreDM M a-mean** | **T2D W a-mean** | **T2D M a-mean** |
| Phenylalanine | -1.17e-03 (-1.34e-03, -1.01e-03) ^***^ | -3.31e-04 (-1.01e-03, 3.46e-04) | -6.01e-04 (-2.06e-03, 8.53e-04) | 0.04, (0.04, 0.05) | 0.05, (0.05, 0.05) | 0.05, (0.04, 0.05) | 0.05, (0.05, 0.05) | 0.05, (0.04, 0.05) | 0.05, (0.05, 0.05) |
| Tyrosine | -1.47e-05 (-2.38e-04, 2.09e-04) | 5.10e-04 (-3.68e-04, 1.39e-03) | -2.27e-04 (-2.36e-03, 1.90e-03) | 6.33e-02, (6.29e-02, 6.37e-02) | 0.06, (0.06, 0.06) | 0.06, (0.06, 0.07) | 0.07, (0.06, 0.07) | 0.07, (0.07, 0.07) | 0.07, (0.07, 0.07) |
| **Branched-chain amino acids (BCAA) (mmol/L**) | **Eugly β** | **PreDM** **β** | **New T2D** **β** | **Eugly W a-mean** | **Eugly M a-mean** | **PreDM W a-mean** | **PreDM M a-mean** | **T2D W a-mean** | **T2D M a-mean** |
| Isoleucine | -7.03e-03 (-7.31e-03, -6.75e-03) ^***^ | -0.01 (-0.01, 0) ^***^ | -4.54e-03 (-0.01, 0) ^**^ | 4.84e-02, (4.80e-02, 4.89e-02) | 0.06, (0.06, 0.06) | 0.05, (0.05, 0.05) | 0.06, (0.06, 0.06) | 0.06, (0.05, 0.06) | 0.06, (0.06, 0.06) |
| Leucine | -0.02 (-0.02, -0.01) ^***^ | -0.01 (-0.02, -0.01) ^***^ | -0.01 (-0.02, -0.01) ^***^ | 9.68e-02, (9.61e-02, 9.76e-02) | 0.11, (0.11, 0.11) | 0.10, (0.10, 0.10) | 0.11, (0.11, 0.11) | 0.11, (0.10, 0.11) | 0.12, (0.11, 0.12) |
| Valine | -1.93e-02 (-1.99e-02, -1.86e-02) ^***^ | -0.01 (-0.02, -0.01) ^***^ | -0.01 (-0.02, 0) ^**^ | 1.99e-01, (1.98e-01, 2.00e-01) | 0.22, (0.22, 0.22) | 0.21, (0.20, 0.21) | 0.22, (0.22, 0.22) | 0.22, (0.21, 0.23) | 0.23, (0.23, 0.24) |
| Total BCAA | -4.16e-02 (-4.30e-02, -4.03e-02) ^***^ | -0.03 (-0.04, -0.03) ^***^ | -0.03 (-0.04, -0.01) ^***^ | 0.34, (0.34, 0.35) | 0.39, (0.38, 0.39) | 0.36, (0.35, 0.36) | 0.39, (0.38, 0.39) | 0.38, (0.37, 0.40) | 0.41, (0.40, 0.42) |
| **Glucose metabolism (mmol/l )** | **Eugly β** | **PreDM** **β** | **New T2D** **β** | **Eugly W a-mean** | **Eugly M a-mean** | **PreDM W a-mean** | **PreDM M a-mean** | **T2D W a-mean** | **T2D M a-mean** |
| Glucose | 0.06 (0.05, 0.07) ^***^ | 0.09 (0.05, 0.12) ^***^ | 0.33 (0, 0.67) ^*^ | 3.48, (3.46, 3.50) | 3.41, (3.39, 3.43) | 3.69, (3.65, 3.73) | 3.67, (3.63, 3.71) | 4.77, (4.40, 5.14) | 4.60, (4.29, 4.91) |
| Lactate | -0.15 (-0.17, -0.13) ^***^ | -0.13 (-0.2, -0.06) ^**^ | -0.23 (-0.41, -0.06) ^**^ | 3.75, (3.72, 3.77) | 3.90, (3.88, 3.93) | 3.81, (3.75, 3.86) | 3.87, (3.82, 3.92) | 4.10, (3.89, 4.30) | 4.37, (4.20, 4.54) |
| Pyruvate | 5.13e-03 (4.65e-03, 5.61e-03) ^***^ | 7.32e-03 (5.48e-03, 9.16e-03) ^***^ | 4.85e-03 (5.77e-04, 9.12e-03) ^**^ | 8.21e-02, (8.13e-02, 8.29e-02) | 0.08, (0.08, 0.08) | 0.08, (0.08, 0.08) | 0.08, (0.08, 0.08) | 0.09, (0.08, 0.09) | 0.08, (0.08, 0.09) |
| Citrate | 2.69e-03 (2.50e-03, 2.88e-03) ^***^ | 3.03e-03 (2.31e-03, 3.75e-03) ^***^ | 2.86e-03 (1.03e-03, 4.70e-03) ^**^ | 6.24e-02, (6.21e-02, 6.28e-02) | 0.06, (0.06, 0.06) | 0.06, (0.06, 0.06) | 0.06, (0.06, 0.06) | 0.07, (0.06, 0.07) | 0.06, (0.06, 0.07) |
| **Ketones (mmol/l)** | **Eugly β** | **PreDM** **β** | **New T2D** **β** | **Eugly W a-mean** | **Eugly M a-mean** | **PreDM W a-mean** | **PreDM M a-mean** | **T2D W a-mean** | **T2D M a-mean** |
| β-hydroxybutyrate | 4.41e-03 (3.41e-03, 5.41e-03) ^***^ | -4.15e-03 (-6.63e-03, -1.66e-03) ^**^ | 4.24e-03 (-0.01, 0.02) | 5.67e-02, (5.51e-02, 5.84e-02) | 0.05, (0.05, 0.05) | 0.05, (0.05, 0.06) | 0.05, (0.05, 0.06) | 0.06, (0.05, 0.07) | 0.06, (0.06, 0.07) |
| Acetoacetate | 4.32e-04 (2.29e-04, 6.36e-04) ^***^ | -4.38e-04 (-9.60e-04, 8.37e-05) | 2.51e-03 (1.75e-05, 0.01) | 1.29e-02, (1.25e-02, 1.32e-02) | 0.01, (0.01, 0.01) | 0.01, (0.01, 0.01) | 0.01, (0.01, 0.01) | 0.01, (0.01, 0.02) | 0.01, (0.01, 0.01) |
| Acetone | -5.85e-05 (-1.51e-04, 3.37e-05) | -3.14e-04 (-5.86e-04, -4.31e-05) ^**^ | 6.59e-04 (-5.55e-04, 1.87e-03) | 1.41e-02, (1.39e-02, 1.42e-02) | 0.01, (0.01, 0.01) | 0.01, (0.01, 0.01) | 0.01, (0.01, 0.01) | 0.01, (0.01, 0.01) | 0.01, (0.01, 0.02) |
| Acetate | -1.18e-04 (-3.00e-04, 6.40e-05) | -7.32e-04 (-1.62e-03, 1.58e-04) | -1.44e-03 (-4.06e-03, 1.18e-03) | 0.01, (0.01, 0.02) | 0.01, (0.01, 0.02) | 0.01, (0.01, 0.01) | 0.01, (0.01, 0.02) | 0.01, (0.01, 0.02) | 0.02, (0.01, 0.02) |
| **Inflammation (mmol/l)** | **Eugly β** | **PreDM** **β** | **New T2D** **β** | **Eugly W a-mean** | **Eugly M a-mean** | **PreDM W a-mean** | **PreDM M a-mean** | **T2D W a-mean** | **T2D M a-mean** |
| Glycoprotein acetyls (GlycA) | -0.01 (-0.01, 0) ^***^ | 0.01 (0, 0.02) ^**^ | 0.06 (0.04, 0.07) ^***^ | 7.88e-01, (7.85e-01, 7.91e-01) | 0.80, (0.79, 0.80) | 0.84, (0.83, 0.84) | 0.82, (0.81, 0.82) | 0.87, (0.85, 0.88) | 0.81, (0.80, 0.83) |
| **Electrolytes (mmol/l )** | **Eugly β** | **PreDM** **β** | **New T2D** **β** | **Eugly W a-mean** | **Eugly M a-mean** | **PreDM W a-mean** | **PreDM M a-mean** | **T2D W a-mean** | **T2D M a-mean** |
| Creatinine | -0.01 (-0.01, -0.01) ^***^ | -0.01 (-0.01, -0.01) ^***^ | -0.01 (-0.01, -0.01) ^***^ | 6.05e-02, (6.02e-02, 6.08e-02) | 0.07, (0.07, 0.07) | 0.06, (0.06, 0.06) | 0.07, (0.07, 0.08) | 0.06, (0.06, 0.06) | 0.07, (0.07, 0.07) |
| Albumin | -0.29 (-0.35, -0.24) ^***^ | -0.23 (-0.43, -0.03) ^**^ | -0.74 (-1.21, -0.26) ^**^ | 38.70, (38.61, 38.78) | 39.01, (38.93, 39.10) | 38.21, (38.04, 38.37) | 38.20, (38.05, 38.36) | 37.97, (37.41, 38.54) | 38.83, (38.36, 39.31) |
| **Fatty acids (mmol/l)** | **Eugly β** | **PreDM** **β** | **New T2D** **β** | **Eugly W a-mean** | **Eugly M a-mean** | **PreDM W a-mean** | **PreDM M a-mean** | **T2D W a-mean** | **T2D M a-mean** |
| Total fatty acids | 0.14 (0.11, 0.18) ^***^ | 0.52 (0.37, 0.66) ^***^ | 1.01 (0.64, 1.38) ^***^ | 11.64, (11.58, 11.70) | 11.52, (11.46, 11.58) | 12.18, (12.06, 12.29) | 11.64, (11.52, 11.75) | 12.32, (11.88, 12.76) | 11.37, (11.00, 11.74) |
| Omega-3 fatty acids | 0.06 (0.06, 0.07) ^***^ | 0.09 (0.08, 0.11) ^***^ | 0.08 (0.05, 0.11) ^***^ | 0.58, (0.58, 0.59) | 0.52, (0.51, 0.52) | 0.60, (0.59, 0.61) | 0.52, (0.51, 0.53) | 0.60, (0.56, 0.63) | 0.52, (0.49, 0.55) |
| Omega-6 fatty acids | 0.18 (0.17, 0.19) ^***^ | 0.27 (0.23, 0.31) ^***^ | 0.36 (0.26, 0.45) ^***^ | 4.42, (4.41, 4.44) | 4.25, (4.23, 4.27) | 4.55, (4.52, 4.58) | 4.25, (4.22, 4.28) | 4.45, (4.34, 4.56) | 4.11, (4.02, 4.20) |
| Polyunsaturated fatty acids (PUFA) | 0.24 (0.23, 0.25) ^***^ | 0.37 (0.32, 0.41) ^***^ | 0.43 (0.32, 0.54) ^***^ | 5.01, (4.99, 5.03) | 4.77, (4.75, 4.79) | 5.15, (5.11, 5.19) | 4.77, (4.73, 4.81) | 5.05, (4.92, 5.18) | 4.63, (4.52, 4.74) |
| Monounsaturated fatty acids (MUFA) | -0.14 (-0.15, -0.12) ^***^ | -0.02 (-0.07, 0.04) | 0.19 (0.06, 0.33) ^**^ | 2.69, (2.67, 2.71) | 2.84, (2.82, 2.86) | 2.89, (2.85, 2.93) | 2.88, (2.84, 2.92) | 3.05, (2.89, 3.21) | 2.88, (2.74, 3.01) |
| Saturated fatty acids (SFA) | 0.04 (0.02, 0.05) ^***^ | 0.17 (0.11, 0.22) ^***^ | 0.38 (0.24, 0.53) ^***^ | 3.95, (3.92, 3.97) | 3.92, (3.89, 3.94) | 4.14, (4.09, 4.18) | 3.98, (3.93, 4.03) | 4.22, (4.05, 4.40) | 3.86, (3.71, 4.01) |
| Linoleic acid | 0.12 (0.11, 0.13) ^***^ | 0.21 (0.17, 0.25) ^***^ | 0.27 (0.18, 0.37) ^***^ | 3.31, (3.30, 3.33) | 3.20, (3.18, 3.22) | 3.44, (3.41, 3.47) | 3.19, (3.16, 3.22) | 3.34, (3.23, 3.44) | 3.08, (2.99, 3.17) |
| Docosahexaenoic acid (DHA) | 0.04 (0.04, 0.04) ^***^ | 0.05 (0.04, 0.05) ^***^ | 0.04 (0.02, 0.05) ^***^ | 2.60e-01, (2.58e-01, 2.62e-01) | 0.22, (0.22, 0.22) | 0.26, (0.25, 0.26) | 0.22, (0.21, 0.22) | 0.25, (0.24, 0.26) | 0.22, (0.21, 0.23) |
| **Fatty acid ratio (%)** | **Eugly β** | **PreDM** **β** | **New T2D** **β** | **Eugly W a-mean** | **Eugly M a-mean** | **PreDM W a-mean** | **PreDM M a-mean** | **T2D W a-mean** | **T2D M a-mean** |
| Omega-3 fatty acids to total fatty acids | 4.78e-03 (4.53e-03, 5.03e-03) ^***^ | 5.69e-03 (4.73e-03, 6.65e-03) ^***^ | 2.43e-03 (3.38e-04, 4.53e-03) ^**^ | 4.99e-02, (4.95e-02, 5.03e-02) | 0.04, (0.04, 0.05) | 0.05, (0.05, 0.05) | 0.04, (0.04, 0.05) | 0.05, (0.05, 0.05) | 0.05, (0.04, 0.05) |
| Omega-6 fatty acids to total fatty acids | 9.14e-03 (8.59e-03, 9.69e-03) ^***^ | 5.69e-03 (3.56e-03, 7.81e-03) ^***^ | -2.18e-03 (-7.66e-03, 3.29e-03) | 0.38, (0.38, 0.39) | 0.38, (0.37, 0.38) | 0.38, (0.38, 0.38) | 0.37, (0.37, 0.37) | 0.37, (0.36, 0.37) | 0.37, (0.36, 0.37) |
| PUFA to total fatty acids | 1.39e-02 (1.34e-02, 1.45e-02) ^***^ | 1.14e-02 (9.17e-03, 1.36e-02) ^***^ | 2.51e-04 (-0.01, 0.01) | 0.43, (0.43, 0.44) | 0.42, (0.42, 0.42) | 0.43, (0.43, 0.43) | 0.42, (0.41, 0.42) | 0.42, (0.41, 0.42) | 0.41, (0.41, 0.42) |
| MUFA to total fatty acids | -1.37e-02 (-1.41e-02, -1.33e-02) ^***^ | -1.09e-02 (-1.25e-02, -9.39e-03) ^***^ | -4.43e-03 (-8.37e-03, -4.93e-04) ^**^ | 2.28e-01, (2.27e-01, 2.28e-01) | 0.24, (0.24, 0.24) | 0.23, (0.23, 0.24) | 0.24, (0.24, 0.25) | 0.24, (0.24, 0.25) | 0.25, (0.24, 0.25) |
| Saturated fatty acids to total fatty acids | -2.50e-04 (-5.57e-04, 5.66e-05) | -4.47e-04 (-1.64e-03, 7.50e-04) | 4.18e-03 (1.19e-03, 7.17e-03) ^***^ | 3.37e-01, (3.37e-01, 3.38e-01) | 0.34, (0.34, 0.34) | 0.34, (0.34, 0.34) | 0.34, (0.34, 0.34) | 0.34, (0.34, 0.34) | 0.34, (0.33, 0.34) |
| Linoleic acid to total fatty acids | 5.48e-03 (4.98e-03, 5.99e-03) ^***^ | 4.88e-03 (2.95e-03, 6.81e-03) ^***^ | -2.55e-04 (-4.77e-03, 4.26e-03) | 2.86e-01, (2.86e-01, 2.87e-01) | 0.28, (0.28, 0.28) | 0.28, (0.28, 0.29) | 0.28, (0.28, 0.28) | 0.27, (0.27, 0.28) | 0.27, (0.27, 0.28) |
| DHA to total fatty acids | 2.98e-03 (2.87e-03, 3.09e-03) ^***^ | 2.86e-03 (2.44e-03, 3.28e-03) ^***^ | 1.09e-03 (1.37e-04, 2.03e-03) ^**^ | 2.27e-02, (2.25e-02, 2.28e-02) | 0.02, (0.02, 0.02) | 0.02, (0.02, 0.02) | 0.02, (0.02, 0.02) | 0.02, (0.02, 0.02) | 0.02, (0.02, 0.02) |
| PUFA to MUFA ratio | 0.16 (0.16, 0.17) ^***^ | 0.13 (0.11, 0.15) ^***^ | 0.02 (-0.02, 0.07) | 1.95, (1.94, 1.96) | 1.78, (1.77, 1.79) | 1.86, (1.85, 1.88) | 1.75, (1.74, 1.77) | 1.74, (1.69, 1.80) | 1.71, (1.67, 1.76) |
| Omega-6 fatty acids to omega-3 fatty acids | -0.93 (-1, -0.86) ^***^ | -1.13 (-1.35, -0.91) ^***^ | -0.63 (-1.13, -0.14) ^**^ | 8.63, (8.52, 8.74) | 9.56, (9.45, 9.67) | 8.56, (8.35, 8.77) | 9.53, (9.34, 9.73) | 8.70, (8.11, 9.30) | 9.38, (8.88, 9.88) |
| Degree of unsaturation | 0.04 (0.04, 0.04) ^***^ | 0.04 (0.04, 0.05) ^***^ | 0.02 (0.01, 0.03) ^**^ | 1.40e+00, (1.40e+00, 1.40e+00) | 1.35, (1.35, 1.36) | 1.39, (1.38, 1.39) | 1.35, (1.35, 1.35) | 1.37, (1.35, 1.38) | 1.34, (1.33, 1.35) |

FDR-adjusted P *<.1; ** <.05; ***<.0001

Fully adjusted for age, race, income, area deprivation, smoking, alcohol drinking, physical activity, BMI, and medications for hypertension, hyperlipidemia, and diabetes.
