## Supplemental Table 2 for "Sex differences in circulating metabolites across glycemic status and risk of coronary heart disease"

**Supplemental Table 2. Adjusted β Coefficients (95% CI) for Sex Differences of Metabolites Across Glycemic Status by Menopausal Status**

|  | **Postmenopausal** |  |  | **Pre-menopausal** |  |  |
| --- | --- | --- | --- | --- | --- | --- |
|  | **Euglycemia** | **Pre-DM** | **New T2D** | **Euglycemia** | **Pre-DM** | **New T2D** |
| **Cholesterol (mmol/l)** | **β coefficient** | **β coefficient** | **β coefficient** | **β coefficient** | **β coefficient** | **β coefficient** |
| Total-c | 0.5 (0.48, 0.52) ^***^ | 0.52 (0.48, 0.56) ^***^ | 0.58 (0.43, 0.73) ^***^ | 0.01 (-0.02, 0.03) | 0.01 (-0.10, 0.13) | 0.09 (-0.39, 0.57) |
| Non-HDL-c | 0.18 (0.17, 0.2) ^***^ | 0.26 (0.23, 0.30) ^***^ | 0.36 (0.22, 0.49) ^***^ | -0.29 (-0.31, -0.27) ^***^ | -0.17 (-0.28, -0.07) ^**^ | -0.02 (-0.44, 0.41) |
| Remnant-c | 0.09 (0.08, 0.10) ^***^ | 0.14 (0.12, 0.16) ^***^ | 0.18 (0.12, 0.25) ^***^ | -0.14 (-0.15, -0.13) ^***^ | -0.08 (-0.13, -0.02) ^**^ | 0.02 (-0.19, 0.23) |
| VLDL-c | -0.02 (-0.03, -0.02) ^***^ | 0.02 (0.01, 0.03) ^***^ | 0.07 (0.03, 0.11) ^**^ | -0.15 (-0.16, -0.15) ^***^ | -0.10 (-0.13, -0.07) ^***^ | -0.04 (-0.16, 0.09) |
| LDL-c | 0.09 (0.08, 0.10) ^***^ | 0.13 (0.11, 0.15) ^***^ | 0.17 (0.10, 0.24) ^***^ | -0.15 (-0.16, -0.14) ^***^ | -0.10 (-0.16, -0.04) ^**^ | -0.04 (-0.26, 0.18) |
| HDL-c | 0.32 (0.31, 0.32) ^***^ | 0.25 (0.24, 0.26) ^***^ | 0.22 (0.18, 0.27) ^***^ | 0.29 (0.29, 0.30) ^***^ | 0.19 (0.15, 0.22) ^***^ | 0.11 (-0.01, 0.23) |
| **Triglycerides (mmol/l)** | **Postmeno eugly β** | **Postmeno preDM β** | **Postmeno T2D β** | **Premeno eugly β** | **Premeno preDM β** | **Premeno T2D β** |
| Total TG | -0.2 (-0.21, -0.19)^***^ | -0.12 (-0.14, -0.09) ^***^ | 0.04 (-0.08, 0.15) | -0.40 (-0.42, -0.39) ^***^ | -0.31 (-0.39, -0.23) ^***^ | -0.27 (-0.64, 0.10) |
| TG in VLDL | -0.2 (-0.21, -0.19) ^***^ | -0.13 (-0.15, -0.10) ^***^ | -4.23e-03 (-0.10, 0.09) | -0.36 (-0.38, -0.35) ^***^ | -0.28 (-0.35, -0.21) ^***^ | -0.27 (-0.59, 0.05) |
| TG in LDL | -1.90e-03 (-2.69e-03, -1.11e-03)  ^***^ | 1.24e-03 (-8.32e-04, 3.31e-03) | 0.01 (0.01, 0.02) ^**^ | -0.02 (-0.02, -0.02) ^***^ | -0.02 (-0.02, -0.01) ^***^ | -0.01 (-0.03, 0.03) |
| TG in HDL | 1.23e-03 (3.01e-04, 2.15e. -03) ^**^ | 4.98e-03 (2.65e-03, 7.30e-03) ^***^ | 0.01 (0.01, 0.02) ^**^ | -0.01 (-0.01, -0.01) ^***^ | -0.01 (-0.02, 0) ^**^ | -6.90e-04 (-0.03, 0.03) |
| **Cholesteryl esters (mmol/l)** | **Postmeno eugly β** | **Postmeno preDM β** | **Postmeno T2D β** | **Premeno eugly β** | **Premeno preDM β** | **Premeno T2D β** |
| Total esterified cholesterol | 0.38 (0.37, 0.39) ^***^ | 0.39 (0.36, 0.42) ^***^ | 0.43 (0.32, 0.53) ^***^ | 0.03 (0.02, 0.05) ^***^ | 0.03 (-0.05, 0.11) | 0.07 (-0.27, 0.42) |
| Cholesteryl esters in VLDL | -1.28e-05 (-2.74e-03, 2.72e-03) | 0.02 (0.02, 0.03) ^***^ | 0.04 (0.02, 0.07) ^***^ | -0.08 (-0.08, -0.07) ^***^ | -0.05 (-0.06, -0.03) ^***^ | -4.39e-03 (-0.07, 0.06) |
| Cholesteryl esters in LDL | 0.06 (0.05, 0.06) ^***^ | 0.09 (0.07, 0.10) ^***^ | 0.12 (0.07, 0.18) ^***^ | -0.13 (-0.14, -0.12) ^***^ | -0.09 (-0.13, -0.04) ^***^ | -0.04 (-0.21, 0.13) |
| Cholesteryl esters in HDL | 0.24 (0.24, 0.25) ^***^ | 0.19 (0.18, 0.20) ^***^ | 0.17 (0.13, 0.21) ^***^ | 0.23 (0.22, 0.24) ^***^ | 0.15 (0.12, 0.17) ^***^ | 0.08 (-0.01, 0.17) |
| **Free cholesterol (mmol/l)** | **Postmeno eugly β** | **Postmeno preDM β** | **Postmeno T2D β** | **Premeno eugly β** | **Premeno preDM β** | **Premeno T2D β** |
| Total free cholesterol | 0.12 (0.11, 0.12) ^***^ | 0.13 (0.12, 0.14) ^***^ | 0.15 (0.11, 0.19) ^***^ | -0.03 (-0.04, -0.02) ^***^ | -0.02 (-0.05, 0.02) | 0.02 (-0.13, 0.16) |
| Free cholesterol in VLDL | -2.07e-02 (-2.27e-02, -1.86e-02) ^***^ | -1.05e-03 (-6.16e-03, 4.07e-03) | 0.02 (0, 0.04) ^**^ | -0.08 (-0.08, -0.07) ^***^ | -0.05 (-0.07, -0.04) ^***^ | -0.03 (-0.09, 0.03) |
| Free cholesterol in LDL | 0.04 (0.03, 0.04) ^***^ | 0.04 (0.04, 0.05) ^***^ | 0.05 (0.03, 0.07) ^***^ | -0.02 (-0.03, -0.02) ^***^ | -0.01 (-0.03, 0.00) | -2.37e-04 (-0.06, 0.06) |
| Free cholesterol in HDL | 7.29e-02 (7.16e-02, 7.41e-02) ^***^ | 0.06 (0.05, 0.06) ^***^ | 0.05 (0.04, 0.06) ^***^ | 0.06 (0.06, 0.07) ^***^ | 0.04 (0.03, 0.05) ^***^ | 0.03 (0, 0.06) |
| **Total lipids (mmol/l)** | **Postmeno eugly β** | **Postmeno preDM β** | **Postmeno T2D β** | **Premeno eugly β** | **Premeno preDM β** | **Premeno T2D β** |
| Total lipids in lipoprotein particles | 0.62 (0.59, 0.65) ^***^ | 0.71 (0.63, 0.79) ^***^ | 0.98 (0.69, 1.27) ^***^ | -0.30 (-0.35, -0.26) ^***^ | -0.25 (-0.47, -0.03) ^**^ | -0.12 (-1.17, 0.94) |
| Total lipids in VLDL | -0.27 (-0.28, -0.25) ^***^ | -0.11 (-0.16, -0.07) ^***^ | 0.09 (-0.07, 0.25) | -0.65 (-0.67, -0.63) ^***^ | -0.47 (-0.59, -0.35) ^***^ | -0.36 (-0.88, 0.16) |
| Total lipids in LDL | 0.11 (0.10, 0.12) ^***^ | 0.17 (0.14, 0.19) ^***^ | 0.24 (0.14, 0.34) ^***^ | -0.23 (-0.24, -0.21) ^***^ | -0.15 (-0.23, -0.07) ^***^ | -0.06 (-0.37, 0.25) |
| Total lipids in HDL | 0.62 (0.61, 0.63) ^***^ | 0.50 (0.48, 0.53) ^***^ | 0.48 (0.38, 0.58) ^***^ | 0.56 (0.54, 0.57) ^***^ | 0.35 (0.28, 0.41) ^***^ | 0.22 (-0.07, 0.50) |
| **Phospholipids (mmol/l)** | **Postmeno eugly β** | **Postmeno preDM β** | **Postmeno T2D β** | **Premeno eugly β** | **Premeno preDM β** | **Premeno T2D β** |
| Total phospholipids | 0.32 (0.31, 0.33) ^***^ | 0.31 (0.29, 0.33) ^***^ | 0.36 (0.28, 0.45) ^***^ | 0.09 (0.08, 0.10) ^***^ | 0.05 (-0.01, 0.11) | 0.06 (-0.22, 0.35) |
| Phospholipids in VLDL | -0.04 (-0.05, -0.04) ^***^ | -0.01 (-0.02, 0) ^**^ | 0.03 (0.00, 0.06) ^*^ | -0.13 (-0.14, -0.13) ^***^ | -0.09 (-0.12, -0.07) ^***^ | -0.06 (-0.16, 0.05) |
| Phospholipids in LDL | 0.02 (0.02, 0.03) ^***^ | 0.04 (0.03, 0.04) ^***^ | 0.05 (0.03, 0.08) ^***^ | -0.05 (-0.06, -0.05) ^***^ | -0.03 (-0.05, -0.02) ^***^ | -0.01 (-0.08, 0.06) |
| Phospholipids in HDL | 0.30 (0.30, 0.31) ^***^ | 0.25 (0.23, 0.26) ^***^ | 0.24 (0.19, 0.29) ^***^ | 0.27 (0.27, 0.28) ^***^ | 0.17 (0.14, 0.20) ^***^ | 0.11 (-0.04, 0.26) |
| **Apolipoproteins (g/l)** | **Postmeno eugly β** | **Postmeno preDM β** | **Postmeno T2D β** | **Premeno eugly β** | **Premeno preDM β** | **Premeno T2D β** |
| Apolipoprotein B | 1.92e-02 (1.55e-02, 2.30e-02)  ^***^ | 0.04 (0.03, 0.05) ^***^ | 0.07 (0.04, 0.10) ^***^ | -0.09 (-0.10, -0.09) ^***^ | -0.06 (-0.08, -0.03) ^***^ | -0.01 (-0.11, 0.09) |
| Apolipoprotein A1 | 0.22 (0.22, 0.23) ^***^ | 0.19 (0.18, 0.20) ^***^ | 0.18 (0.14, 0.23) ^***^ | 0.18 (0.18, 0.19) ^***^ | 0.11 (0.09, 0.14) ^***^ | 0.06 (-0.06, 0.18) |
| Apo(B) to apo(A1) ratio | -0.08 (-0.08, -0.07) ^***^ | -0.06 (-0.06, -0.04) ^***^ | -0.02 (-0.04, -0.01) | -0.14 (-0.15, -0.14) ^***^ | -0.10 (-0.12, -0.08) ^***^ | -0.03 (-0.12, 0.05) |
| **Lipoprotein particle size (nm)** | **Postmeno eugly β** | **Postmeno preDM β** | **Postmeno T2D β** | **Premeno eugly β** | **Premeno preDM β** | **Premeno T2D β** |
| Average diameter for VLDL particles | -0.84 (-0.86, -0.82) ^***^ | -0.60 (-0.65, -0.54) ^***^ | -0.50 (-0.75, -0.24) ^***^ | -1.29 (-1.32, -1.26) ^***^ | -0.84 (-0.99, -0.68) ^***^ | -0.97 (-1.54, -0.40) ^**^ |
| Average diameter for LDL particles | 3.87e-02 (3.69e-02, 4.04e-02) ^***^ | 0.03 (0.03, 0.04) ^***^ | 0.03 (0.01, 0.05) ^**^ | 4.07e-02 (3.84e-02, 4.31e-02) ^***^ | 0.03 (0.02, 0.05) ^***^ | 0.06 (0.02, 0.10) ^*^ |
| Average diameter for HDL particles | 1.90e-01 (1.86e-01, 1.94e-01) ^***^ | 0.14 (0.13, 0.15) ^***^ | 0.12 (0.09, 0.15) ^***^ | 0.23 (0.22, 0.23) ^***^ | 0.15 (0.13, 0.17) ^***^ | 0.12 (0.07, 0.18) ^**^ |
| **Total concentration of lipoprotein particles** | 1.99e-03 (1.94e-03, 2.03e-03) ^***^ | 1.85e-03 (1.74e-03, 1.96e-03) ^***^ | 1.95e-03 (1.51e-03, 2.40e-03) ^***^ | 1.14e-03 (1.08e-03, 1.20e-03) ^***^ | 7.36e-04 (4.41e-04, 1.03e-03) ^***^ | 2.17e-04 (-1.21e-03, 1.64e-03) |
| Concentration of VLDL particles | -4.75e-06 (-5.61e-06, -3.89e-06) ^***^ | 2.46e-06 (3.41e-07, 4.58e-06) ^**^ | 1.21e-05 (4.47e-06, 1.98e-05) ^**^ | -2.74e-05 (-2.85e-05, -2.63e-05) ^***^ | -1.88e-05 (-2.47e-05, -1.28e-05) ^***^ | -6.68e-06 (-3.11e-05, 1.78e-05) |
| Concentration of LDL particles | 2.05e-05 (1.52e-05, 2.58e-05) ^***^ | 5.38e-05 (4.09e-05, 6.67e-05) ^***^ | 8.81e-05 (4.25e-05, 1.34e-04) ^***^ | -1.36e-04 (-1.44e-04, -1.29e-04) ^***^ | -8.80e-05 (-1.24e-04, -5.15e-05) ^***^ | -2.72e-05 (-1.71e-04, 1.16e-04) |
| Concentration of HDL particles | 1.95e-03 (1.90e-03, 1.99e-03) ^***^ | 1.76e-03 (1.66e-03, 1.87e-03) ^***^ | 1.82e-03 (1.39e-03, 2.25e-03) ^***^ | 1.32e-03 (1.26e-03, 1.38e-03) ^***^ | 8.49e-04 (5.70e-04, 1.13e-03) ^***^ | 2.34e-04 (-1.11e-03, 1.58e-03) |
| **Lipoprotein particle size** | **Postmeno eugly β** | **Postmeno preDM β** | **Postmeno T2D β** | **Premeno eugly β** | **Premeno preDM β** | **Premeno T2D β** |
| XXL_VLDL | -1.91e-02 (-2.00e-02, -1.83e-02) ^***^ | -1.11e-02 (-1.32e-02, -8.99e-03) ^***^ | -1.23e-03 (-9.34e-03, 6.88e-03) | -3.31e-02 (-3.42e-02, -3.20e-02) ^***^ | -0.02 (-0.03, -0.02) ^***^ | -0.02 (-0.05, 0) |
| XL_VLDL | -1.17e-02 (-1.23e-02, -1.12e-02) ^***^ | -5.72e-03 (-7.07e-03, -4.36e-03) ^***^ | 7.29e-04 (-4.26e-03, 5.71e-03) | -0.02 (-0.03, -0.02) ^***^ | -0.02 (-0.02, -0.01) ^***^ | -0.01 (-0.03, 0) |
| L_VLDL | -1.81e-02 (-1.91e-02, -1.72e-02) ^***^ | -8.18e-03 (-1.04e-02, -5.95e-03) ^***^ | 1.81e-03 (-6.36e-03, 9.98e-03) | -4.02e-02 (-4.14e-02, -3.90e-02) ^***^ | -0.03 (-0.03, -0.02) ^***^ | -0.02 (-0.04, 0) |
| M_VLDL | 9.30e-03 (8.08e-03, 1.05e-02) ^***^ | 1.76e-02 (1.47e-02, 2.06e-02) ^***^ | 0.02 (0.01, 0.03) ^***^ | -0.03 (-0.03, -0.02) ^***^ | -0.01 (-0.02, -0.01) ^**^ | 6.19e-04 (-2.99e-02, 3.12e-02) |
| S_VLDL | -3.58e-03 (-4.57e-03, -2.59e-03) ^***^ | 5.72e-03 (3.33e-03, 8.11e-03) ^***^ | 0.01 (0.01, 0.02) ^**^ | -3.09e-02 (-3.22e-02, -2.96e-02) ^***^ | -0.02 (-0.03, -0.01) ^***^ | -1.92e-03 (-2.58e-02, 2.19e-02) |
| XS_VLDL | 2.26e-02 (2.17e-02, 2.35e-02) ^***^ | 2.42e-02 (2.20e-02, 2.64e-02) ^***^ | 0.03 (0.02, 0.04) ^***^ | -8.22e-04 (-3.75e-04, 2.02e-03) | 0 (0, 0.01) | 0.02 (0, 0.04) |
| IDL | 0.11 (0.11, 0.12) ^***^ | 1.14e-01 (1.05e-01, 1.23e-01) ^***^ | 0.12 (0.09, 0.15) ^***^ | 0.02 (0.01, 0.02) ^***^ | 0.02 (0.00, 0.05) | 0.06 (-0.04, 0.16) |
| L_LDL | 0.09 (0.08, 0.09) ^***^ | 1.04e-01 (9.14e-02, 1.17e-01) ^***^ | 0.13 (0.08, 0.17) ^***^ | -0.06 (-0.07, -0.06) ^***^ | -0.04 (-0.07, 0.00) | 2.42e-03 (-0.13, 0.14) |
| M_LDL | 2.15e-03 (-7.84e-05, 4.38e-03) ^*^ | 1.80e-02 (1.25e-02, 2.34e-02) ^***^ | 0.03 (0.01, 0.05) ^**^ | -0.07 (-0.07, -0.06) ^***^ | -0.04 (-0.06, -0.03) ^***^ | -0.03 (-0.09, 0.03) |
| S_LDL | 4.68e-04 (-3.64e-04, 1.30e-03) | 6.00e-03 (3.97e-03, 8.03e-03) ^***^ | 0.01 (0, 0.02) ^**^ | -0.02 (-0.03, -0.02) ^***^ | -0.02 (-0.02, -0.01) ^***^ | -0.01 (-0.03, 0.01) |
| XL_HDL | 2.96e-02 (2.90e-02, 3.03e-02) ^***^ | 2.02e-02 (1.89e-02, 2.15e-02) ^***^ | 0.02 (0.01, 0.02) ^***^ | 3.06e-02 (2.99e-02, 3.14e-02) ^***^ | 0.02 (0.01, 0.02) ^***^ | 0.02 (0.01, 0.03) ^*^ |
| L_HDL | 1.60e-01 (1.57e-01, 1.63e-01) ^***^ | 1.17e-01 (1.10e-01, 1.23e-01) ^***^ | 0.09 (0.07, 0.11) ^***^ | 1.69e-01 (1.65e-01, 1.72e-01) ^***^ | 0.11 (0.09, 0.12) ^***^ | 0.07 (0.03, 0.12) ^**^ |
| M_HDL | 1.10e-01 (1.08e-01, 1.12e-01) ^***^ | 9.17e-02 (8.67e-02, 9.68e-02) ^***^ | 0.09 (0.07, 0.11) ^***^ | 1.004-01 (9.73e-02, 1.03e-01) ^***^ | 0.06 (0.05, 0.08) ^***^ | 0.03 (-0.03, 0.09) |
| S_HDL | 1.75e-02 (1.63e-02, 1.87e-02) ^***^ | 2.35e-02 (2.07e-02, 2.64e-02) ^***^ | 0.03 (0.02, 0.04) ^***^ | -5.07e-03 (-6.67e-03, -3.46e-03) ^***^ | -2.08e-03 (-9.80e-03, 5.64e-03) | -0.02 (-0.05, 0.02) |
| **Other lipids (mmol/L)** | **Postmeno eugly β** | **Postmeno preDM β** | **Postmeno T2D β** | **Premeno eugly β** | **Premeno preDM β** | **Premeno T2D β** |
| Phosphoglycerides | 0.29 (0.28, 0.30) ^***^ | 0.28 (0.26, 0.29) ^***^ | 0.32 (0.25, 0.39) ^***^ | 0.13 (0.12, 0.14) ^***^ | 0.07 (0.02, 0.12) ^**^ | 0.06 (-0.19, 0.31) |
| TG to phosphoglycerides ratio | -0.16 (-0.16, -0.15) ^***^ | -0.13 (-0.14, -0.12) ^***^ | -0.09 (-0.13, -0.04) ^***^ | -0.21 (-0.22, -0.21) ^***^ | -0.16 (-0.19, -0.13) ^***^ | -0.16 (-0.26, -0.05) ^*^ |
| Total cholines | 0.32 (0.31, 0.32) ^***^ | 0.30 (0.28, 0.32) ^***^ | 0.33 (0.23, 0.36) ^***^ | 0.14 (0.13, 0.15) ^***^ | 0.09 (0.04, 0.14) ^**^ | 0.07 (-0.17, 0.32) |
| Phosphatidylcholines | 0.29 (0.28, 0.30) ^***^ | 0.27 (0.26, 0.29) ^***^ | 0.30 (0.23, 0.36) ^***^ | 0.16 (0.15, 0.17) ^***^ | 0.10 (0.05, 0.14) ^***^ | 0.08 (-0.15, 0.30) |
| Sphingomyelins | 0.06 (0.05, 0.06) ^***^ | 5.06e-02 (4.76e-02, 5.37e-02) ^***^ | 0.06 (0.05, 0.07) ^***^ | 2.35e-02 (2.18e-02, 2.52e-02) ^***^ | 0.02 (0.01, 0.02) ^***^ | 0.02 (-0.01, 0.06) |
| **Amino acids (mmol/L)** | **Postmeno eugly β** | **Postmeno preDM β** | **Postmeno T2D β** | **Premeno eugly β** | **Premeno preDM β** | **Premeno T2D β** |
| Alanine | -1.37e-02 (-1.52e-02, -1.22e-02) ^***^ | -0.01 (-0.01, 0) ^**^ | 0 (-0.01, 0.02) | -1.95e-02 (-2.17e-02, -1.74e-02) ^***^ | -0.01 (-0.02, 0.00) | 0.01 (-0.04, 0.06) |
| Glutamine | -7.56e-03 (-9.15e-03, -5.98e-03) ^***^ | -5.83e-04 (-4.33e-03, 3.17e-03) | -0.01 (-0.02, 0.01) | -0.03 (-0.04, -0.03) ^***^ | -0.04 (-0.05, -0.03) ^***^ | -0.04 (-0.08, 0.01) |
| Glycine | 0.04 (0.04, 0.05) ^***^ | 4.00e-02 (3.74e-02, 4.27e-02) ^***^ | 0.02 (0.01, 0.03) ^***^ | 4.05e-02 (3.88e-02, 4.22e-02) ^***^ | 0.04 (0.03, 0.04) ^***^ | 0.03 (0.01, 0.06) |
| Histidine | -1.76e-03 (-1.97e-03, -1.56e-03) ^***^ | -1.99e-03 (-2.50e-03, -1.49e-03) ^***^ | -8.10e-05 (-2.04e-03, 1.88e-03) | -2.78e-03 (-3.07e-03, -2.48e-03) ^***^ | -4.49e-03 (-6.23e-03, -2.74e-03) ^***^ | -3.96e-03 (-8.20-03, 2.85e-04) |
| **Aromatic amino acids (mmol/L)** | **Postmeno eugly β** | **Postmeno preDM β** | **Postmeno T2D β** | **Premeno eugly β** | **Premeno preDM β** | **Premeno T2D β** |
| Phenylalanine | -1.38e-03 (-1.61e-03, -1.15e-03) ^***^ | -1.62e-03 (-2.12e-03, -1.13e-03) ^***^ | -7.97e-04 (-2.81e-03, 1.22e-03) | -5.47e-04 (-8.79e-04, -2.14e-04) ^**^ | -5.19e-04 (-1.90e-03, 8.64e-04) | 2.69e-03 (-1.38e-03, 6.76e-03) |
| Tyrosine | 1.05e-03 (7.70e-04, 1.33e-03) ^***^ | -6.78e-05 (-7.54e-04, 6.18e-04) | 3.97e-04 (-2.42e-03, 3.21e-03) | -2.00e-03 (-2.37e-03, -1.64e-03) ^***^ | -2.84e-03 (-4.69e-03, -9.86e-04) ^**^ | 9.98e-04 (-6.62e-03, 8.61e-03) |
| **Branched-chain amino acids (BCAA) (mmol/L)** | **Postmeno eugly β** | **Postmeno preDM β** | **Postmeno T2D β** | **Premeno eugly β** | **Premeno preDM β** | **Premeno T2D β** |
| Isoleucine | -6.29e-03 (-6.64e-03, -5.94e-03) ^***^ | -5.93e-03 (-6.78e-03, -5.09e-03) ^***^ | -4.31e-03 (-7.88e-03, -7.41e-04) ^**^ | -8.97e-03 (-9.44e-03, -8.50e-03) ^***^ | -0.01 (-0.01, 0.00) ^***^ | -0.01 (-0.01, 0) |
| Leucine | -1.42e-02 (-1.47e-02, -1.36e-02) ^***^ | -1.24e-02 (-1.37e-02, -1.11e-02) ^***^ | -1.06e-02 (-1.63e-02, -4.97e-03) ^***^ | -1.93e-02 (-2.00e-02, -1.85e-02) ^***^ | -0.02 (-0.02, -0.01) ^***^ | -0.02 (-0.03, 0) |
| Valine | -1.65e-02 (-1.73e-02, -1.56e-02) ^***^ | -1.40e-02 (-1.60e-02, -1.20e-02) ^***^ | -8.13e-03 (-1.63e-02, 6.01e-05) ^*^ | -2.76e-02 (-2.87e-02, -2.65e-02) ^***^ | -0.02 (-0.03, -0.01) ^***^ | -0.02 (-0.04, 0.01) |
| Total BCAA | -3.69e-02 (-3.85e-02, -3.53e-02) ^***^ | -3.24e-02 (-3.63e-02, -2.84e-02) ^***^ | -0.02 (-0.04, -0.01) ^**^ | -0.06 (-0.06, -0.05) ^***^ | -0.04 (-0.05, -0.03) ^***^ | -0.04 (-0.09, 0.01) |
| **Ketones (mmol/L)** | **Postmeno eugly β** | **Postmeno preDM β** | **Postmeno T2D β** | **Premeno eugly β** | **Premeno preDM β** | **Premeno T2D β** |
| 3-hydroxybutyrate | 3.79e-03 (2.52e-03, 5.06e-03) ^***^ | -1.27e-03 (-3.50e-03, 9.55e-04) | 8.45e-05 (-8.86e-03, 9.03e-03) | 6.43e-03 (4.78e-03, 8.07e-03) ^***^ | 1.26e-03 (-3.62e-03, 6.15e-03) | -5.75e-03 (-1.97e-02, 8.23e-03) |
| Acetoacetate | 6.54e-04 (3.96e-04, 9.12e-04) ^***^ | -5.75e-04 (-1.09e-03, -5.95e-05) ^**^ | 1.85e-03 (-2.20e-04, 3.92e-03) | 3.04e-04 (-2.38e-05, 6.31e-04) ^*^ | -1.09e-03 (-2.13e-03, -6.04e-05) ^*^ | -1.15e-03 (-5.54e-03, 3.25e-03) |
| Acetone | -1.28e-06 (-1.17e-04, 1.15e-04) | -5.03e-04 (-7.31e-04, -2.76e-04) ^***^ | 2.70e-04 (-6.93e-04, 1.23e-03) | -3.45e-05 (-1.83e-04, 1.14e-04) | -3.52e-04 (-8.30e-04, 1.26e-04) | -3.07e-04 (-2.11e-03, 1.50e-03) |
| Acetate | 9.39e-06 (-2.37e-04, 2.56e-04) | -5.88e-04 (-1.15e-03, -2.62e-05) ^**^ | -2.20e-05 (-1.35e-03, 1.31e-03) | -4.87e-05 (-3.43e-04, 2.45e-04) | 9.50e-06 (-8.51e-04, 8.70e-04) | 6.49e-04 (-2.38e-03, 3.67e-03) |
| **Inflammation (mmol/l)** | **Postmeno eugly β** | **Postmeno preDM β** | **Postmeno T2D β** | **Premeno eugly β** | **Premeno preDM β** | **Premeno T2D β** |
| Glycoprotein acetyls (GlycA) | 4.68e-03 (2.50e-03, 6.85e-03) ^***^ | 0.02 (0.01, 0.03) ^***^ | 0.06 (0.04, 0.08) ^***^ | -3.89e-02 (-4.19e-02, -3.59e-02) ^***^ | -3.67e-04 (-1.58e-02, 1.50e-02) | 0.02 (-0.04, 0.09) |
| **Electrolyte (mmol/l )** | **Postmeno eugly β** | **Postmeno preDM β** | **Postmeno T2D β** | **Premeno eugly β** | **Premeno preDM β** | **Premeno T2D β** |
| Creatinine | -1.32e-02 (-1.35e-02, -1.30e-02) ^***^ | -1.27e-02 (-1.33e-02, -1.21e-02) ^***^ | -1.20e-02 (-1.43e-02, -9.60e-03) ^***^ | -1.43e-02 (-1.45e-02, -1.40e-02) ^***^ | -1.25e-02 (-1.40e-02, -1.10e-02) ^***^ | -7.18e-03 (-1.23e-02, -2.02e-03) ^*^ |
| Albumin | 1.43e-01 (7.71e-02, 2.08e-01) ^***^ | 0.2 (0.04, 0.36) ^**^ | -0.47 (-1.12, 0.17) | -1.22 (-1.31, -1.13)  ^***^ | -1.26 (-1.71, -0.81) ^***^ | -2.65 (-4.56, -0.74) ^*^ |
| **Fatty acids (mmol/l)** | **Postmeno eugly β** | **Postmeno preDM β** | **Postmeno T2D β** | **Premeno eugly β** | **Premeno preDM β** | **Premeno T2D β** |
| Total fatty acids | 0.49 (0.45, 0.54) ^***^ | 0.67 (0.56, 0.79) ^***^ | 1.22 (0.74, 1.70) ^***^ | -0.79 (-0.85, -0.72) ^***^ | -0.64 (-0.98, -0.30) ^***^ | -0.52 (-2.22, 1.18) |
| Omega-3 fatty acids | 0.09 (0.09, 0.1) ^***^ | 0.09 (0.08, 0.10) ^***^ | 0.10 (0.05, 0.14) ^***^ | -0.01 (-0.01, 0.00) ^**^ | -0.03 (-0.05, 0) ^**^ | 3.85e-03 (-0.12, 0.12) |
| Omega-6 fatty acids | 0.29 (0.27, 0.3) ^***^ | 0.34 (0.31, 0.37) ^***^ | 0.39 (0.26, 0.51) ^***^ | -0.08 (-0.10, -0.06) ^***^ | -0.03 (-0.12, 0.06) | -0.05 (-0.44, 0.34) |
| Polyunsaturated fatty acids (PUFA) | 0.38 (0.37, 0.4) ^***^ | 0.43 (0.39, 0.46) ^***^ | 0.48 (0.33, 0.63) ^***^ | -0.09 (-0.11, -0.07) ^***^ | -0.06 (-0.16, 0.05) | -0.05 (-0.51, 0.41) |
| Monounsaturated fatty acids (MUFA) | -0.04 (-0.06, -0.03) ^***^ | 0.05 (0.01, 0.09) ^**^ | 0.27 (0.10, 0.44) ^**^ | -0.41 (-0.43, -0.39) ^***^ | -0.3 (-0.42, -0.19) ^***^ | -0.25 (-0.83, 0.32) |
| Saturated fatty acids (SFA) | 0.15 (0.13, 0.17) ^***^ | 0.20 (0.15, 0.25) ^***^ | 0.47 (0.28, 0.66) ^***^ | -0.29 (-0.31, -0.26) ^***^ | -0.28 (-0.42, -0.14) ^***^ | -0.22 (-0.95, 0.51) |
| Linoleic acid | 0.22 (0.2, 0.23) ^***^ | 0.28 (0.25, 0.31) ^***^ | 0.29 (0.17, 0.41) ^***^ | -0.10 (-0.12, -0.09) ^***^ | -0.02 (-0.11, 0.07) | -0.07 (-0.43, 0.3) |
| Docosahexaenoic acid (DHA) | 4.85e-02 (4.68e-02, 5.02e-02) ^***^ | 0.04 (0.04, 0.05) ^***^ | 0.04 (0.02, 0.05) ^***^ | 2.09e-02 (1.89e-02, 2.28e-02) ^***^ | 0.01 (0, 0.02) | 0.02 (-0.03, 0.06) |
| **Fatty acid to total ratio (%)** | **Postmeno eugly β** | **Postmeno preDM β** | **Postmeno T2D β** | **Premeno eugly β** | **Premeno preDM β** | **Premeno T2D β** |
| Omega-3 fatty acids to total fatty acids | 6.11e-03 (5.79e-03, 6.43e-03) ^***^ | 4.80e-03 (4.08e-03, 5.52e-03) ^***^ | 3.20e-03 (3.95e-04, 6.01e-03) ^**^ | 2.26e-03 (1.86e-03, 2.65e-03) ^***^ | -3.59e-04 (-2.14e-03, 1.43e-03) | 1.54e-03(-0.01, 0.01) |
| Omega-6 fatty acids to total fatty acids | 6.77e-03 (6.10e-03, 7.44e-03) ^***^ | 6.13e-03 (4.42e-03, 7.85e-03) ^***^ | -5.60e-03 (-1.29e-02, 1.75e-03) | 1.77e-02 (1.68e-02, 1.86e-02) ^***^ | 0.02 (0.01, 0.02) ^***^ | 0.01 (-0.01, 0.03) |
| PUFA to total fatty acids | 1.29e-02 (1.22e-02, 1.36e-02) ^***^ | 1.09e-02 (9.16e-03, 1.27e-02) ^***^ | -2.40e-03 (-1.00e-02, 5.21e-03) | 1.99e-02 (1.90e-02, 2.09e-02) ^***^ | 0.02 (0.01, 0.02) ^***^ | 0.01 (-0.01, 0.04) |
| MUFA to total fatty acids | -1.22e-02 (-1.27e-02, -1.18e-02) ^***^ | -9.05e-03 (-1.02e-02, -7.86e-03) ^***^ | -2.73e-03 (-8.06e-03, 2.60e-03) | -1.91e-02 (-1.98e-02, -1.84e-02) ^***^ | -0.01 (-0.02, -0.01) ^***^ | -0.01 (-0.03, 0) |
| Saturated fatty acids to total fatty acids | -6.43e-04 (-1.02e-03, -2.62e-04) ^**^ | -1.88e-03 (-2.85e-03, -9.11e-04) ^***^ | 5.13e-03 (1.06e-03, 9.19e-03) ^**^ | -8.50e-04 (-1.37e-03, -3.27e-04) ^**^ | -3.83e-03 (-6.80e-03, -8.68e-04) ^**^ | -3.17e-03 (-0.02, 0.01) |
| Linoleic acid to total fatty acids | 4.91e-03 (4.28e-03, 5.53e-03) ^***^ | 6.59e-03 (5.06e-03, 8.13e-03) ^***^ | -3.58e-03 (-9.82e-03, 2.66e-03) | 9.88e-03 (9.03e-03, 1.07e-02) ^***^ | 1.22e-02 (7.73e-03, 1.67e-02) ^***^ | 0.01 (-0.01, 0.02) |
| DHA to total fatty acids | 3.12e-03 (2.98e-03, 3.25e-03) ^***^ | 2.33e-03 (2.02e-03, 2.65e-03) ^***^ | 1.04e-03 (-2.29e-04, 2.31e-03) | 3.08e-03 (2.91e-03, 3.25e-03) ^***^ | 1.43e-03 (6.11e-04, 2.25e-03) ^**^ | 2.17e-03 (-1.46e-03, 5.80e-03) |
| PUFA to MUFA ratio | 0.15 (0.14, 0.15) ^***^ | 0.11 (0.09, 0.12) ^***^ | 4.44e-03 (-6.15e-02, 7.04e-02) | 2.34e-01 (2.25e-01, 2.43e-01) ^***^ | 0.15 (0.1, 0.19) ^***^ | 0.12 (-0.04, 0.28) |
| Omega-6 fatty acids to omega-3 fatty acids | -1.26 (-1.34, -1.18) ^***^ | -1.12 (-1.32, -0.91) ^***^ | -0.7 (-1.3, -0.09) ^**^ | -0.14 (-0.27, -0.02) ^**^ | 0.1 (-0.59, 0.79) | -0.07 (-2.16, 2.03) |
| Degree of unsaturation | 4.75e-02 (4.60e-02, 4.90e-02) ^***^ | 4.06e-02 (3.68e-02, 4.43e-02) ^***^ | 0.02 (0.01, 0.04) ^**^ | 3.77e-02 (3.57e-02, 3.96e-02) ^***^ | 0.02 (0.01, 0.03) ^***^ | 0.03 (-0.02, 0.07) |

*FDR-adjusted p<.1; ** <.05; ***<.0001

Fully adjusted for age, race, income, area deprivation, smoking, alcohol drinking, physical activity, BMI, and medications for hypertension, hyperlipidemia, and diabetes.
