## Supplemental Table 3 for "Sex differences in circulating metabolites across glycemic status and risk of coronary heart disease"

**Supplemental Table** **3. Hazard Ratios (95% CI) of Metabolites and Risk of CHD by Sex Across Glycemic Status**

|  | **New T2D** |  |  |  | **Pre-DM** |  |  |  | **Euglycemia** |  |  |  |
| --- | --- | --- | --- | --- | --- | --- | --- | --- | --- | --- | --- | --- |
|  | **All** | **Men** | **Women** | **P-interaction** | **All** | **Men** | **Women** | **P-interaction** | **All** | **Men** | **Women** | **P-interaction** |
| **Cholesterol (mmol/l)** |  |  |  |  |  |  |  |  |  |  |  |  |
| Total-c | 0.90 (0.39, 2.07) | 0.91 (0.35, 2.39) | 0.94 (0.18, 5.08) | 0.943 | 1.04 (0.71, 1.51) | 0.95 (0.61, 1.48) | 1.17 (0.60, 2.30) | 0.568 | 1.39 (1.15, 1.68) ** | 1.41 (1.13, 1.77) ** | 1.17 (0.81, 1.69) | 0.586 |
| Non-HDL-c | 1.05 (0.54, 2.02) | 1.08 (0.49, 2.36) | 0.98 (0.29, 3.38) | 0.971 | 1.18 (0.87, 1.59) | 1.10 (0.76, 1.58) | 1.27 (0.75, 2.16) | 0.612 | 1.58 (1.36, 1.83) *** | 1.53 (1.28, 1.84) *** | 1.52 (1.15, 2.01) ** | 0.863 |
| Remnant-c | 1.12 (0.59, 2.13) | 1.16 (0.55, 2.48) | 1.03 (0.31, 3.41) | 0.937 | 1.25 (0.93, 1.67) | 1.14 (0.80, 1.63) | 1.39 (0.84, 2.31) | 0.47 | 1.62 (1.40, 1.88) *** | 1.58 (1.32, 1.88) *** | 1.59 (1.22, 2.07) ** | 0.775 |
| VLDL-c | 1.31 (0.79, 2.16) | 1.28 (0.69, 2.38) | 1.33 (0.56, 3.14) | 0.884 | 1.16 (0.93, 1.45) | 1.11 (0.84, 1.46) | 1.20 (0.83, 1.74) | 0.632 | 1.52 (1.36, 1.70) *** | 1.46 (1.27, 1.67) *** | 1.55 (1.28, 1.88) *** | 0.474 |
| LDL-c | 0.99 (0.52, 1.86) | 1.01 (0.47, 2.15) | 0.96 (0.29, 3.15) | 0.989 | 1.09 (0.82, 1.46) | 1.05 (0.74, 1.48) | 1.13 (0.67, 1.90) | 0.801 | 1.47 (1.27, 1.70) *** | 1.44 (1.21, 1.72) *** | 1.39 (1.05, 1.83) ** | 0.984 |
| HDL-c | 0.62 (0.25, 1.51) | 0.57 (0.19, 1.66) | 0.94 (0.18, 4.91) | 0.697 | 0.60 (0.42, 0.85) ** | 0.58 (0.37, 0.90) | 0.65 (0.36, 1.18) | 0.735 | 0.50 (0.42, 0.59) *** | 0.56 (0.45, 0.69) *** | 0.40 (0.29, 0.55) *** | 0.105 |
| **Triglycerides (mmol/l)** | **T2D_all** | **T2D_M** | **T2D_W** | **P-interaction** | **PreDM_all** | **PreDM_M** | **PreDM_W** | **P-interaction** | **Eugly_all** | **Eulgy_M** | **Eugly_W** | **P-interaction** |
| Total TG | 1.41 (0.92, 2.16) | 1.10 (0.66, 1.85) | 2.46 (1.06, 5.72) | 0.107 | 0.97 (0.81, 1.15) | 0.95 (0.77, 1.19) | 0.94 (0.70, 1.27) | 0.884 | 1.31 (1.20, 1.43) *** | 1.22 (1.10, 1.36) *** | 1.45 (1.24, 1.70) *** | 0.053 |
| TG in VLDL | 1.35 (0.94, 1.93) | 1.10 (0.71, 1.70) | 2.05 (1.02, 4.13) | 0.136 | 0.95 (0.83, 1.10) | 0.96 (0.80, 1.15) | 0.91 (0.72, 1.16) | 0.911 | 1.22 (1.14, 1.31) *** | 1.15 (1.06, 1.26) ** | 1.32 (1.16, 1.49) *** | 0.069 |
| TG in LDL | 1.50 (0.80, 2.82) | 1.15 (0.54, 2.44) | 3.14 (0.93, 10.68) | 0.145 | 1.20 (0.91, 1.57) | 1.10 (0.79, 1.54) | 1.32 (0.82, 2.12) | 0.454 | 1.73 (1.52, 1.98) *** | 1.61 (1.37, 1.89) *** | 1.95 (1.52, 2.51) *** | 0.156 |
| TG in HDL | 1.35 (0.77, 2.36) | 0.97 (0.50, 1.88) | 3.30 (1.04, 10.49) | 0.068 | 0.92 (0.73, 1.16) | 0.87 (0.66, 1.16) | 0.98 (0.66, 1.46) | 0.498 | 1.29 (1.15, 1.44) *** | 1.15 (1.00, 1.32) * | 1.55 (1.27, 1.90) *** | 0.011 |
| **Cholesteryl esters (mmol/l)** | **T2D_all** | **T2D_M** | **T2D_W** | **P-interaction** | **PreDM_all** | **PreDM_M** | **PreDM_W** | **P-interaction** | **Eugly_all** | **Eulgy_M** | **Eugly_W** | **P-interaction** |
| Total esterified cholesterol | 0.85 (0.37, 1.98) | 0.87 (0.33, 2.30) | 0.89 (0.16, 4.95) | 0.953 | 1.01 (0.69, 1.47) | 0.93 (0.59, 1.46) | 1.13 (0.57, 2.23) | 0.605 | 1.33 (1.09, 1.61) ** | 1.35 (1.08, 1.70) ** | 1.11 (0.76, 1.61) | 0.564 |
| Cholesteryl esters in VLDL | 1.24 (0.74, 2.07) | 1.30 (0.69, 2.44) | 1.12 (0.47, 2.68) | 0.858 | 1.22 (0.97, 1.54) | 1.15 (0.86, 1.53) | 1.30 (0.88, 1.91) | 0.55 | 1.55 (1.38, 1.73) *** | 1.49 (1.29, 1.71) *** | 1.57 (1.28, 1.91) *** | 0.549 |
| Cholesteryl esters in LDL | 1.03 (0.54, 1.95) | 1.02 (0.47, 2.18) | 1.09 (0.34, 3.56) | 0.847 | 1.07 (0.80, 1.43) | 1.03 (0.73, 1.46) | 1.10 (0.65, 1.84) | 0.821 | 1.50 (1.30, 1.74) *** | 1.46 (1.23, 1.74) *** | 1.45 (1.10, 1.91) ** | 0.867 |
| Cholesteryl esters in HDL | 0.59 (0.25, 1.43) | 0.56 (0.19, 1.63) | 0.82 (0.16, 4.18) | 0.786 | 0.59 (0.41, 0.83) ** | 0.57 (0.37, 0.88) | 0.64 (0.36, 1.15) | 0.733 | 0.48 (0.40, 0.57) *** | 0.54 (0.44, 0.66) *** | 0.39 (0.29, 0.54) *** | 0.123 |
| **Free cholesterol (mmol/l)** | **T2D_all** | **T2D_M** | **T2D_W** | **P-interaction** | **PreDM_all** | **PreDM_M** | **PreDM_W** | **P-interaction** | **Eugly_all** | **Eulgy_M** | **Eugly_W** | **P-interaction** |
| Total free cholesterol | 1.02 (0.46, 2.27) | 1.03 (0.41, 2.61) | 1.07 (0.22, 5.12) | 0.93 | 1.10 (0.77, 1.58) | 1.00 (0.65, 1.54) | 1.26 (0.66, 2.40) | 0.496 | 1.52 (1.27, 1.83) *** | 1.54 (1.24, 1.91) *** | 1.31 (0.93, 1.86) | 0.632 |
| Free cholesterol in VLDL | 1.36 (0.85, 2.16) | 1.23 (0.69, 2.21) | 1.57 (0.69, 3.58) | 0.597 | 1.09 (0.89, 1.34) | 1.06 (0.82, 1.37) | 1.10 (0.78, 1.54) | 0.75 | 1.45 (1.31, 1.61) *** | 1.38 (1.22, 1.57) *** | 1.51 (1.27, 1.80) *** | 0.34 |
| Free cholesterol in LDL | 0.90 (0.51, 1.61) | 1.02 (0.51, 2.01) | 0.70 (0.25, 2.01) | 0.614 | 1.13 (0.86, 1.47) | 1.09 (0.79, 1.49) | 1.18 (0.71, 1.94) | 0.801 | 1.32 (1.15, 1.52) *** | 1.32 (1.12, 1.56) ** | 1.20 (0.91, 1.58) | 0.746 |
| Free cholesterol in HDL | 0.78 (0.33, 1.84) | 0.66 (0.24, 1.82) | 1.52 (0.29, 7.85) | 0.455 | 0.68 (0.48, 0.98) | 0.66 (0.42, 1.04) | 0.72 (0.39, 1.31) | 0.785 | 0.61 (0.51, 0.72) *** | 0.70 (0.57, 0.87) ** | 0.46 (0.34, 0.63) *** | 0.047 |
| **Total lipids (mmol/l)** | **T2D_all** | **T2D_M** | **T2D_W** | **P-interaction** | **PreDM_all** | **PreDM_M** | **PreDM_W** | **P-interaction** | **Eugly_all** | **Eulgy_M** | **Eugly_W** | **P-interaction** |
| Total lipids in lipoprotein particles | 1.20 (0.50, 2.88) | 0.90 (0.33, 2.51) | 2.75 (0.49, 15.34) | 0.252 | 0.93 (0.63, 1.38) | 0.85 (0.53, 1.38) | 1.02 (0.50, 2.06) | 0.589 | 1.61 (1.32, 1.97) *** | 1.56 (1.23, 1.99) ** | 1.54 (1.05, 2.26) ** | 0.853 |
| Total lipids in VLDL | 1.39 (0.90, 2.14) | 1.17 (0.69, 1.99) | 1.93 (0.86, 4.33) | 0.292 | 1.02 (0.85, 1.22) | 1.01 (0.80, 1.27) | 1.00 (0.74, 1.36) | 0.892 | 1.37 (1.25, 1.50) *** | 1.29 (1.15, 1.44) *** | 1.47 (1.25, 1.72) *** | 0.144 |
| Total lipids in LDL | 1.01 (0.51, 2.00) | 1.01 (0.45, 2.28) | 1.04 (0.29, 3.70) | 0.894 | 1.13 (0.83, 1.55) | 1.07 (0.74, 1.57) | 1.19 (0.69, 2.06) | 0.751 | 1.57 (1.34, 1.83) *** | 1.53 (1.27, 1.84) *** | 1.50 (1.12, 2.01) ** | 0.92 |
| Total lipids in HDL mmol/l | 0.72 (0.27, 1.91) | 0.52 (0.16, 1.68) | 1.95 (0.31, 12.44) | 0.29 | 0.53 (0.35, 0.79) ** | 0.51 (0.31, 0.84) | 0.56 (0.28, 1.12) | 0.742 | 0.51 (0.41, 0.62) *** | 0.55 (0.43, 0.70) *** | 0.43 (0.30, 0.62) *** | 0.326 |
| **Phospholipids (mmol/l)** | **T2D_all** | **T2D_M** | **T2D_W** | **P-interaction** | **PreDM_all** | **PreDM_M** | **PreDM_W** | **P-interaction** | **Eugly_all** | **Eulgy_M** | **Eugly_W** | **P-interaction** |
| Total phospholipids | 1.07 (0.38, 3.01) | 0.76 (0.23, 2.51) | 3.44 (0.42, 28.55) | 0.216 | 0.80 (0.50, 1.27) | 0.72 (0.41, 1.26) | 0.90 (0.39, 2.08) | 0.564 | 1.32 (1.03, 1.67) ** | 1.32 (0.99, 1.75) * | 1.16 (0.73, 1.83) | 0.869 |
| Phospholipids in VLDL | 1.38 (0.88, 2.17) | 1.22 (0.70, 2.14) | 1.71 (0.76, 3.85) | 0.477 | 1.08 (0.89, 1.31) | 1.05 (0.82, 1.35) | 1.08 (0.78, 1.49) | 0.792 | 1.43 (1.30, 1.58) *** | 1.36 (1.20, 1.53) *** | 1.51 (1.28, 1.79) *** | 0.227 |
| Phospholipids in LDL | 0.97 (0.47, 2.02) | 1.01 (0.42, 2.41) | 0.92 (0.24, 3.53) | 0.992 | 1.19 (0.86, 1.65) | 1.12 (0.75, 1.67) | 1.28 (0.72, 2.26) | 0.708 | 1.61 (1.37, 1.90) *** | 1.57 (1.29, 1.91) *** | 1.54 (1.14, 2.09) ** | 0.89 |
| Phospholipids in HDL | 0.80 (0.30, 2.12) | 0.54 (0.17, 1.70) | 2.70 (0.41, 17.70) | 0.185 | 0.51 (0.34, 0.77) ** | 0.50 (0.30, 0.83) | 0.53 (0.26, 1.08) | 0.817 | 0.52 (0.42, 0.64) *** | 0.56 (0.44, 0.71) *** | 0.47 (0.32, 0.67) *** | 0.52 |
| **Apolipoproteins (g/l)** | **T2D_all** | **T2D_M** | **T2D_W** | **P-interaction** | **PreDM_all** | **PreDM_M** | **PreDM_W** | **P-interaction** | **Eugly_all** | **Eulgy_M** | **Eugly_W** | **P-interaction** |
| Apolipoprotein B | 1.17 (0.55, 2.48) | 1.30 (0.53, 3.23) | 0.95 (0.26, 3.53) | 0.777 | 1.33 (0.96, 1.86) | 1.22 (0.81, 1.84) | 1.47 (0.83, 2.59) | 0.571 | 1.82 (1.55, 2.14) *** | 1.81 (1.48, 2.20) *** | 1.68 (1.25, 2.25) ** | 0.843 |
| Apolipoprotein A1 | 0.63 (0.19, 2.08) | 0.40 (0.10, 1.64) | 2.57 (0.27, 24.73) | 0.207 | 0.42 (0.26, 0.70) ** | 0.40 (0.22, 0.74) | 0.46 (0.20, 1.07) | 0.748 | 0.46 (0.36, 0.60) *** | 0.48 (0.36, 0.65) *** | 0.43 (0.28, 0.67) ** | 0.797 |
| Apo(B) to apo(A1) ratio | 1.34 (0.67, 2.67) | 1.81 (0.76, 4.34) | 0.77 (0.25, 2.35) | 0.303 | 1.68 (1.25, 2.24) ** | 1.63 (1.12, 2.35) | 1.69 (1.05, 2.74) | 0.895 | 1.99 (1.73, 2.30) *** | 2.01 (1.69, 2.40) *** | 1.82 (1.43, 2.32) *** | 0.593 |
| **Lipoprotein particle size (nm)** | **T2D_all** | **T2D_M** | **T2D_W** | **P-interaction** | **PreDM_all** | **PreDM_M** | **PreDM_W** | **P-interaction** | **Eugly_all** | **Eulgy_M** | **Eugly_W** | **P-interaction** |
| Average diameter for VLDL particles | 67.58 (0.29, 1.58e+04) | 2.28 (2.69e-03, 1.93e+03) | 4.97e+04 (1.44, 1.71e+09) | 0.129 | 0.13 (0.01, 1.40) | 0.25 (0.01, 4.66) | 0.03 (0.00, 1.64) | 0.492 | 7.48 (2.30, 24.28) ** | 2.66 (0.65, 10.95) | 40.64 (4.81, 343.15) ** | 0.03 |
| Average diameter for LDL particles | 8.29e-06 (1.38e-25, 4.97e+14) | 8.92e+06 (5.75e-18, 1.38e+31) | 9.47e-27 (3.11e-62, 2.88e+09) | 0.156 | 0.17 (7.43e-10, 3.71e+07) | 1.02e+03 (7.93e-08, 1.30e+13) | 2.69e-09 (4.96e-24, 1.46e+06) | 0.15 | 6.83e+02 (3.97e-02, 1.17e+07) | 4.19e+03 (4.65e-02, 3.78e+08) | 1.74 (1.26e-08, 2.38e+08) | 0.457 |
| Average diameter for HDL particles | 0.12 (1.19e-06, 1.22e+04) | 3.56 (1.97e-06, 6.44e+06) | 4.41e-03 (8.86e-12, 2.20e+06) | 0.515 | 0.06 (0.00, 4.85) | 0.03 (0.00, 10.71) | 0.27 (0.00, 260.30) | 0.582 | 2.18e-04 (2.67e-05, 1.78e-03) *** | 4.03e-03 (2.90e-04, 5.60e-02) *** | 5.50e-06 (1.71e-07, 1.77e-04) *** | 0.004 |
| Total concentration of lipoprotein particles | 0.60 (0.18, 1.95) | 0.38 (0.10, 1.55) | 2.36 (0.24, 23.64) | 0.209 | 0.47 (0.28, 0.78) | 0.45 (0.24, 0.83) | 0.49 (0.20, 1.21) | 0.829 | 0.66 (0.51, 0.85) ** | 0.63 (0.46, 0.86) ** | 0.65 (0.40, 1.05) | 0.769 |
| Concentration of VLDL particles | 1.43 (0.81, 2.53) | 1.30 (0.64, 2.62) | 1.68 (0.62, 4.57) | 0.62 | 1.16 (0.91, 1.49) | 1.10 (0.80, 1.50) | 1.22 (0.80, 1.84) | 0.596 | 1.63 (1.44, 1.84) *** | 1.54 (1.33, 1.80) *** | 1.70 (1.37, 2.11) *** | 0.373 |
| Concentration of LDL particles | 1.16 (0.55, 2.43) | 1.28 (0.52, 3.16) | 0.95 (0.26, 3.45) | 0.776 | 1.30 (0.93, 1.80) | 1.21 (0.80, 1.82) | 1.39 (0.79, 2.45) | 0.663 | 1.78 (1.51, 2.10) *** | 1.79 (1.47, 2.18) *** | 1.60 (1.19, 2.15) ** | 0.699 |
| Concentration of HDL particles | 0.58 (0.19, 1.79) | 0.36 (0.09, 1.39) | 2.30 (0.26, 20.41) | 0.186 | 0.44 (0.27, 0.71) ** | 0.43 (0.24, 0.77) | 0.44 (0.19, 1.04) | 0.907 | 0.55 (0.43, 0.71) *** | 0.54 (0.40, 0.72) *** | 0.56 (0.36, 0.88) ** | 0.738 |
| **Lipoprotein size** | **T2D_all** | **T2D_M** | **T2D_W** | **P-interaction** | **PreDM_all** | **PreDM_M** | **PreDM_W** | **P-interaction** | **Eugly_all** | **Eulgy_M** | **Eugly_W** | **P-interaction** |
| XXL_VLDL | 1.20 (0.95, 1.51) | 1.12 (0.84, 1.48) | 1.34 (0.85, 2.11) | 0.518 | 0.99 (0.91, 1.07) | 0.98 (0.88, 1.09) | 0.98 (0.86, 1.12) | 0.808 | 1.12 (1.07, 1.16) *** | 1.07 (1.01, 1.12) ** | 1.18 (1.10, 1.26) *** | 0.018 |
| XL_VLDL | 1.28 (0.93, 1.77) | 1.21 (0.79, 1.85) | 1.34 (0.78, 2.30) | 0.731 | 1.02 (0.90, 1.16) | 1.05 (0.89, 1.25) | 0.97 (0.81, 1.17) | 0.61 | 1.23 (1.16, 1.31) *** | 1.18 (1.09, 1.28) *** | 1.29 (1.16, 1.43) *** | 0.144 |
| L_VLDL | 1.30 (0.90, 1.86) | 1.20 (0.74, 1.95) | 1.38 (0.76, 2.52) | 0.681 | 1.03 (0.89, 1.19) | 1.06 (0.87, 1.29) | 0.97 (0.78, 1.20) | 0.66 | 1.27 (1.19, 1.37) *** | 1.21 (1.11, 1.32) *** | 1.33 (1.19, 1.50) *** | 0.15 |
| M_VLDL | 1.07 (0.75, 1.53) | 1.21 (0.79, 1.87) | 0.83 (0.44, 1.56) | 0.362 | 1.11 (0.93, 1.34) | 1.07 (0.85, 1.33) | 1.18 (0.85, 1.65) | 0.572 | 1.30 (1.18, 1.43) *** | 1.28 (1.15, 1.44) *** | 1.25 (1.05, 1.49) ** | 0.987 |
| S_VLDL | 1.23 (0.72, 2.08) | 1.30 (0.66, 2.55) | 1.10 (0.46, 2.61) | 0.856 | 1.25 (0.99, 1.58) | 1.18 (0.87, 1.59) | 1.32 (0.89, 1.95) | 0.582 | 1.59 (1.42, 1.78) *** | 1.51 (1.31, 1.74) *** | 1.63 (1.34, 1.99) *** | 0.427 |
| XS_VLDL | 1.16 (0.61, 2.18) | 1.29 (0.61, 2.72) | 0.89 (0.26, 3.00) | 0.674 | 1.52 (1.13, 2.05) ** | 1.34 (0.93, 1.91) | 1.89 (1.11, 3.20) | 0.243 | 1.66 (1.43, 1.92) *** | 1.65 (1.38, 1.97) *** | 1.56 (1.19, 2.04) ** | 0.94 |
| IDL | 0.85 (0.45, 1.63) | 0.99 (0.48, 2.07) | 0.54 (0.14, 2.13) | 0.497 | 1.25 (0.93, 1.69) | 1.16 (0.81, 1.66) | 1.41 (0.82, 2.42) | 0.544 | 1.33 (1.14, 1.55) *** | 1.33 (1.11, 1.60) ** | 1.20 (0.89, 1.61) | 0.759 |
| L_LDL | 0.95 (0.50, 1.79) | 0.98 (0.46, 2.06) | 0.91 (0.27, 3.07) | 0.983 | 1.10 (0.85, 1.42) | 1.08 (0.82, 1.43) | 1.11 (0.65, 1.90) | 0.904 | 1.41 (1.22, 1.64) *** | 1.40 (1.17, 1.67) ** | 1.31 (0.99, 1.75) * | 0.928 |
| M_LDL | 1.05 (0.59, 1.88) | 1.04 (0.51, 2.10) | 1.07 (0.38, 3.06) | 0.884 | 1.06 (0.81, 1.37) | 1.00 (0.73, 1.39) | 1.10 (0.69, 1.75) | 0.72 | 1.47 (1.29, 1.68) *** | 1.42 (1.21, 1.66) *** | 1.45 (1.14, 1.86) ** | 0.694 |
| S_LDL | 1.10 (0.55, 2.18) | 1.16 (0.50, 2.67) | 0.99 (0.30, 3.31) | 0.91 | 1.14 (0.84, 1.56) | 1.07 (0.73, 1.57) | 1.21 (0.71, 2.06) | 0.705 | 1.60 (1.38, 1.87) *** | 1.60 (1.33, 1.93) *** | 1.46 (1.11, 1.94) ** | 0.763 |
| XL_HDL | 1.07 (0.70, 1.65) | 1.37 (0.80, 2.33) | 0.68 (0.30, 1.51) | 0.142 | 1.12 (0.92, 1.36) | 1.15 (0.90, 1.46) | 1.09 (0.79, 1.51) | 0.84 | 0.82 (0.75, 0.90) *** | 0.97 (0.86, 1.09) | 0.62 (0.53, 0.72) *** | < 0.001 |
| L_HDL | 0.95 (0.71, 1.27) | 1.00 (0.71, 1.42) | 0.88 (0.49, 1.57) | 0.631 | 0.97 (0.86, 1.10) | 0.99 (0.85, 1.14) | 0.93 (0.73, 1.19) | 0.708 | 0.81 (0.77, 0.86) *** | 0.86 (0.81, 0.92) *** | 0.67 (0.59, 0.75) *** | < 0.001 |
| M_HDL | 0.65 (0.30, 1.41) | 0.51 (0.20, 1.27) | 1.41 (0.32, 6.17) | 0.305 | 0.56 (0.41, 0.77) ** | 0.56 (0.38, 0.83) | 0.56 (0.32, 0.98) | 0.966 | 0.72 (0.67, 0.77) *** | 0.74 (0.68, 0.80) *** | 0.51 (0.38, 0.68) *** | 0.024 |
| S_HDL | 0.54 (0.16, 1.84) | 0.28 (0.07, 1.22) | 2.62 (0.27, 25.37) | 0.109 | 0.48 (0.29, 0.82) ** | 0.47 (0.25, 0.90) | 0.45 (0.18, 1.14) | 0.909 | 0.93 (0.72, 1.22) | 0.68 (0.49, 0.94) *** | 1.55 (0.96, 2.50) | 0.004 |
| **Other lipids (mmol/L)** | **T2D_all** | **T2D_M** | **T2D_W** | **P-interaction** | **PreDM_all** | **PreDM_M** | **PreDM_W** | **P-interaction** | **Eugly_all** | **Eulgy_M** | **Eugly_W** | **P-interaction** |
| Phosphoglycerides | 0.99 (0.39, 2.52) | 0.67 (0.23, 1.98) | 3.58 (0.52, 24.49) | 0.136 | 0.69 (0.45, 1.06) | 0.66 (0.40, 1.10) | 0.70 (0.32, 1.52) | 0.791 | 1.10 (0.88, 1.37) | 1.08 (0.83, 1.40) | 1.05 (0.69, 1.60) | 0.85 |
| TG to phosphoglycerides ratio | 1.71 (1.00, 2.94) | 1.35 (0.70, 2.63) | 2.71 (0.96, 7.62) | 0.268 | 1.04 (0.85, 1.28) | 1.04 (0.80, 1.35) | 1.00 (0.71, 1.40) | 0.969 | 1.40 (1.26, 1.54) *** | 1.28 (1.14, 1.45) *** | 1.59 (1.33, 1.90) *** | 0.043 |
| Total cholines | 0.93 (0.32, 2.69) | 0.65 (0.19, 2.21) | 3.21 (0.38, 27.40) | 0.205 | 0.70 (0.44, 1.12) | 0.65 (0.37, 1.15) | 0.74 (0.32, 1.72) | 0.725 | 1.09 (0.86, 1.39) | 1.10 (0.82, 1.47) | 0.97 (0.61, 1.53) | 0.865 |
| Phosphatidylcholines | 1.05 (0.41, 2.67) | 0.78 (0.26, 2.28) | 2.83 (0.42, 19.15) | 0.24 | 0.67 (0.44, 1.02) | 0.64 (0.38, 1.07) | 0.68 (0.32, 1.45) | 0.771 | 1.04 (0.84, 1.30) | 1.03 (0.80, 1.34) | 0.97 (0.65, 1.47) | 0.967 |
| Sphingomyelins | 0.66 (0.21, 2.07) | 0.67 (0.18, 2.51) | 0.81 (0.08, 8.24) | 0.865 | 1.17 (0.71, 1.91) | 1.02 (0.56, 1.85) | 1.47 (0.61, 3.51) | 0.458 | 1.31 (1.02, 1.68) ** | 1.42 (1.06, 1.91) ** | 0.95 (0.59, 1.52) | 0.26 |
| **Amino acids (mmol/L)** | **T2D_all** | **T2D_M** | **T2D_W** | **P-interaction** | **PreDM_all** | **PreDM_M** | **PreDM_W** | **P-interaction** | **Eugly_all** | **Eulgy_M** | **Eugly_W** | **P-interaction** |
| Alanine | 1.03 (0.53, 1.99) | 0.81 (0.37, 1.78) | 2.07 (0.56, 7.69) | 0.241 | 0.98 (0.73, 1.30) | 1.11 (0.78, 1.58) | 0.74 (0.45, 1.22) | 0.226 | 1.06 (0.92, 1.21) | 1.02 (0.86, 1.21) | 1.12 (0.87, 1.43) | 0.52 |
| Glutamine | 1.79 (0.63, 5.13) | 2.05 (0.58, 7.28) | 2.45 (0.36, 16.84) | 0.921 | 1.48 (0.93, 2.37) | 1.49 (0.84, 2.65) | 1.48 (0.64, 3.42) | 0.978 | 0.99 (0.78, 1.25) | 1.09 (0.82, 1.45) | 0.71 (0.46, 1.08) | 0.141 |
| Glycine | 0.82 (0.57, 1.19) | 0.93 (0.59, 1.47) | 0.74 (0.36, 1.52) | 0.528 | 1.03 (0.85, 1.26) | 1.08 (0.83, 1.40) | 1.02 (0.75, 1.37) | 0.725 | 0.81 (0.74, 0.89) *** | 0.78 (0.70, 0.87) *** | 0.88 (0.76, 1.03) | 0.189 |
| Histidine | 1.85 (0.65, 5.32) | 1.46 (0.40, 5.25) | 5.29 (0.65, 42.87) | 0.414 | 0.66 (0.43, 1.01) | 0.68 (0.40, 1.16) | 0.66 (0.32, 1.38) | 0.949 | 0.93 (0.74, 1.16) | 1.00 (0.76, 1.31) | 0.78 (0.52, 1.17) | 0.323 |
| **Aromatic amino acides (mmol/L)** | **T2D_all** | **T2D_M** | **T2D_W** | **P-interaction** | **PreDM_all** | **PreDM_M** | **PreDM_W** | **P-interaction** | **Eugly_all** | **Eulgy_M** | **Eugly_W** | **P-interaction** |
| Phenylalanine | 1.06 (0.47, 2.36) | 0.80 (0.31, 2.10) | 1.97 (0.44, 8.86) | 0.27 | 1.65 (1.19, 2.29) ** | 1.64 (1.09, 2.47) | 1.72 (1.00, 2.96) | 0.854 | 1.29 (1.11, 1.51) ** | 1.29 (1.06, 1.56) ** | 1.32 (1.00, 1.73) * | 0.879 |
| Tyrosine | 0.73 (0.33, 1.59) | 0.46 (0.18, 1.16) | 2.45 (0.50, 11.90) | 0.065 | 1.07 (0.77, 1.49) | 1.03 (0.68, 1.55) | 1.18 (0.68, 2.05) | 0.66 | 1.29 (1.09, 1.52) ** | 1.32 (1.08, 1.62) ** | 1.20 (0.91, 1.59) | 0.739 |
| **Branched-chain amino acids (BCAA) (mmol/L)** | **T2D_all** | **T2D_M** | **T2D_W** | **P-interaction** | **PreDM_all** | **PreDM_M** | **PreDM_W** | **P-interaction** | **Eugly_all** | **Eulgy_M** | **Eugly_W** | **P-interaction** |
| Isoleucine | 1.26 (0.73, 2.17) | 1.25 (0.65, 2.41) | 1.24 (0.41, 3.69) | 0.906 | 0.91 (0.73, 1.14) | 0.85 (0.64, 1.13) | 1.03 (0.71, 1.50) | 0.437 | 1.23 (1.10, 1.37) *** | 1.23 (1.08, 1.42) ** | 1.18 (0.98, 1.42) | 0.729 |
| Leucine | 1.35 (0.69, 2.65) | 1.42 (0.63, 3.19) | 1.26 (0.34, 4.71) | 0.972 | 0.93 (0.70, 1.23) | 0.77 (0.54, 1.11) | 1.25 (0.78, 1.98) | 0.124 | 1.22 (1.06, 1.41) ** | 1.14 (0.96, 1.37) | 1.31 (1.03, 1.66) ** | 0.379 |
| Valine | 0.99 (0.41, 2.42) | 0.73 (0.24, 2.18) | 2.09 (0.39, 11.28) | 0.256 | 0.77 (0.52, 1.12) | 0.70 (0.43, 1.12) | 0.90 (0.48, 1.70) | 0.498 | 1.20 (0.99, 1.45) ** | 1.14 (0.91, 1.44) | 1.23 (0.89, 1.69) | 0.658 |
| Total BCAA | 1.19 (0.55, 2.61) | 1.06 (0.41, 2.76) | 1.65 (0.36, 7.52) | 0.549 | 0.83 (0.60, 1.17) | 0.73 (0.48, 1.12) | 1.05 (0.60, 1.82) | 0.311 | 1.25 (1.06, 1.47) ** | 1.19 (0.97, 1.47) | 1.27 (0.96, 1.68) | 0.673 |
| **Ketones (mmol/L)** | **T2D_all** | **T2D_M** | **T2D_W** | **P-interaction** | **PreDM_all** | **PreDM_M** | **PreDM_W** | **P-interaction** | **Eugly_all** | **Eulgy_M** | **Eugly_W** | **P-interaction** |
| 3-hydroxybutyrate | 0.89 (0.67, 1.17) | 1.12 (0.76, 1.64) | 0.68 (0.48, 0.95) | 0.054 | 1.01 (0.90, 1.13) | 0.91 (0.79, 1.05) | 1.18 (0.99, 1.40) | 0.02 | 1.01 (0.97, 1.06) | 1.03 (0.97, 1.09) | 0.99 (0.92, 1.07) | 0.605 |
| Acetoacetate | 0.98 (0.76, 1.25) | 1.09 (0.81, 1.47) | 0.74 (0.46, 1.17) | 0.158 | 1.00 (0.91, 1.10) | 0.97 (0.86, 1.08) | 1.08 (0.92, 1.28) | 0.245 | 1.04 (0.99, 1.08) | 1.05 (0.99, 1.10) | 1.02 (0.94, 1.10) | 0.605 |
| Acetone | 1.09 (0.56, 2.11) | 2.27 (1.06, 4.86) | 0.11 (0.02, 0.56) | 0.001 | 0.83 (0.64, 1.07) | 0.68 (0.49, 0.93) | 1.23 (0.80, 1.90) | 0.022 | 0.95 (0.84, 1.07) | 0.99 (0.86, 1.14) | 0.85 (0.69, 1.06) | 0.294 |
| Acetate | 0.75 (0.53, 1.05) | 0.69 (0.46, 1.01) | 1.13 (0.53, 2.43) | 0.257 | 0.94 (0.81, 1.08) | 0.91 (0.76, 1.09) | 1.01 (0.79, 1.30) | 0.549 | 0.97 (0.91, 1.04) | 1.02 (0.94, 1.11) | 0.87 (0.77, 0.99) * | 0.054 |
| **Inflammation (mmol/l)** | **T2D_all** | **T2D_M** | **T2D_W** | **P-interaction** | **PreDM_all** | **PreDM_M** | **PreDM_W** | **P-interaction** | **Eugly_all** | **Eulgy_M** | **Eugly_W** | **P-interaction** |
| Glycoprotein acetyls (GlycA) | 0.89 (0.25, 3.19) | 0.42 (0.09, 1.93) | 4.40 (0.36, 53.85) | 0.081 | 2.15 (1.27, 3.66) ** | 2.08 (1.08, 3.97) | 2.20 (0.87, 5.58) | 0.84 | 3.04 (2.35, 3.93) *** | 2.49 (1.83, 3.37) *** | 4.62 (2.85, 7.49) *** | 0.025 |
| **Renal function (mmol/l )** | **T2D_all** | **T2D_M** | **T2D_W** | **P-interaction** | **PreDM_all** | **PreDM_M** | **PreDM_W** | **P-interaction** | **Eugly_all** | **Eulgy_M** | **Eugly_W** | **P-interaction** |
| Creatinine | 1.79 (0.69, 4.69) | 2.42 (0.74, 7.87) | 1.08 (0.17, 6.73) | 0.437 | 1.41 (0.92, 2.16) | 0.95 (0.55, 1.64) | 2.68 (1.38, 5.20) | 0.014 | 1.61 (1.30, 1.99) *** | 1.53 (1.18, 1.98) ** | 1.62 (1.12, 2.35) ** | 0.736 |
| Albumin | 0.64 (0.09, 4.41) | 0.11 (0.01, 1.18) | 37.49 (1.07, 1318.54) | 0.01 | 0.28 (0.13, 0.62) ** | 0.32 (0.12, 0.84) | 0.23 (0.06, 0.90) | 0.705 | 0.39 (0.26, 0.59) *** | 0.41 (0.25, 0.68) ** | 0.29 (0.14, 0.63) ** | 0.472 |
| **Fatty acids (mmol/l)** | **T2D_all** | **T2D_M** | **T2D_W** | **P-interaction** | **PreDM_all** | **PreDM_M** | **PreDM_W** | **P-interaction** | **Eugly_all** | **Eulgy_M** | **Eugly_W** | **P-interaction** |
| Total fatty acids | 1.34 (0.61, 2.95) | 0.91 (0.36, 2.29) | 4.28 (0.86, 21.32) | 0.094 | 0.89 (0.62, 1.27) | 0.84 (0.55, 1.30) | 0.90 (0.47, 1.73) | 0.757 | 1.62 (1.35, 1.94) *** | 1.50 (1.21, 1.86) ** | 1.81 (1.27, 2.58) ***\ | 0.267 |
| Omega-3 fatty acids | 0.88 (0.58, 1.34) | 0.78 (0.47, 1.28) | 1.22 (0.51, 2.90) | 0.452 | 0.80 (0.67, 0.95) * | 0.82 (0.66, 1.00) | 0.72 (0.52, 0.99) | 0.634 | 0.98 (0.90, 1.07) | 0.99 (0.90, 1.10) | 0.92 (0.78, 1.08) | 0.71 |
| Omega-6 fatty acids | 1.13 (0.37, 3.48) | 1.04 (0.28, 3.84) | 1.72 (0.19, 15.80) | 0.687 | 0.86 (0.53, 1.41) | 0.83 (0.46, 1.51) | 0.83 (0.35, 1.99) | 0.995 | 1.60 (1.25, 2.05) *** | 1.63 (1.22, 2.19) ** | 1.32 (0.83, 2.10) | 0.595 |
| Polyunsaturated fatty acids (PUFA) | 0.96 (0.34, 2.74) | 0.83 (0.24, 2.80) | 1.72 (0.21, 14.01) | 0.563 | 0.76 (0.48, 1.21) | 0.76 (0.43, 1.32) | 0.68 (0.30, 1.56) | 0.856 | 1.42 (1.13, 1.79) ** | 1.45 (1.10, 1.91) ** | 1.17 (0.75, 1.83) | 0.633 |
| Monounsaturated fatty acids (MUFA) | 1.42 (0.81, 2.51) | 1.05 (0.54, 2.04) | 3.32 (1.02, 10.84) | 0.088 | 0.95 (0.73, 1.23) | 0.91 (0.67, 1.25) | 0.97 (0.60, 1.55) | 0.696 | 1.53 (1.34, 1.74) *** | 1.38 (1.19, 1.61) *** | 1.87 (1.45, 2.40) *** | 0.029 |
| Saturated fatty acids (SFA) | 1.37 (0.69, 2.71) | 0.89 (0.40, 1.99) | 5.17 (1.22, 21.85) | 0.033 | 0.95 (0.69, 1.29) | 0.88 (0.61, 1.28) | 1.02 (0.58, 1.80) | 0.54 | 1.53 (1.31, 1.80) *** | 1.40 (1.16, 1.69) ** | 1.80 (1.32, 2.46) ** | 0.12 |
| Linoleic acid | 1.19 (0.52, 2.74) | 1.22 (0.46, 3.22) | 1.22 (0.24, 6.24) | 0.948 | 0.92 (0.64, 1.33) | 0.92 (0.58, 1.44) | 0.86 (0.45, 1.66) | 0.862 | 1.37 (1.13, 1.65) ** | 1.39 (1.12, 1.74) ** | 1.16 (0.82, 1.66) | 0.534 |
| Docosahexaenoic acid (DHA) | 0.67 (0.41, 1.11) | 0.60 (0.34, 1.09) | 0.91 (0.32, 2.55) | 0.568 | 0.80 (0.66, 0.98) | 0.83 (0.66, 1.05) | 0.72 (0.49, 1.04) | 0.57 | 0.88 (0.80, 0.98) ** | 0.93 (0.83, 1.05) | 0.74 (0.60, 0.91) ** | 0.135 |
| **Fatty acid ratio (%)** | **T2D_all** | **T2D_M** | **T2D_W** | **P-interaction** | **PreDM_all** | **PreDM_M** | **PreDM_W** | **P-interaction** | **Eugly_all** | **Eulgy_M** | **Eugly_W** | **P-interaction** |
| Omega-3 fatty acids to total fatty acids | 0.71 (0.42, 1.21) | 0.69 (0.37, 1.31) | 0.70 (0.24, 2.09) | 0.899 | 0.74 (0.60, 0.92) ** | 0.78 (0.61, 1.01) | 0.64 (0.44, 0.95) | 0.483 | 0.83 (0.75, 0.92) ** | 0.87 (0.77, 0.98) ** | 0.75 (0.62, 0.90) ** | 0.338 |
| Omega-6 fatty acids to total fatty acids | 0.41 (0.09, 1.87) | 1.53 (0.25, 9.44) | 0.01 (6.00e-04, 0.26) | 0.008 | 1.15 (0.59, 2.28) | 1.29 (0.58, 2.88) | 1.01 (0.28, 3.65) | 0.544 | 0.43 (0.30, 0.61) *** | 0.60 (0.40, 0.90) ** | 0.17 (0.09, 0.34) *** | 0.001 |
| PUFA to total fatty acids | 0.23 (0.04, 1.23) | 0.93 (0.13, 6.60) | 2.67e-03 (6.42e-05, 0.11) | 0.007 | 0.83 (0.40, 1.72) | 1.02 (0.43, 2.45) | 0.55 (0.14, 2.15) | 0.311 | 0.31 (0.21, 0.45) *** | 0.47 (0.30, 0.72) ** | 0.09 (0.04, 0.20) *** | < 0.001 |
| MUFA to total fatty acids | 4.73 (0.94, 23.69) | 2.13 (0.32, 14.09) | 29.53 (1.04, 837.44) | 0.18 | 1.06 (0.53, 2.11) | 0.98 (0.43, 2.26) | 1.17 (0.35, 3.95) | 0.665 | 3.32 (2.37, 4.65) *** | 2.23 (1.50, 3.32) *** | 8.12 (4.29, 15.38) *** | 0.001 |
| Saturated fatty acids to total fatty acids | 4.84 (0.26, 88.66) | 0.44 (0.01, 13.94) | 6.35e+03 (1.12e+01, 3.62e+06) | 0.01 | 1.64 (0.50, 5.35) | 0.99 (0.24, 4.04) | 4.31 (0.48, 38.66) | 0.166 | 2.50 (1.35, 4.62) ** | 1.68 (0.82, 3.44) | 7.42 (2.17, 25.38) ** | 0.034 |
| Linoleic acid to total fatty acids | 0.65 (0.16, 2.60) | 2.38 (0.45, 12.66) | 0.03 (1.97e-03, 0.42) | 0.008 | 1.12 (0.62, 2.02) | 1.28 (0.63, 2.58) | 0.89 (0.30, 2.66) | 0.418 | 0.59 (0.44, 0.80) ** | 0.77 (0.54, 1.10) | 0.29 (0.16, 0.53) *** | 0.004 |
| DHA to total fatty acids | 0.60 (0.36, 0.98) | 0.64 (0.36, 1.13) | 0.49 (0.18, 1.39) | 0.597 | 0.84 (0.69, 1.02) | 0.88 (0.70, 1.11) | 0.74 (0.51, 1.08) | 0.459 | 0.76 (0.69, 0.84) *** | 0.83 (0.74, 0.93) ** | 0.59 (0.48, 0.73) *** | 0.011 |
| PUFA to MUFA ratio | 0.44 (0.19, 1.04) | 0.80 (0.30, 2.14) | 0.08 (0.01, 0.54) | 0.038 | 0.94 (0.65, 1.35) | 1.01 (0.65, 1.57) | 0.83 (0.42, 1.62) | 0.465 | 0.52 (0.43, 0.63) *** | 0.65 (0.53, 0.81) *** | 0.30 (0.21, 0.43) *** | < 0.001 |
| Omega-6 fatty acids to omega-3 fatty acids | 1.21 (0.75, 1.97) | 1.42 (0.80, 2.53) | 0.86 (0.32, 2.29) | 0.476 | 1.30 (1.07, 1.58) ** | 1.26 (1.00, 1.59) | 1.46 (1.02, 2.10) | 0.631 | 1.10 (1.00, 1.21) * | 1.09 (0.97, 1.22) | 1.15 (0.97, 1.38) | 0.838 |
| Degree of unsaturation | 0.04 (0.00, 0.77) | 0.23 (0.01, 6.37) | 1.93e-04 (2.18e-07, 0.17) | 0.063 | 0.44 (0.13, 1.46) | 0.62 (0.14, 2.69) | 0.20 (0.02, 1.74) | 0.297 | 0.19 (0.10, 0.36) *** | 0.34 (0.17, 0.70) ** | 0.04 (0.01, 0.14) *** | 0.005 |

FDR-adjusted P*<.1; ** <.05; ***<.0001

Fully adjusted for age, race, income, area deprivation, smoking, alcohol drinking, physical activity, BMI, and medications for hypertension, hyperlipidemia, and diabetes, and additionally adjusted for menopausal status for women.
