## Supplemental Table 4 for "Sex differences in circulating metabolites across glycemic status and risk of coronary heart disease"

**Supplemental Table 4.** **Adjusted β Coefficients (95% CI) for Sex Differences of Metabolites Across Glycemic Status, Excluding Those with Lipid-Lowering Medications**

|  | **Sex Difference** |  |  |
| --- | --- | --- | --- |
|  | **Euglycemia** | **Pre-DM** | **New T2D** |
| **Cholesterol (mmol/l)** | **β coefficient** | **β coefficient** | **β coefficient** |
| Total-c | 0.37 (0.36, 0.39) *** | 0.51 (0.48, 0.55) *** | 0.53 (0.41, 0.65) *** |
| Non-HDL-c | 0.06 (0.05, 0.08) *** | 0.27 (0.23, 0.31) *** | 0.31 (0.2, 0.42) *** |
| Remnant-c | 0.04 (0.03, 0.04) *** | 0.14 (0.12, 0.16) *** | 0.17 (0.11, 0.22) *** |
| VLDL-c | -0.05 (-0.06, -0.05) *** | 0.02 (0.01, 0.03) *** | 0.05 (0.02, 0.09) ** |
| LDL-c | 0.03 (0.02, 0.03) *** | 0.13 (0.11, 0.15) *** | 0.15 (0.09, 0.2) *** |
| HDL-c | 0.31 (0.31, 0.31) *** | 0.24 (0.23, 0.26) *** | 0.22 (0.18, 0.25) *** |
| **Triglycerides (mmol/l)** |  |  |  |
| Total TG | -0.25 (-0.26, -0.24) *** | -0.13 (-0.16, -0.1) *** | -0.01 (-0.1, 0.08) |
| TG in VLDL | -0.24 (-0.25, -0.24) *** | -0.14 (-0.16, -0.12) *** | -0.04 (-0.12, 0.03) |
| TG in LDL | -0.01 (-0.01, -0.01) *** | -1.24e-05 (-1.89e-03, 1.87e-03) | 0.01 (0, 0.02) ** |
| TG in HDL | -1.83e-03 (-2.57e-03, -1.10e-03) *** | 3.58e-03 (1.47e-03, 5.70e-03) ** | 0.01 (0, 0.02) ** |
| **Cholesteryl esters (mmol/l)** |  |  |  |
| Total esterified cholesterol | 0.29 (0.28, 0.3) *** | 0.38 (0.36, 0.41) *** | 0.39 (0.3, 0.48) *** |
| Cholesteryl esters in VLDL | -0.02 (-0.02, -0.02) *** | 0.02 (0.02, 0.03) *** | 0.04 (0.02, 0.06) *** |
| Cholesteryl esters in LDL | 7.60e-03 (2.69e-03, 1.25e-02) ** | 0.09 (0.07, 0.1) *** | 0.1 (0.06, 0.15) *** |
| Cholesteryl esters in HDL | 2.39e-01 (2.35e-01, 2.42e-01) *** | 0.19 (0.18, 0.2) *** | 0.17 (0.14, 0.19) *** |
| **Free cholesterol (mmol/l)** |  |  |  |
| Total free cholesterol | 0.08 (0.08, 0.09) *** | 0.13 (0.12, 0.14) *** | 0.14 (0.1, 0.17) *** |
| Free cholesterol in VLDL | -0.03 (-0.04, -0.03) *** | -1.82e-03 (-6.63e-03, 2.98e-03) | 0.01 (0, 0.03) * |
| Free cholesterol in LDL | 0.02 (0.02, 0.02) *** | 0.04 (0.04, 0.05) *** | 0.04 (0.03, 0.06) *** |
| Free cholesterol in HDL | 0.07 (0.07, 0.07) *** | 0.06 (0.05, 0.06) *** | 0.05 (0.04, 0.06) *** |
| **Total lipids (mmol/l)** |  |  |  |
| Total lipids in lipoprotein particles | 0.38 (0.36, 0.41) *** | 0.68 (0.61, 0.75) *** | 0.85 (0.61, 1.09) *** |
| Total lipids in VLDL | -0.36 (-0.38, -0.35) *** | -0.13 (-0.17, -0.09) *** | 0.03 (-0.1, 0.16) |
| Total lipids in LDL | 0.02 (0.02, 0.03) *** | 0.17 (0.14, 0.19) *** | 0.2 (0.12, 0.28) *** |
| Total lipids in HDL | 0.6 (0.59, 0.61) *** | 0.49 (0.46, 0.51) *** | 0.46 (0.39, 0.54) *** |
| **Phospholipids (mmol/l)** |  |  |  |
| Total phospholipids | 0.26 (0.25, 0.27) *** | 0.3 (0.28, 0.32) *** | 0.33 (0.27, 0.4) *** |
| Phospholipids in VLDL | -0.07 (-0.07, -0.06) *** | -0.01 (-0.02, -4.69e-03) ** | 0.02 (-0.01, 0.04) |
| Phospholipids in LDL | 3.06e-03 (9.18e-04, 5.20e-03) ** | 0.04 (0.03, 0.04) *** | 0.04 (0.03, 0.06) *** |
| Phospholipids in HDL | 0.3 (0.29, 0.3) *** | 0.24 (0.23, 0.25) *** | 0.24 (0.2, 0.27) *** |
| **Apolipoproteins (g/l)** |  |  |  |
| Apolipoprotein B | -8.89e-03 (-1.19e-02, -5.84e-03) *** | 0.04 (0.04, 0.05) *** | 0.06 (0.03, 0.08) *** |
| Apolipoprotein A1 | 2.10e-01 (2.07e-01, 2.14e-01) *** | 0.18 (0.17, 0.19) *** | 0.18 (0.15, 0.21) *** |
| Apo(B) to apo(A1) ratio | -9.09e-02 (-9.35e-02, -8.84e-02) *** | -0.04 (-0.05, -0.04) *** | -0.02 (-0.05, 0) * |
| **Lipoprotein particle size (nm)** |  |  |  |
| Average diameter for VLDL particles | -0.97 (-0.99, -0.95) *** | -0.63 (-0.68, -0.58) *** | -0.53 (-0.72, -0.34) *** |
| Average diameter for LDL particles | 3.96e-02 (3.83e-02, 4.10e-02) *** | 0.04 (0.03, 0.04) *** | 0.03 (0.02, 0.05) *** |
| Average diameter for HDL particles | 2.00e-01 (1.98e-01, 2.03e-01) *** | 0.14 (0.13, 0.15) *** | 0.12 (0.1, 0.14) *** |
| **Total concentration of lipoprotein particles** | 1.75e-03 (1.71e-03, 1.79e-03) *** | 1.76e-03 (1.66e-03, 1.86e-03) *** | 1.85e-03 (1.51e-03, 2.19e-03) *** |
| Concentration of VLDL particles | -1.05e-05 (-1.11e-05, -9.78e-06) ****** | 2.20e-06 (2.15e-07, 4.18e-06) ** | 8.89e-06 (2.70e-06, 1.51e-05) ** |
| Concentration of LDL particles | -1.98e-05 (-2.41e-05, -1.55e-05) *** | 5.59e-05 (4.34e-05, 6.84e-05) *** | 6.99e-05 (3.19e-05, 1.08e-04) ** |
| Concentration of HDL particles | 1.77e-03 (1.73e-03, 1.80e-03) *** | 1.67e-03 (1.58e-03, 1.77e-03) *** | 1.74e-03 (1.41e-03, 2.07e-03) *** |
| **Lipoprotein size** |  |  |  |
| XXL_VLDL | -2.27e-02 (-2.33e-02, -2.20e-02) *** | -1.21e-02 (-1.41e-02, -1.02e-02) *** | -3.86e-03 (-1.03e-02, 2.61e-03) |
| XL_VLDL | -0.01 (-0.02, -0.01) *** | -6.01e-03 (-7.27e-03, -4.75e-03) *** | -1.00e-03 (-4.96e-03, 2.95e-03) |
| L_VLDL | -2.38e-02 (-2.46e-02, -2.31e-02) *** | -8.70e-03 (-1.08e-02, -6.64e-03) *** | -1.24e-03 (-7.68e-03, 5.20e-03) |
| M_VLDL | 4.98e-04 (-5.03e-04, 1.50e-03) | 1.86e-02 (1.57e-02, 2.15e-02) *** | 0.02 (0.01, 0.03) *** |
| S_VLDL | -1.05e-02 (-1.13e-02, -9.66e-03) *** | 5.97e-03 (3.70e-03, 8.24e-03) *** | 0.01 (0, 0.02) ** |
| XS_VLDL | 1.76e-02 (1.69e-02, 1.84e-02) *** | 0.03 (0.02, 0.03) *** | 0.03 (0.02, 0.03) *** |
| IDL | 9.01e-02 (8.70e-02, 9.32e-02) *** | 0.12 (0.11, 0.13) *** | 0.12 (0.09, 0.14) *** |
| L_LDL | 4.96e-02 (4.54e-02, 5.39e-02) *** | 0.11 (0.09, 0.12) *** | 0.11 (0.07, 0.15) *** |
| M_LDL | -0.02 (-0.02, -0.01) *** | 0.02 (0.01, 0.02) *** | 0.03 (0.01, 0.04) ** |
| S_LDL | -6.05e-03 (-6.73e-03, -5.37e-03) *** | 6.09e-03 (4.14e-03, 8.05e-03) *** | 8.52e-03 (2.47e-03, 1.46e-02)  ** |
| XL_HDL | 3.00e-02 (2.95e-02, 3.05e-02) *** | 2.03e-02 (1.91e-02, 2.15e-02) *** | 0.01 (0.01, 0.02) *** |
| L_HDL | 1.62e-01 (1.60e-01, 1.64e-01) *** | 0.11 (0.11, 0.12) *** | 0.09 (0.07, 0.1) *** |
| M_HDL | 0.11 (0.1, 0.11) *** | 0.09 (0.08, 0.09) *** | 0.09 (0.07, 0.1) *** |
| S_HDL | 1.10e-02 (1.00e-02, 1.19e-02) *** | 2.14e-02 (1.88e-02, 2.40e-02) *** | 0.03 (0.02, 0.04) *** |
| **Other lipids (mmol/L)** |  |  |  |
| Phosphoglycerides | 0.25 (0.24, 0.25) *** | 0.26 (0.25, 0.28) *** | 0.3 (0.24, 0.35) *** |
| TG to phosphoglycerides ratio | -0.17 (-0.17, -0.17) *** | -0.13 (-0.14, -0.12) *** | -0.1 (-0.14, -0.07) *** |
| Total cholines | 0.27 (0.26, 0.28) *** | 0.29 (0.27, 0.3) *** | 0.31 (0.25, 0.37) *** |
| Phosphatidylcholines | 0.26 (0.25, 0.26) *** | 0.26 (0.25, 0.28) *** | 0.28 (0.23, 0.34) *** |
| Sphingomyelins | 4.71e-02 (4.61e-02, 4.81e-02) *** | 5.05e-02 (4.77e-02, 5.34e-02) *** | 0.05 (0.04, 0.06) *** |
| **Amino acids (mmol/L)** |  |  |  |
| Alanine | -0.02 (-0.02, -0.01) *** | -6.91e-03 (-1.02e-02, -3.66e-03) *** | -2.07e-03 (-1.49e-02, 1.08e-02) |
| Glutamine | -0.02 (-0.02, -0.01) *** | -4.76e-03 (-8.17e-03, -1.35e-03) ** | -8.81e-03 (-2.10e-02, 3.41e-03) |
| Glycine | 4.26e-02 (4.16e-02, 4.36e-02) *** | 3.79e-02 (3.54e-02, 4.03e-02) *** | 0.02 (0.01, 0.03) *** |
| Histidine | -2.13e-03 (-2.29e-03, -1.96e-03) *** | -2.08e-03 (-2.53e-03, -1.62e-03) *** | -1.74e-03 (-3.25e-03, -2.26e-04) ** |
| **Aromatic amino acids (mmol/L)** |  |  |  |
| Phenylalanine | -1.11e-03 (-1.29e-03, -9.40e-04) *** | -1.42e-03 (-1.87e-03, -9.66e-04) *** | -7.19e-04 (-2.24e-03, 8.07e-04) |
| Tyrosine | 1.56e-05 (-2.02e-04, 2.33e-04) | -2.96e-04 (-9.20e-04, 3.27e-04) | -2.80e-04 (-2.50e-03, 1.94e-03) |
| **Branched-chain amino acids (BCAA) (mmol/L)** |  |  |  |
| Isoleucine | -7.08e-03 (-7.36e-03, -6.81e-03) *** | -5.85e-03 (-6.62e-03, -5.08e-03) *** | -4.68e-03 (-7.55e-03, -1.82e-03) ** |
| Leucine | -1.57e-02 (-1.61e-02, -1.53e-02) *** | -1.25e-02 (-1.37e-02, -1.13e-02) *** | -0.01 (-0.02, -0.01) *** |
| Valine | -1.99e-02 (-2.05e-02, -1.92e-02) *** | -1.43e-02 (-1.61e-02, -1.25e-02) *** | -1.13e-02 (-1.78e-02, -4.68e-03) ** |
| Total BCAA | -4.27e-02 (-4.39e-02, -4.14e-02) *** | -3.27e-02 (-3.63e-02, -2.90e-02) *** | -0.03 (-0.04, -0.01) *** |
| **Ketones (mmol/L)** |  |  |  |
| 3-hydroxybutyrate | 4.24e-03 (3.26e-03, 5.22e-03) *** | -6.34e-04 (-2.61e-03, 1.35e-03) | -2.89e-03 (-9.96e-03, 4.19e-03) |
| Acetoacetate | 4.56e-04 (2.58e-04, 6.53e-04) *** | -6.73e-04 (-1.13e-03, -2.15e-04) ** | 9.51e-04 (-6.51e-04, 2.55e-03) |
| Acetone | -4.61e-05 (-1.36e-04, 4.36e-05) | -5.05e-04 (-7.06e-04, -3.04e-04) *** | -2.83e-04 (-9.85e-04, 4.19e-04) |
| Acetate | -5.76e-05 (-2.35e-04, 1.20e-04) | -3.97e-04 (-9.42e-04, 1.47e-04) | -8.92e-04 (-2.47e-03, 6.83e-04) |
| **Inflammation (mmol/l)** |  |  |  |
| Glycoprotein acetyls (GlycA) | -8.11e-03 (-9.83e-03, -6.39e-03) *** | 2.05e-02 (1.56e-02, 2.55e-02) *** | 0.05 (0.04, 0.07) *** |
| **Renal function (mmol/l )** |  |  |  |
| Creatinine | -1.36e-02 (-1.37e-02, -1.34e-02) *** | -1.28e-02 (-1.33e-02, -1.23e-02) *** | -0.01 (-0.01, -0.01) *** |
| Albumin | -0.34 (-0.39, -0.29) *** | -0.04 (-0.18, 0.11) | -0.88 (-1.38, -0.39) ** |
| **Fatty acids (mmol/l)** |  |  |  |
| Total fatty acids | 0.15 (0.11, 0.19) *** | 0.59 (0.48, 0.7) *** | 0.97 (0.59, 1.36) *** |
| Omega-3 fatty acids | 0.06 (0.06, 0.07) *** | 0.07 (0.07, 0.08) *** | 0.08 (0.04, 0.11) *** |
| Omega-6 fatty acids | 0.19 (0.18, 0.2) *** | 0.32 (0.29, 0.35) *** | 0.35 (0.25, 0.45) *** |
| Polyunsaturated fatty acids (PUFA) | 0.25 (0.24, 0.26) *** | 0.4 (0.36, 0.43) *** | 0.42 (0.31, 0.54) *** |
| Monounsaturated fatty acids (MUFA) | -0.14 (-0.16, -0.13) *** | 0.02 (-0.02, 0.06) | 0.18 (0.04, 0.31) ** |
| Saturated fatty acids (SFA) | 0.04 (0.03, 0.05) *** | 0.18 (0.13, 0.22) *** | 0.37 (0.22, 0.53) *** |
| Linoleic acid | 0.13 (0.12, 0.14) *** | 0.27 (0.24, 0.3) *** | 0.27 (0.18, 0.37) *** |
| Docosahexaenoic acid (DHA) | 3.99e-02 (3.86e-02, 4.11e-02) *** | 3.86e-02 (3.50e-02, 4.22e-02) *** | 0.03 (0.02, 0.05) *** |
| **Fatty acid ratio (%)** |  |  |  |
| Omega-3 fatty acids to total fatty acids | 4.70e-03 (4.46e-03, 4.94e-03) *** | 3.97e-03 (3.32e-03, 4.63e-03) *** | 2.46e-03 (2.46e-04, 4.67e-03) ** |
| Omega-6 fatty acids to total fatty acids | 9.84e-03 (9.32e-03, 1.04e-02) *** | 7.45e-03 (5.89e-03, 9.01e-03) *** | -1.41e-03 (-7.16e-03, 4.34e-03) |
| PUFA to total fatty acids | 0.01 (0.01, 0.02) *** | 1.14e-02 (9.82e-03, 1.30e-02) *** | 1.04e-03 (-4.88e-03, 6.97e-03) |
| MUFA to total fatty acids | -1.43e-02 (-1.47e-02, -1.39e-02) *** | -9.81e-03 (-1.09e-02, -8.73e-03) *** | -5.19e-03 (-9.31e-03, -1.06e-03) ** |
| Saturated fatty acids to total fatty acids | -2.59e-04 (-5.55e-04, 3.81e-05) * | -1.61e-03 (-2.50e-03, -7.32e-04) *** | 4.15e-03 (1.02e-03, 7.27e-03) ** |
| Linoleic acid to total fatty acids | 6.64e-03 (6.14e-03, 7.13e-03) *** | 8.01e-03 (6.58e-03, 9.45e-03) *** | 5.05e-04 (-4.43e-03, 5.44e-03) |
| DHA to total fatty acids | 3.01e-03 (2.91e-03, 3.11e-03) *** | 2.14e-03 (1.86e-03, 2.43e-03) *** | 1.14e-03 (1.44e-04, 2.13e-03) ** |
| PUFA to MUFA ratio | 0.17 (0.17, 0.18) *** | 3.97e-03 (3.32e-03, 4.63e-03) *** | 0.03 (-0.02, 0.08) |
| Omega-6 fatty acids to omega-3 fatty acids | -0.86 (-0.93, -0.8) *** | -0.87 (-1.06, -0.68) *** | -0.61 (-1.14, -0.08) ** |
| Degree of unsaturation | 4.32e-02 (4.20e-02, 4.44e-02) *** | 0.04 (0.03, 0.04) *** | 0.02 (0.01, 0.04) *** |

*FDR-adjusted p<.1; ** <.05; ***<.0001

Fully adjusted for age, race, income, area deprivation, smoking, alcohol drinking, physical activity, BMI, and medications for hypertension and diabetes.
