## Supplemental Table 5 for "Sex differences in circulating metabolites across glycemic status and risk of coronary heart disease"

**Supplemental Table 5. Adjusted β Coefficients (95% CI) for Sex Differences of and Adjusted Mean (95% CI) Clinical Chemistry Biomarkers Across Glycemic Status**

|  |  | **Euglycemia** | **Pre-DM** | **New T2D** | **Women** | **Men** | **Women** | **Men** | **Women** | **Men** |
| --- | --- | --- | --- | --- | --- | --- | --- | --- | --- | --- |
|  | **Filed ID** | β coefficient | β coefficient | β coefficient | Adjusted mean | Adjusted mean | Adjusted mean | Adjusted mean | Adjusted mean | Adjusted mean |
| Total-c g/L | 30690 | 0.21 (0.19, 0.23) ^***^ | 0.41 (0.36, 0.46) ^***^ | 0.56 (0.41, 0.70) ^***^ | 5.39, (5.36, 5.42) | 5.18, (5.16, 5.21) | 5.61, (5.55, 5.66) | 5.20, (5.15, 5.25) | 5.23, (5.06, 5.39) | 4.67, (4.53, 4.81) |
| LDL-c g/L | 30780 | -0.03 (-0.04, -0.01) ^***^ | 0.17 (0.13, 0.20) ^***^ | 0.28 (0.17, 0.39) ^***^ | 3.23, (3.21, 3.26) | 3.26, (3.24, 3.28) | 3.45, (3.41, 3.49) | 3.28, (3.24, 3.32) | 3.17, (3.05, 3.30) | 2.90, (2.79, 3.00) |
| HDL-c g/L | 30760 | 0.35 (0.34, 0.35) ^***^ | 0.29 (0.28, 0.31) ^***^ | 0.27 (0.23, 0.31) ^***^ | 1.57, (1.56, 1.58) | 1.23, (1.22, 1.24) | 1.49, (1.47, 1.50) | 1.20, (1.18, 1.21) | 1.37, (1.33, 1.42) | 1.10, (1.06, 1.14) |
| Triglycerides g/L | 30870 | -0.48 (-0.49, -0.46) ^***^ | -0.34 (-0.39, -0.29) ^***^ | -0.15 (-0.33, 0.02) ^*^ | 1.56, (1.53, 1.58) | 2.04, (2.01, 2.06) | 1.83, (1.78, 1.89) | 2.18, (2.13, 2.23) | 2.09, (1.89, 2.29) | 2.24, (2.07, 2.41) |
| Apolipoprotein B g/L | 30640 | -0.03 (-0.03, -0.03) ^***^ | 0.02 (0.01, 0.03) ^**^ | 0.06 (0.03, 0.09) ^***^ | 0.95, (0.95, 0.96) | 0.98, (0.98, 0.99) | 1.01, (1.00, 1.03) | 1.00, (0.99, 1.01) | 0.98, (0.94, 1.01) | 0.92, (0.89, 0.95) |
| Apolipoprotein A g/L | 30630 | 0.23 (0.23, 0.24) ^***^ | 0.21 (0.2, 0.22) ^***^ | 0.21 (0.17, 0.24) ^***^ | 1.64, (1.63, 1.65) | 1.41, (1.40, 1.41) | 1.60, (1.58, 1.61) | 1.38, (1.37, 1.40) | 1.53, (1.49, 1.57) | 1.32, (1.29, 1.36) |
| Albumin g/L | 30600 | -0.6 (-0.65, -0.56) ^***^ | -0.22 (-0.35, -0.1) ^***^ | -0.78 (-1.2, -0.36) ^***^ | 44.99, (44.92, 45.07) | 45.59, (45.52, 45.67) | 44.80, (44.66, 44.93) | 45.02, (44.90, 45.15) | 44.85, (44.37, 45.33) | 45.63, (45.22, 46.04) |

FDR-adjusted p *<.1; ** <.05; ***<.0001

Fully adjusted for age, race, income, area deprivation, smoking, alcohol drinking, physical activity, BMI, and medications for hypertension, hyperlipidemia, and diabetes.
