## Supplemental Table 6 for "Sex differences in circulating metabolites across glycemic status and risk of coronary heart disease"

**Supplemental Table 6. Hazard Ratios (95% CI) of Metabolites and Risk of CHD by Sex Across Glycemic Status, Excluding Lipid-Lowering Medications**

|  | **Euglycemia** |  |  |  | **Pre-DM** |  |  |  | **New T2D** |  |  |  |
| --- | --- | --- | --- | --- | --- | --- | --- | --- | --- | --- | --- | --- |
|  | **All** | **Men** | **Women** | **P-interaction** | **All** | **Men** | **Women** | **P-interaction** | **All** | **Men** | **Women** | **P-interaction** |
| **Cholesterol (mmol/l)** |  |  |  |  |  |  |  |  |  |  |  |  |
| Total-c | 1.03 (0.85, 1.23) | 1.06 (0.85, 1.32) | 0.81 (0.57, 1.15) | 0.332 | 0.69 (0.49, 0.97) | 0.67 (0.44, 1.02) | 0.66 (0.36, 1.21) | 0.99 | 0.91 (0.41, 2.00) | 0.92 (0.37, 2.33) | 0.99 (0.20, 4.77) | 0.896 |
| Non-HDL-c | 1.20 (1.04, 1.38) ^**^ | 1.18 (1.00, 1.40) ^*^ | 1.09 (0.84, 1.42) | 0.818 | 0.81 (0.62, 1.06) | 0.80 (0.57, 1.11) | 0.76 (0.48, 1.22) | 0.911 | 1.04 (0.56, 1.93) | 1.08 (0.51, 2.26) | 1.01 (0.32, 3.19) | 0.981 |
| Remnant-c | 1.21 (1.06, 1.39) ^**^ | 1.19 (1.01, 1.40) ^*^ | 1.13 (0.88, 1.46) | 0.962 | 0.83 (0.64, 1.08) | 0.80 (0.59, 1.11) | 0.82 (0.52, 1.28) | 0.903 | 1.11 (0.61, 2.00) | 1.15 (0.57, 2.33) | 1.06 (0.35, 3.17) | 0.985 |
| VLDL-c | 1.27 (1.15, 1.41) ^***^ | 1.23 (1.08, 1.40) ^**^ | 1.28 (1.06, 1.54) ^**^ | 0.572 | 0.90 (0.73, 1.10) | 0.88 (0.68, 1.14) | 0.88 (0.63, 1.23) | 0.913 | 1.28 (0.79, 2.08) | 1.26 (0.69, 2.31) | 1.31 (0.58, 2.97) | 0.871 |
| LDL-c | 1.16 (1.01, 1.33) ^*^ | 1.16 (0.98, 1.37) | 1.04 (0.79, 1.35) | 0.67 | 0.80 (0.62, 1.04) | 0.81 (0.59, 1.11) | 0.72 (0.45, 1.15) | 0.681 | 0.99 (0.54, 1.81) | 1.01 (0.49, 2.09) | 0.99 (0.32, 3.04) | 0.946 |
| HDL-c | 0.49 (0.41, 0.58) ^***^ | 0.55 (0.44, 0.68) ^***^ | 0.39 (0.29, 0.54) ^***^ | 0.104 | 0.59 (0.41, 0.85) ^**^ | 0.56 (0.36, 0.88) | 0.67 (0.37, 1.21) | 0.626 | 0.62 (0.25, 1.51) | 0.57 (0.19, 1.66) | 0.94 (0.18, 4.89) | 0.699 |
| **Triglycerides (mmol/l)** | **Eulgy_all** | **Eugly_M** | **Eugly_W** | **P-interaction** | **PreDM_all** | **PreDM_M** | **PreDM_W** | **P-interaction** | **T2D_all** | **T2D_M** | **T2D_W** | **P-interaction** |
| Total TG | 1.28 (1.17, 1.39) ^***^ | 1.19 (1.07, 1.33) ^**^ | 1.41 (1.20, 1.65) ^***^ | 0.06 | 0.92 (0.77, 1.10) | 0.92 (0.74, 1.14) | 0.88 (0.65, 1.18) | 0.975 | 1.41 (0.92, 2.16) | 1.10 (0.66, 1.84) | 2.46 (1.06, 5.71) | 0.106 |
| TG in VLDL | 1.19 (1.11, 1.28) ^***^ | 1.13 (1.04, 1.23) ^**^ | 1.29 (1.13, 1.46) ^***^ | 0.077 | 0.92 (0.79, 1.06) | 0.93 (0.77, 1.11) | 0.86 (0.68, 1.09) | 0.765 | 1.35 (0.94, 1.93) | 1.10 (0.71, 1.70) | 2.05 (1.02, 4.13) | 0.135 |
| TG in LDL | 1.66 (1.45, 1.90) ^***^ | 1.55 (1.32, 1.82) ^***^ | 1.85 (1.43, 2.38) ^***^ | 0.193 | 1.09 (0.83, 1.43) | 1.02 (0.73, 1.42) | 1.14 (0.71, 1.84) | 0.589 | 1.50 (0.80, 2.82) | 1.15 (0.54, 2.44) | 3.12 (0.93, 10.53) | 0.146 |
| TG in HDL | 1.29 (1.15, 1.44) ^***^ | 1.16 (1.01, 1.33) ^*^ | 1.55 (1.27, 1.90) ^***^ | 0.012 | 0.92 (0.73, 1.16) | 0.87 (0.66, 1.15) | 0.97 (0.65, 1.44) | 0.511 | 1.35 (0.77, 2.35) | 0.97 (0.50, 1.88) | 3.30 (1.04, 10.49) | 0.067 |
| **Cholesteryl esters (mmol/l)** | **Eulgy_all** | **Eugly_M** | **Eugly_W** | **P-interaction** | **PreDM_all** | **PreDM_M** | **PreDM_W** | **P-interaction** | **T2D_all** | **T2D_M** | **T2D_W** | **P-interaction** |
| Total esterified cholesterol | 0.99 (0.82, 1.19) | 1.03 (0.82, 1.27) | 0.78 (0.54, 1.11) | 0.32 | 0.68 (0.48, 0.96) | 0.67 (0.44, 1.01) | 0.64 (0.35, 1.20) | 0.96 | 0.86 (0.39, 1.93) | 0.88 (0.35, 2.25) | 0.94 (0.19, 4.69) | 0.905 |
| Cholesteryl esters in VLDL | 1.25 (1.12, 1.40) ^***^ | 1.21 (1.06, 1.38) ^**^ | 1.24 (1.03, 1.51) ^**^ | 0.652 | 0.90 (0.73, 1.11) | 0.87 (0.67, 1.14) | 0.89 (0.63, 1.26) | 0.861 | 1.21 (0.75, 1.97) | 1.27 (0.69, 2.32) | 1.12 (0.50, 2.55) | 0.894 |
| Cholesteryl esters in LDL | 1.20 (1.04, 1.38) ^**^ | 1.19 (1.00, 1.40) ^*^ | 1.09 (0.84, 1.43) | 0.805 | 0.80 (0.61, 1.04) | 0.80 (0.58, 1.11) | 0.71 (0.44, 1.14) | 0.697 | 1.03 (0.56, 1.90) | 1.02 (0.49, 2.12) | 1.11 (0.36, 3.41) | 0.816 |
| Cholesteryl esters in HDL | 0.48 (0.40, 0.57) ^***^ | 0.53 (0.43, 0.66) ^***^ | 0.39 (0.29, 0.53) ^***^ | 0.123 | 0.60 (0.42, 0.85) ^**^ | 0.57 (0.37, 0.88) | 0.68 (0.38, 1.21) | 0.62 | 0.59 (0.25, 1.43) | 0.56 (0.19, 1.63) | 0.82 (0.16, 4.16) | 0.789 |
| **Free cholesterol (mmol/l)** | **Eulgy_all** | **Eugly_M** | **Eugly_W** | **P-interaction** | **PreDM_all** | **PreDM_M** | **PreDM_W** | **P-interaction** | **T2D_all** | **T2D_M** | **T2D_W** | **P-interaction** |
| Total free cholesterol | 1.11 (0.93, 1.32) | 1.14 (0.93, 1.40) | 0.90 (0.65, 1.25) | 0.364 | 0.72 (0.52, 1.00) | 0.70 (0.47, 1.04) | 0.70 (0.39, 1.24) | 0.94 | 1.02 (0.48, 2.17) | 1.03 (0.42, 2.51) | 1.10 (0.25, 4.72) | 0.889 |
| Free cholesterol in VLDL | 1.28 (1.16, 1.41) ^***^ | 1.22 (1.08, 1.38) ^**^ | 1.30 (1.10, 1.55) ^**^ | 0.429 | 0.90 (0.74, 1.09) | 0.89 (0.70, 1.14) | 0.87 (0.63, 1.19) | 0.996 | 1.34 (0.85, 2.12) | 1.23 (0.69, 2.18) | 1.55 (0.70, 3.44) | 0.6 |
| Free cholesterol in LDL | 1.05 (0.93, 1.19) | 1.07 (0.92, 1.24) | 0.90 (0.69, 1.16) | 0.371 | 0.85 (0.68, 1.07) | 0.87 (0.67, 1.13) | 0.75 (0.48, 1.17) | 0.558 | 0.91 (0.53, 1.57) | 1.02 (0.53, 1.95) | 0.74 (0.27, 2.03) | 0.675 |
| Free cholesterol in HDL | 0.57 (0.48, 0.68) ^***^ | 0.66 (0.54, 0.82) ^**^ | 0.43 (0.32, 0.59) ^***^ | 0.045 | 0.62 (0.44, 0.89) ^*^ | 0.60 (0.38, 0.94) | 0.67 (0.37, 1.23) | 0.704 | 0.78 (0.33, 1.84) | 0.66 (0.24, 1.82) | 1.52 (0.30, 7.86) | 0.45 |
| **Total lipids (mmol/l)** | **Eulgy_all** | **Eugly_M** | **Eugly_W** | **P-interaction** | **PreDM_all** | **PreDM_M** | **PreDM_W** | **P-interaction** | **T2D_all** | **T2D_M** | **T2D_W** | **P-interaction** |
| Total lipids in lipoprotein particles | 1.24 (1.02, 1.51) ^**^ | 1.23 (0.97, 1.55) | 1.11 (0.76, 1.62) | 0.872 | 0.66 (0.45, 0.95) | 0.63 (0.40, 1.00) | 0.62 (0.32, 1.21) | 0.926 | 1.19 (0.51, 2.81) | 0.91 (0.33, 2.49) | 2.64 (0.50, 13.85) | 0.255 |
| Total lipids in VLDL | 1.27 (1.16, 1.39) ^***^ | 1.20 (1.07, 1.34) ^**^ | 1.34 (1.14, 1.58) ^**^ | 0.191 | 0.91 (0.76, 1.08) | 0.91 (0.72, 1.14) | 0.85 (0.63, 1.15) | 0.885 | 1.39 (0.90, 2.13) | 1.17 (0.69, 1.99) | 1.91 (0.86, 4.24) | 0.296 |
| Total lipids in LDL | 1.22 (1.06, 1.42) ^**^ | 1.21 (1.02, 1.45) ^*^ | 1.11 (0.84, 1.46) | 0.757 | 0.81 (0.61, 1.07) | 0.81 (0.57, 1.14) | 0.74 (0.45, 1.21) | 0.78 | 1.01 (0.52, 1.94) | 1.01 (0.46, 2.22) | 1.06 (0.32, 3.53) | 0.859 |
| Total lipids in HDL mmol/l | 0.51 (0.41, 0.62) ^***^ | 0.55 (0.43, 0.71) ^***^ | 0.43 (0.30, 0.62) ^***^ | 0.338 | 0.53 (0.35, 0.80) ^**^ | 0.50 (0.30, 0.84) | 0.61 (0.31, 1.21) | 0.599 | 0.72 (0.27, 1.91) | 0.52 (0.16, 1.68) | 1.94 (0.30, 12.37) | 0.293 |
| **Phospholipids (mmol/l)** | **Eulgy_all** | **Eugly_M** | **Eugly_W** | **P-interaction** | **PreDM_all** | **PreDM_M** | **PreDM_W** | **P-interaction** | **T2D_all** | **T2D_M** | **T2D_W** | **P-interaction** |
| Total phospholipids | 1.02 (0.81, 1.30) | 1.04 (0.78, 1.38) | 0.86 (0.55, 1.34) | 0.681 | 0.56 (0.36, 0.88) ^*^ | 0.53 (0.31, 0.91) | 0.56 (0.25, 1.25) | 0.804 | 1.06 (0.38, 2.96) | 0.76 (0.23, 2.50) | 3.33 (0.42, 26.22) | 0.215 |
| Phospholipids in VLDL | 1.29 (1.18, 1.42) ^***^ | 1.23 (1.09, 1.38) ^**^ | 1.35 (1.14, 1.59) ^**^ | 0.292 | 0.92 (0.76, 1.11) | 0.91 (0.72, 1.16) | 0.88 (0.65, 1.20) | 0.972 | 1.37 (0.88, 2.14) | 1.22 (0.70, 2.13) | 1.68 (0.76, 3.72) | 0.482 |
| Phospholipids in LDL | 1.23 (1.05, 1.44) ^**^ | 1.22 (1.01, 1.46) ^*^ | 1.12 (0.84, 1.49) | 0.803 | 0.82 (0.61, 1.10) | 0.81 (0.56, 1.17) | 0.76 (0.45, 1.26) | 0.821 | 0.97 (0.49, 1.95) | 1.01 (0.44, 2.32) | 0.96 (0.27, 3.39) | 0.963 |
| Phospholipids in HDL | 0.53 (0.43, 0.65) ^***^ | 0.57 (0.44, 0.73) ^***^ | 0.48 (0.33, 0.69) ^***^ | 0.556 | 0.54 (0.36, 0.81) ^**^ | 0.51 (0.31, 0.85) | 0.60 (0.30, 1.21) | 0.639 | 0.80 (0.30, 2.12) | 0.54 (0.17, 1.69) | 2.68 (0.41, 17.53) | 0.188 |
| **Apolipoproteins (g/l)** | **Eulgy_all** | **Eugly_M** | **Eugly_W** | **P-interaction** | **PreDM_all** | **PreDM_M** | **PreDM_W** | **P-interaction** | **T2D_all** | **T2D_M** | **T2D_W** | **P-interaction** |
| Apolipoprotein B | 1.33 (1.14, 1.55) ^**^ | 1.33 (1.10, 1.61) ^**^ | 1.17 (0.89, 1.55) | 0.611 | 0.85 (0.63, 1.15) | 0.83 (0.57, 1.20) | 0.82 (0.49, 1.36) | 0.998 | 1.15 (0.57, 2.32) | 1.27 (0.54, 3.00) | 0.99 (0.29, 3.35) | 0.831 |
| Apolipoprotein A1 | 0.47 (0.37, 0.61) ^***^ | 0.49 (0.36, 0.66) ^***^ | 0.44 (0.29, 0.69) ^**^ | 0.841 | 0.45 (0.27, 0.73) ^**^ | 0.41 (0.22, 0.77) | 0.52 (0.22, 1.21) | 0.605 | 0.63 (0.19, 2.08) | 0.40 (0.10, 1.64) | 2.55 (0.27, 24.42) | 0.21 |
| Apo(B) to apo(A1) ratio | 1.54 (1.35, 1.76) ^***^ | 1.55 (1.31, 1.83) ^***^ | 1.40 (1.11, 1.76) ^**^ | 0.553 | 1.12 (0.85, 1.46) | 1.12 (0.80, 1.58) | 1.03 (0.67, 1.58) | 0.747 | 1.29 (0.67, 2.46) | 1.69 (0.75, 3.82) | 0.81 (0.28, 2.33) | 0.361 |
| **Lipoprotein particle size (nm)** | **Eulgy_all** | **Eugly_M** | **Eugly_W** | **P-interaction** | **PreDM_all** | **PreDM_M** | **PreDM_W** | **P-interaction** | **T2D_all** | **T2D_M** | **T2D_W** | **P-interaction** |
| Average diameter for VLDL particles | 7.58 (2.30, 24.92) ^**^ | 2.75 (0.66, 11.45) | 40.71 (4.65, 356.17) ^**^ | 0.035 | 0.13 (0.01, 1.43) | 0.28 (0.01, 5.40) | 0.02 (0.00, 1.12) | 0.365 | 63.94 (0.29, 1.43e+04) | 2.21 (2.78e-03, 1.76e+03) | 4.39e+04 (1.41, 1.37e+09) | 0.129 |
| Average diameter for LDL particles | 35.10 (2.13e-03, 5.78e+05) | 2.21e+02 (2.61e-03, 1.88e+07) | 0.08 (5.65e-10, 9.98e+06) | 0.439 | 1.25e-03 (6.99e-12, 2.22e+05) | 9.22 (9.66e-10, 8.81e+10) | 0.00 (0.00, 4400.13) | 0.133 | 9.92e-06 (2.50e-25, 3.94e+14) | 7.77e+06 (8.17e-18, 7.38e+30) | 1.58e-26 (4.29e-62, 5.81e+09) | 0.16 |
| Average diameter for HDL particles | 1.34e-04 (1.64e-05, 1.10e-03) ^***^ | 2.43e-03 (1.75e-04, 3.37e-02) ^***^ | 2.93e-06 (9.06e-08, 9.48e-05) ^***^ | 0.003 | 0.02 (0.00, 1.92) | 0.01 (2.16e-05, 3.09) | 0.20 (0.00, 206.14) | 0.445 | 0.12 (1.20e-06, 1.21e+04) | 3.59 (2.00e-06, 6.44e+06) | 1.58e-26 (4.29e-62, 5.81e+09) | 0.519 |
| **Total concentration of lipoprotein particles** | 0.62 (0.48, 0.80) ^**^ | 0.60 (0.44, 0.81) ^**^ | 0.61 (0.37, 0.99) ^*^ | 0.768 | 0.44 (0.26, 0.72) ^**^ | 0.42 (0.23, 0.77) | 0.45 (0.18, 1.11) | 0.833 | 0.60 (0.18, 1.95) | 0.38 (0.10, 1.55) | 2.37 (0.24, 23.65) | 0.207 |
| Concentration of VLDL particles | 1.40 (1.24, 1.57) ^***^ | 1.33 (1.15, 1.54) ^**^ | 1.42 (1.15, 1.76) ^**^ | 0.479 | 0.92 (0.73, 1.17) | 0.89 (0.66, 1.20) | 0.90 (0.61, 1.33) | 0.861 | 1.41 (0.81, 2.47) | 1.29 (0.65, 2.59) | 1.65 (0.63, 4.35) | 0.623 |
| Concentration of LDL particles | 1.32 (1.13, 1.54) ^**^ | 1.34 (1.11, 1.62) ^**^ | 1.13 (0.86, 1.50) | 0.465 | 0.85 (0.63, 1.14) | 0.83 (0.57, 1.21) | 0.79 (0.48, 1.32) | 0.896 | 1.14 (0.56, 2.29) | 1.26 (0.53, 2.97) | 0.98 (0.29, 3.30) | 0.827 |
| Concentration of HDL particles | 0.56 (0.43, 0.71) ^***^ | 0.54 (0.40, 0.72) ^***^ | 0.57 (0.36, 0.90) ^**^ | 0.683 | 0.45 (0.28, 0.73) ^**^ | 0.43 (0.24, 0.78) | 0.48 (0.20, 1.11) | 0.811 | 0.58 (0.19, 1.79) | 0.36 (0.09, 1.39) | 2.28 (0.26, 20.19) | 0.188 |
| **Lipoprotein size** | **Eulgy_all** | **Eugly_M** | **Eugly_W** | **P-interaction** | **PreDM_all** | **PreDM_M** | **PreDM_W** | **P-interaction** | **T2D_all** | **T2D_M** | **T2D_W** | **P-interaction** |
| XXL_VLDL | 1.10 (1.05, 1.14) ^***^ | 1.05 (1.00, 1.10) | 1.16 (1.08, 1.24) ^***^ | 0.017 | 0.96 (0.88, 1.04) | 0.95 (0.85, 1.06) | 0.95 (0.83, 1.08) | 0.902 | 1.20 (0.95, 1.51) | 1.12 (0.84, 1.48) | 1.34 (0.85, 2.10) | 0.515 |
| XL_VLDL | 1.17 (1.10, 1.24) ^***^ | 1.11 (1.03, 1.20) ^**^ | 1.21 (1.09, 1.34) ^**^ | 0.158 | 0.94 (0.83, 1.06) | 0.97 (0.82, 1.14) | 0.89 (0.75, 1.05) | 0.549 | 1.28 (0.93, 1.76) | 1.21 (0.79, 1.84) | 1.34 (0.79, 2.26) | 0.732 |
| L_VLDL | 1.20 (1.12, 1.29) ^***^ | 1.14 (1.05, 1.25) ^**^ | 1.25 (1.11, 1.41) ^**^ | 0.169 | 0.93 (0.81, 1.08) | 0.96 (0.80, 1.17) | 0.87 (0.71, 1.07) | 0.562 | 1.29 (0.90, 1.86) | 1.20 (0.75, 1.94) | 1.38 (0.76, 2.49) | 0.68 |
| M_VLDL | 1.08 (0.99, 1.18) | 1.08 (0.97, 1.19) | 1.00 (0.85, 1.18) | 0.638 | 0.87 (0.74, 1.03) | 0.87 (0.71, 1.06) | 0.84 (0.63, 1.12) | 0.895 | 1.06 (0.76, 1.49) | 1.19 (0.79, 1.79) | 0.86 (0.48, 1.56) | 0.425 |
| S_VLDL | 1.33 (1.19, 1.48) ^***^ | 1.27 (1.11, 1.46) ^**^ | 1.35 (1.11, 1.63) ^**^ | 0.499 | 0.95 (0.77, 1.19) | 0.92 (0.70, 1.22) | 0.94 (0.66, 1.34) | 0.865 | 1.21 (0.72, 2.01) | 1.28 (0.67, 2.45) | 1.11 (0.49, 2.52) | 0.888 |
| XS_VLDL | 1.21 (1.05, 1.39) ^**^ | 1.21 (1.02, 1.44) ^**^ | 1.10 (0.85, 1.42) | 0.721 | 0.96 (0.74, 1.24) | 0.89 (0.65, 1.23) | 1.01 (0.64, 1.60) | 0.605 | 1.13 (0.63, 2.03) | 1.25 (0.63, 2.50) | 0.94 (0.31, 2.82) | 0.743 |
| IDL | 0.99 (0.86, 1.14) | 1.01 (0.85, 1.20) | 0.84 (0.64, 1.11) | 0.414 | 0.83 (0.64, 1.09) | 0.82 (0.60, 1.13) | 0.79 (0.49, 1.28) | 0.909 | 0.87 (0.48, 1.58) | 1.00 (0.51, 1.97) | 0.63 (0.18, 2.17) | 0.593 |
| L_LDL | 1.12 (0.97, 1.28) | 1.12 (0.95, 1.32) | 0.98 (0.74, 1.28) | 0.565 | 0.94 (0.81, 1.08) | 0.96 (0.81, 1.13) | 0.70 (0.43, 1.14) | 0.247 | 0.95 (0.52, 1.74) | 0.98 (0.48, 2.01) | 0.94 (0.30, 2.98) | 0.971 |
| M_LDL | 1.21 (1.07, 1.38) ^**^ | 1.18 (1.01, 1.38) ^*^ | 1.15 (0.91, 1.46) | 0.964 | 0.81 (0.64, 1.04) | 0.80 (0.60, 1.09) | 0.76 (0.50, 1.16) | 0.841 | 1.04 (0.60, 1.83) | 1.04 (0.52, 2.06) | 1.09 (0.40, 2.96) | 0.854 |
| S_LDL | 1.24 (1.07, 1.44) ^**^ | 1.25 (1.05, 1.50) ^**^ | 1.09 (0.83, 1.42) | 0.512 | 0.80 (0.60, 1.07) | 0.79 (0.56, 1.13) | 0.75 (0.46, 1.21) | 0.853 | 1.09 (0.57, 2.09) | 1.15 (0.52, 2.56) | 1.02 (0.32, 3.19) | 0.953 |
| XL_HDL | 0.75 (0.69, 0.82) ^***^ | 0.88 (0.78, 0.98) ^*^ | 0.57 (0.49, 0.66) ^***^ | <0.001 | 0.95 (0.79, 1.14) | 0.96 (0.76, 1.22) | 0.92 (0.67, 1.27) | 0.837 | 1.07 (0.71, 1.62) | 1.35 (0.80, 2.27) | 0.70 (0.32, 1.52) | 0.16 |
| L_HDL | 0.80 (0.76, 0.84) ^***^ | 0.84 (0.79, 0.90) ^***^ | 0.64 (0.57, 0.73) ^***^ | <0.001 | 0.94 (0.83, 1.05) | 0.95 (0.83, 1.09) | 0.90 (0.70, 1.15) | 0.723 | 0.95 (0.71, 1.27) | 1.00 (0.71, 1.42) | 0.88 (0.50, 1.57) | 0.643 |
| M_HDL | 0.73 (0.68, 0.78) ^***^ | 0.74 (0.68, 0.81) ^***^ | 0.55 (0.41, 0.73) ^***^ | 0.063 | 0.62 (0.45, 0.86) ^**^ | 0.61 (0.41, 0.90) | 0.67 (0.39, 1.16) | 0.747 | 0.65 (0.30, 1.41) | 0.51 (0.21, 1.27) | 1.39 (0.32, 6.03) | 0.316 |
| S_HDL | 0.97 (0.75, 1.27) | 0.71 (0.51, 0.98) ^*^ | 1.67 (1.04, 2.69) ^*^ | 0.003 | 0.53 (0.31, 0.89) ^*^ | 0.52 (0.27, 0.98) | 0.49 (0.19, 1.24) | 0.915 | 0.55 (0.16, 1.84) | 0.28 (0.07, 1.22) | 2.59 (0.27, 24.86) | 0.111 |
| **Other lipids (mmol/L**) | **Eulgy_all** | **Eugly_M** | **Eugly_W** | **P-interaction** | **PreDM_all** | **PreDM_M** | **PreDM_W** | **P-interaction** | **T2D_all** | **T2D_M** | **T2D_W** | **P-interaction** |
| Phosphoglycerides | 0.96 (0.77, 1.20) | 0.95 (0.73, 1.23) | 0.90 (0.59, 1.36) | 0.962 | 0.57 (0.38, 0.87) ^*^ | 0.56 (0.34, 0.92) | 0.54 (0.25, 1.16) | 0.926 | 0.99 (0.39, 2.52) | 0.67 (0.23, 1.98) | 3.56 (0.53, 24.03) | 0.134 |
| TG to phosphoglycerides ratio | 1.39 (1.26, 1.54) ^***^ | 1.28 (1.13, 1.44) ^***^ | 1.58 (1.32, 1.89) ^***^ | 0.044 | 1.02 (0.83, 1.26) | 1.03 (0.80, 1.34) | 0.95 (0.68, 1.34) | 0.85 | 1.71 (1.00, 2.93) | 1.35 (0.70, 2.61) | 2.71 (0.96, 7.61) | 0.265 |
| Total cholines | 0.91 (0.71, 1.16) | 0.92 (0.69, 1.23) | 0.77 (0.49, 1.22) | 0.734 | 0.53 (0.34, 0.85) ^*^ | 0.51 (0.29, 0.90) | 0.52 (0.23, 1.19) | 0.899 | 0.93 (0.33, 2.67) | 0.65 (0.19, 2.21) | 3.18 (0.38, 26.41) | 0.202 |
| Phosphatidylcholines | 0.90 (0.73, 1.12) | 0.90 (0.70, 1.17) | 0.82 (0.54, 1.23) | 0.906 | 0.55 (0.36, 0.83) ^**^ | 0.54 (0.32, 0.88) | 0.52 (0.25, 1.10) | 0.918 | 1.05 (0.41, 2.66) | 0.78 (0.27, 2.28) | 2.82 (0.43, 18.71) | 0.236 |
| Sphingomyelins | 0.97 (0.76, 1.23) | 1.07 (0.80, 1.42) | 0.67 (0.42, 1.05) | 0.163 | 0.74 (0.46, 1.18) | 0.69 (0.39, 1.22) | 0.78 (0.34, 1.79) | 0.748 | 0.68 (0.22, 2.05) | 0.68 (0.19, 2.49) | 0.85 (0.09, 8.05) | 0.832 |
| **Amino acids (mmol/L)** | **Eulgy_all** | **Eugly_M** | **Eugly_W** | **P-interaction** | **PreDM_all** | **PreDM_M** | **PreDM_W** | **P-interaction** | **T2D_all** | **T2D_M** | **T2D_W** | **P-interaction** |
| Alanine | 1.07 (0.93, 1.23) | 1.04 (0.88, 1.23) | 1.13 (0.88, 1.45) | 0.532 | 1.03 (0.77, 1.37) | 1.16 (0.82, 1.65) | 0.79 (0.48, 1.31) | 0.261 | 1.03 (0.53, 1.98) | 0.81 (0.37, 1.77) | 2.05 (0.55, 7.60) | 0.248 |
| Glutamine | 0.99 (0.78, 1.25) | 1.10 (0.83, 1.46) | 0.71 (0.46, 1.09) | 0.14 | 1.55 (0.97, 2.48) | 1.57 (0.88, 2.78) | 1.49 (0.65, 3.42) | 0.959 | 1.78 (0.63, 5.07) | 2.02 (0.57, 7.12) | 2.41 (0.35, 16.48) | 0.917 |
| Glycine | 0.81 (0.74, 0.89) ^***^ | 0.78 (0.70, 0.87) ^***^ | 0.88 (0.76, 1.03) | 0.18 | 1.04 (0.85, 1.27) | 1.09 (0.83, 1.41) | 1.01 (0.75, 1.38) | 0.697 | 0.82 (0.57, 1.19) | 0.93 (0.59, 1.47) | 0.74 (0.36, 1.52) | 0.529 |
| Histidine | 0.92 (0.73, 1.15) | 0.99 (0.75, 1.30) | 0.78 (0.52, 1.16) | 0.325 | 0.63 (0.41, 0.97) | 0.65 (0.38, 1.11) | 0.64 (0.31, 1.33) | 0.954 | 1.85 (0.65, 5.32) | 1.46 (0.40, 5.25) | 5.29 (0.65, 42.90) | 0.412 |
| **Aromatic amino acids (mmol/L)** | **Eulgy_all** | **Eugly_M** | **Eugly_W** | **P-interaction** | **PreDM_all** | **PreDM_M** | **PreDM_W** | **P-interaction** | **T2D_all** | **T2D_M** | **T2D_W** | **P-interaction** |
| Phenylalanine | 1.31 (1.12, 1.53) ^**^ | 1.30 (1.08, 1.58) ^**^ | 1.31 (1.00, 1.73) ^*^ | 0.933 | 1.63 (1.17, 2.25) ^**^ | 1.60 (1.07, 2.42) | 1.72 (1.00, 2.95) | 0.806 | 1.06 (0.47, 2.36) | 0.80 (0.31, 2.09) | 1.94 (0.43, 8.70) | 0.277 |
| Tyrosine | 1.30 (1.11, 1.54) ^**^ | 1.35 (1.10, 1.65) ^**^ | 1.20 (0.91, 1.60) | 0.668 | 1.07 (0.77, 1.48) | 1.03 (0.68, 1.55) | 1.17 (0.67, 2.02) | 0.669 | 0.73 (0.33, 1.59) | 0.46 (0.18, 1.16) | 2.45 (0.51, 11.88) | 0.064 |
| **Branched-chain amino acids (BCAA) (mmol/L)** | **Eulgy_all** | **Eugly_M** | **Eugly_W** | **P-interaction** | **PreDM_all** | **PreDM_M** | **PreDM_W** | **P-interaction** | **T2D_all** | **T2D_M** | **T2D_W** | **P-interaction** |
| Isoleucine | 1.23 (1.11, 1.38) ^***^ | 1.24 (1.08, 1.43) ^**^ | 1.19 (0.99, 1.43) | 0.727 | 0.92 (0.74, 1.16) | 0.86 (0.64, 1.14) | 1.05 (0.73, 1.52) | 0.405 | 1.26 (0.73, 2.16) | 1.25 (0.65, 2.41) | 1.23 (0.41, 3.64) | 0.916 |
| Leucine | 1.25 (1.08, 1.44) ^**^ | 1.17 (0.98, 1.40) | 1.33 (1.05, 1.68) ^**^ | 0.402 | 0.95 (0.72, 1.26) | 0.79 (0.55, 1.13) | 1.29 (0.81, 2.04) | 0.109 | 1.35 (0.69, 2.63) | 1.41 (0.63, 3.17) | 1.24 (0.34, 4.60) | 0.961 |
| Valine | 1.21 (1.00, 1.46) ^*^ | 1.16 (0.92, 1.46) | 1.22 (0.89, 1.68) | 0.716 | 0.77 (0.53, 1.13) | 0.69 (0.43, 1.12) | 0.93 (0.49, 1.74) | 0.435 | 0.99 (0.41, 2.41) | 0.73 (0.24, 2.18) | 2.06 (0.38, 11.07) | 0.259 |
| Total BCAA | 1.26 (1.07, 1.49) ^**^ | 1.22 (0.99, 1.49) | 1.28 (0.97, 1.69) | 0.714 | 0.85 (0.61, 1.19) | 0.73 (0.48, 1.12) | 1.08 (0.62, 1.87) | 0.271 | 1.19 (0.55, 2.60) | 1.06 (0.41, 2.75) | 1.63 (0.36, 7.37) | 0.555 |
| **Ketones (mmol/L)** | **Eulgy_all** | **Eugly_M** | **Eugly_W** | **P-interaction** | **PreDM_all** | **PreDM_M** | **PreDM_W** | **P-interaction** | **T2D_all** | **T2D_M** | **T2D_W** | **P-interaction** |
| 3-hydroxybutyrate | 1.01 (0.96, 1.06) | 1.02 (0.96, 1.08) | 0.99 (0.92, 1.07) | 0.679 | 1.00 (0.89, 1.11) | 0.90 (0.78, 1.04) | 1.16 (0.98, 1.38) | 0.022 | 0.89 (0.67, 1.17) | 1.12 (0.76, 1.63) | 0.68 (0.49, 0.95) | 0.055 |
| Acetoacetate | 1.04 (1.00, 1.09) ^*^ | 1.05 (1.00, 1.11) | 1.02 (0.95, 1.10) | 0.693 | 1.01 (0.92, 1.10) | 0.97 (0.87, 1.09) | 1.09 (0.92, 1.28) | 0.229 | 0.98 (0.77, 1.25) | 1.09 (0.81, 1.47) | 0.73 (0.46, 1.17) | 0.154 |
| Acetone | 0.96 (0.85, 1.08) | 1.00 (0.87, 1.15) | 0.87 (0.70, 1.08) | 0.336 | 0.85 (0.66, 1.09) | 0.70 (0.51, 0.96) | 1.26 (0.82, 1.92) | 0.023 | 1.09 (0.56, 2.11) | 2.27 (1.06, 4.85) | 0.13 (0.03, 0.58) | 0.001 |
| Acetate | 0.96 (0.90, 1.03) | 1.01 (0.93, 1.10) | 0.87 (0.76, 0.98) ^**^ | 0.054 | 0.92 (0.80, 1.06) | 0.90 (0.75, 1.07) | 0.99 (0.77, 1.28) | 0.548 | 0.75 (0.53, 1.05) | 0.69 (0.47, 1.01) | 1.14 (0.53, 2.44) | 0.247 |
| **Inflammation (mmol/l)** | **Eulgy_all** | **Eugly_M** | **Eugly_W** | **P-interaction** | **PreDM_all** | **PreDM_M** | **PreDM_W** | **P-interaction** | **T2D_all** | **T2D_M** | **T2D_W** | **P-interaction** |
| Glycoprotein acetyls (GlycA) | 2.98 (2.31, 3.85) ^***^ | 2.44 (1.80, 3.31) ^***^ | 4.58 (2.83, 7.41) ^***^ | 0.023 | 2.07 (1.22, 3.51) ^*^ | 1.99 (1.04, 3.81) | 2.12 (0.84, 5.38) | 0.835 | 0.89 (0.25, 3.19) | 0.42 (0.09, 1.93) | 4.41 (0.36, 54.02) | 0.081 |
| **Renal function (mmol/l )** | **Eulgy_all** | **Eugly_M** | **Eugly_W** | **P-interaction** | **PreDM_all** | **PreDM_M** | **PreDM_W** | **P-interaction** | **T2D_all** | **T2D_M** | **T2D_W** | **P-interaction** |
| Creatinine | 1.65 (1.33, 2.04) ^***^ | 1.56 (1.21, 2.02) ^**^ | 1.70 (1.17, 2.47) ^**^ | 0.635 | 1.50 (0.98, 2.30) | 1.03 (0.59, 1.78) | 2.72 (1.40, 5.27) | 0.02 | 1.79 (0.69, 4.67) | 2.41 (0.74, 7.85) | 1.08 (0.17, 6.68) | 0.435 |
| Albumin | 0.45 (0.30, 0.68) ^***^ | 0.47 (0.28, 0.77) ^**^ | 0.36 (0.17, 0.77) ^**^ | 0.564 | 0.36 (0.16, 0.81) ^*^ | 0.40 (0.15, 1.06) | 0.30 (0.08, 1.22) | 0.762 | 0.65 (0.10, 4.36) | 0.11 (0.01, 1.19) | 31.13 (0.94, 1032.66) | 0.011 |
| **Fatty acids (mmol/l)** | **Eulgy_all** | **Eugly_M** | **Eugly_W** | **P-interaction** | **PreDM_all** | **PreDM_M** | **PreDM_W** | **P-interaction** | **T2D_all** | **T2D_M** | **T2D_W** | **P-interaction** |
| Total fatty acids | 1.44 (1.20, 1.73) ^***^ | 1.35 (1.09, 1.67) ^**^ | 1.55 (1.09, 2.21) ^**^ | 0.376 | 0.74 (0.52, 1.06) | 0.72 (0.47, 1.11) | 0.69 (0.36, 1.32) | 0.969 | 1.34 (0.61, 2.94) | 0.91 (0.36, 2.29) | 4.22 (0.86, 20.81) | 0.094 |
| Omega-3 fatty acids | 1.00 (0.92, 1.09) | 1.02 (0.92, 1.13) | 0.93 (0.79, 1.10) | 0.647 | 0.84 (0.71, 0.99) | 0.85 (0.70, 1.05) | 0.76 (0.55, 1.04) | 0.686 | 0.88 (0.58, 1.34) | 0.78 (0.48, 1.28) | 1.22 (0.51, 2.89) | 0.461 |
| Omega-6 fatty acids | 1.22 (0.96, 1.56) | 1.27 (0.95, 1.69) | 0.95 (0.60, 1.50) | 0.422 | 0.59 (0.37, 0.95) | 0.59 (0.33, 1.05) | 0.51 (0.22, 1.17) | 0.744 | 1.13 (0.37, 3.40) | 1.04 (0.29, 3.79) | 1.72 (0.20, 14.81) | 0.67 |
| Polyunsaturated fatty acids (PUFA) | 1.17 (0.93, 1.47) | 1.21 (0.92, 1.59) | 0.91 (0.59, 1.41) | 0.461 | 0.57 (0.37, 0.90) ^*^ | 0.59 (0.34, 1.02) | 0.47 (0.21, 1.05) | 0.659 | 0.96 (0.34, 2.72) | 0.83 (0.25, 2.80) | 1.73 (0.22, 13.55) | 0.548 |
| Monounsaturated fatty acids (MUFA) | 1.46 (1.28, 1.66) ^***^ | 1.33 (1.14, 1.55) ^**^ | 1.76 (1.37, 2.26) ^***^ | 0.043 | 0.88 (0.68, 1.13) | 0.86 (0.63, 1.17) | 0.84 (0.53, 1.36) | 0.888 | 1.42 (0.81, 2.51) | 1.05 (0.54, 2.04) | 3.32 (1.02, 10.81) | 0.087 |
| Saturated fatty acids (SFA) | 1.39 (1.19, 1.63) ^***^ | 1.28 (1.06, 1.54) ^**^ | 1.59 (1.17, 2.17) ^**^ | 0.164 | 0.81 (0.60, 1.10) | 0.77 (0.53, 1.11) | 0.81 (0.46, 1.43) | 0.713 | 1.36 (0.69, 2.70) | 0.89 (0.40, 1.99) | 5.07 (1.21, 21.16) | 0.033 |
| Linoleic acid | 1.06 (0.88, 1.27) | 1.10 (0.88, 1.36) | 0.85 (0.61, 1.20) | 0.318 | 0.65 (0.46, 0.93) ^*^ | 0.68 (0.44, 1.04) | 0.55 (0.30, 1.02) | 0.563 | 1.18 (0.53, 2.65) | 1.21 (0.47, 3.13) | 1.23 (0.26, 5.79) | 0.921 |
| Docosahexaenoic acid (DHA) | 0.90 (0.82, 1.00) ^*^ | 0.95 (0.85, 1.07) | 0.75 (0.61, 0.92) ^**^ | 0.108 | 0.85 (0.70, 1.03) | 0.88 (0.70, 1.10) | 0.75 (0.52, 1.08) | 0.557 | 0.67 (0.41, 1.11) | 0.61 (0.34, 1.09) | 0.90 (0.32, 2.53) | 0.581 |
| **Fatty acid ratio (%)** | **Eulgy_all** | **Eugly_M** | **Eugly_W** | **P-interaction** | **PreDM_all** | **PreDM_M** | **PreDM_W** | **P-interaction** | **T2D_all** | **T2D_M** | **T2D_W** | **P-interaction** |
| Omega-3 fatty acids to total fatty acids | 0.90 (0.81, 0.99) ^*^ | 0.93 (0.83, 1.05) | 0.80 (0.67, 0.97) ^**^ | 0.333 | 0.86 (0.70, 1.05) | 0.89 (0.69, 1.14) | 0.77 (0.53, 1.12) | 0.644 | 0.72 (0.43, 1.21) | 0.70 (0.38, 1.30) | 0.70 (0.24, 2.05) | 0.875 |
| Omega-6 fatty acids to total fatty acids | 0.38 (0.27, 0.54) ^***^ | 0.54 (0.36, 0.82) ^**^ | 0.15 (0.07, 0.29) ^***^ | 0.001 | 1.01 (0.52, 1.99) | 1.14 (0.51, 2.54) | 0.87 (0.24, 3.13) | 0.495 | 0.42 (0.09, 1.87) | 1.52 (0.25, 9.19) | 1.32e-02 (6.42e-04, 0.27) | 0.009 |
| PUFA to total fatty acids | 0.30 (0.20, 0.44) ^***^ | 0.46 (0.30, 0.71) ^**^ | 0.09 (0.04, 0.18) ^***^ | <0.001 | 0.83 (0.40, 1.73) | 1.03 (0.43, 2.44) | 0.58 (0.15, 2.28) | 0.333 | 0.23 (0.04, 1.24) | 0.93 (0.13, 6.57) | 2.69e-03 (6.49e-05, 0.11) | 0.006 |
| MUFA to total fatty acids | 3.63 (2.59, 5.09) ^***^ | 2.44 (1.64, 3.63) ^***^ | 9.06 (4.77, 17.19) ^***^ | 0.001 | 1.21 (0.61, 2.42) | 1.16 (0.50, 2.66) | 1.22 (0.36, 4.20) | 0.772 | 4.53 (0.93, 22.14) | 2.06 (0.32, 13.24) | 27.90 (1.02, 760.89) | 0.179 |
| Saturated fatty acids to total fatty acids | 2.12 (1.14, 3.93) ^**^ | 1.41 (0.69, 2.88) | 6.93 (2.02, 23.74) ^**^ | 0.023 | 1.19 (0.37, 3.88) | 0.72 (0.18, 2.92) | 3.11 (0.35, 27.86) | 0.163 | 4.81 (0.26, 87.72) | 0.45 (0.01, 13.96) | 6131.70 (11.09, 3.39e+06) | 0.01 |
| Linoleic acid to total fatty acids | 0.44 (0.33, 0.59) ^***^ | 0.58 (0.41, 0.82) ^**^ | 0.20 (0.12, 0.35) ^***^ | 0.001 | 0.71 (0.40, 1.24) | 0.86 (0.44, 1.68) | 0.47 (0.17, 1.32) | 0.211 | 0.67 (0.18, 2.54) | 2.22 (0.45, 10.88) | 0.04 (0.00, 0.52) | 0.012 |
| DHA to total fatty acids | 0.81 (0.73, 0.89) ^***^ | 0.88 (0.78, 0.98) ^**^ | 0.64 (0.52, 0.78) ^***^ | 0.014 | 0.93 (0.77, 1.13) | 0.97 (0.77, 1.21) | 0.85 (0.59, 1.22) | 0.573 | 0.60 (0.37, 0.98) | 0.64 (0.36, 1.13) | 0.51 (0.19, 1.37) | 0.598 |
| PUFA to MUFA ratio | 0.50 (0.42, 0.61) ^***^ | 0.63 (0.51, 0.79) ^***^ | 0.28 (0.20, 0.41) ^***^ | <0.001 | 0.90 (0.62, 1.31) | 0.97 (0.62, 1.50) | 0.82 (0.42, 1.63) | 0.523 | 0.45 (0.19, 1.05) | 0.80 (0.30, 2.14) | 0.09 (0.01, 0.55) | 0.038 |
| Omega-6 fatty acids to omega-3 fatty acids | 1.03 (0.94, 1.13) | 1.01 (0.91, 1.13) | 1.08 (0.90, 1.28) | 0.841 | 1.14 (0.95, 1.38) | 1.12 (0.89, 1.41) | 1.24 (0.87, 1.75) | 0.807 | 1.20 (0.75, 1.92) | 1.40 (0.80, 2.45) | 0.87 (0.33, 2.30) | 0.507 |
| Degree of unsaturation | 0.22 (0.12, 0.41) ^***^ | 0.40 (0.19, 0.81) | 0.05 (0.02, 0.16) ^***^ | 0.006 | 0.58 (0.18, 1.94) | 0.77 (0.18, 3.28) | 0.35 (0.04, 2.99) | 0.429 | 0.04 (0.00, 0.77) | 0.23 (0.01, 6.38) | 1.95e-04 (2.22e-07, 0.17) | 0.063 |

FDR-adjusted p*<.1; ** <.05; ***<.0001

Fully adjusted for age, race, income, area deprivation, smoking, alcohol drinking, physical activity, BMI, and medications for hypertension and diabetes, and additionally adjusted for menopausal status for women.
